# A Systematic Review of the use of Precision Diagnostics in Monogenic Diabetes

**DOI:** 10.1101/2023.04.15.23288269

**Authors:** Rinki Murphy, Kevin Colclough, Toni I. Pollin, Jennifer M. Ikle, Pernille Svalastoga, Kristin A. Maloney, Cécile Saint-Martin, Janne Molnes, ADA/EASD Precision Medicine Diabetes Initiative, Shivani Misra, Ingvild Aukrust, AIElisa de Franco, Sarah E. Flanagan, Pål R. Njølstad, Liana K. Billings, Katharine R Owen, Anna L Gloyn

**Affiliations:** Department of Medicine, Faculty of Medical and Health Sciences, University of Auckland, Auckland, New Zealand; Auckland Diabetes Centre, Te Whatu Ora Health New Zealand, Te Tokai Tumai, Auckland, New Zealand; Exeter Genomics Laboratory, Royal Devon University Healthcare NHS Foundation Trust, Exeter, United Kingdom; Department of Medicine, University of Maryland School of Medicine, Baltimore, Maryland, USA; Department of Pediatrics, Division of Endocrinology & Diabetes, Stanford School of Medicine, Stanford, CA, USA; Stanford Diabetes Research Center, Stanford School of Medicine, Stanford, CA, USA; Children and Youth Clinic, Haukeland University Hospital, Bergen, Norway; Mohn Center for Diabetes Precision Medicine, Department of Clinical Science, University of Bergen, Bergen, Norway; Department of Medical Genetics, AP-HP Pitié-Salpêtrière Hospital, Sorbonne University, Paris, France; Department of Medical Genetics, Haukeland University Hospital, Bergen, Norway; American Diabetes Association/European Association for the Study of Diabetes Precision Medicine Initiative; Department of Metabolism, Digestion and Reproduction, Imperial College London, London, UK; Department of Diabetes and Endocrinology, Imperial College Healthcare NHS Trust, London, UK; Department of Clinical and Biomedical Science, Faculty of Health and Life Sciences, University of Exeter, UK; Division of Endocrinology, NorthShore University HealthSystem, Skokie, IL, USA; Department of Medicine, Pritzker School of Medicine, University of Chicago, Chicago, IL, USA; Oxford Center for Diabetes, Endocrinology & Metabolism, University of Oxford, UK; NIHR Oxford Biomedical Research Centre, Oxford, UK; Department of Genetics, Stanford School of Medicine, Stanford, CA, USA

## Abstract

Monogenic forms of diabetes present opportunities for precision medicine as identification of the underlying genetic cause has implications for treatment and prognosis. However, genetic testing remains inconsistent across countries and health providers, often resulting in both missed diagnosis and misclassification of diabetes type. One of the barriers to deploying genetic testing is uncertainty over whom to test as the clinical features for monogenic diabetes overlap with those for both type 1 and type 2 diabetes. In this review, we perform a systematic evaluation of the evidence for the clinical and biochemical criteria used to guide selection of individuals with diabetes for genetic testing and review the evidence for the optimal methods for variant detection in genes involved in monogenic diabetes. In parallel we revisit the current clinical guidelines for genetic testing for monogenic diabetes and provide expert opinion on the interpretation and reporting of genetic tests. We provide a series of recommendations for the field informed by our systematic review, synthesizing evidence, and expert opinion. Finally, we identify major challenges for the field and highlight areas for future research and investment to support wider implementation of precision diagnostics for monogenic diabetes.

**Plan Language Summary:** Since monogenic diabetes misclassification can occur and lead to missed opportunities for optimal management, and several diagnostic technologies are available, we systematically review the yield of monogenic diabetes using different criteria to select people with diabetes for genetic testing and the technologies used.

## Introduction

The use of precision diabetes medicine has gained increased awareness to improve diagnosis and treatment for patients with diabetes^1^. While the majority of those living with diabetes globally have polygenic disorders categorized as type 1 diabetes (the predominant form in those diagnosed in childhood and early adulthood), or type 2 diabetes (the predominant form in older people), approximately 1-2% have monogenic forms of diabetes, which is most commonly found in diabetes onset in neonates through to young adulthood^2^. Knowledge of the exact molecular defect and mechanism of disease is crucial for precision diagnostics, which informs treatment, prognostics, and monitoring. Monogenic diabetes types, such as neonatal diabetes (NDM), maturity-onset diabetes of the young (MODY) and mitochondrial diabetes, are caused by a variant in a single gene in a given individual, and there are now some 40 different subtypes^3,4^. Improved insight into the mechanism of disease has been important to enable precision diabetes treatment for several of these disorders, e.g., sulfonylurea agents for the treatment of K_ATP_ neonatal diabetes^5,6^, HNF4A-MODY and HNF1A-MODY^7–9^. This diagnosis informs precision prognostics e.g., lack of microvascular or macrovascular complications in GCK-MODY and informs precision monitoring particularly in syndromic forms where the genetic diagnosis precedes the development of additional clinical features such as hepatic dysfunction and skeletal dysplasia in *EIF2AK3* or hearing and vision loss in *WFS1*^10,11^. Thus, diagnosing monogenic diabetes presents an opportunity to identify those who would benefit from precision medicine.

There are, however, key knowledge gaps that are obstacles for precision diagnostics in monogenic diabetes. The clinical diagnosis of diabetes is based on the measurement of a single molecule, glucose. The correct classification of diabetes relies on differentiating based on overlapping clinical features such as age, body mass index (BMI), history of diabetic keotacidosis, glycemic response to non-insulin therapies and the selective use of C-peptide and autoantibodies^12^. These features are even less reliable for correct diabetes classification in people of non-European ancestry, in whom the prevalence of type 2 diabetes is usually greater and often occurs from a younger age than in Europeans. The classical criteria for MODY, autosomal dominant inheritance pattern, onset of diabetes before 25 years, and residual beta cell function^13^, are not specific as they overlap with the clinical features seen in type 1 and type 2 diabetes^14^, particularly since markers of beta cell function are difficult to define and not routinely measured in all individuals with diabetes. These classical criteria are also not completely sensitive, since there are spontaneous mutations occurring in individuals without family history, autosomal recessive cases^15–17^, and later onset MODY cases. The term MODY originates from the time when the terms juvenile-onset and maturity-onset were used to distinguish between type 1 and type 2 diabetes and does not distinguish the various phenotypes associated with the numerous genetic etiologies for monogenic diabetes subtypes ^18^. Recent studies show that people with monogenic diabetes are often misdiagnosed as type 1 diabetes or type 2 diabetes^19^. Given the currently prohibitive cost and low yield of universal genetic testing in the vast majority with clinically classified type 1 and type 2 diabetes^14,20–22^, there is therefore a need for more knowledge on who to test for monogenic diabetes using various clinical and biomarker based criteria that increase the yield for this diagnosis, thereby, making such genetic testing more cost-effective.

Recent breakthroughs in sequencing technologies makes it possible to sequence the genome in a patient in less than a day^23,24^. Genome sequencing may not be appropriate for diagnosing monogenic diabetes due to costs, interpretation challenges, and ethical issues in reporting of incidental findings^25^. Less resource-demanding technologies are exome sequencing, panel exome sequencing and next-generation sequencing (NGS) using a targeted panel where many or all monogenic diabetes genes can be investigated simultaneously^26^. In some instances, like diagnosing a known disease-causing variant in additional family members, traditional Sanger sequencing might be preferred due to economy, speed, and reliability. The use of real-time PCR such as for detecting and quantifying mitochondrial m.3243A>G variant load, droplet digital PCR for analysis of both paternally and maternally inherited fetal alleles, copy number variant analysis for detecting gene deletions and methylation sensitive assays (e.g., for 6q24 abnormalities as a common cause of transient neonatal diabetes) are all available technologies. Thus, there are knowledge gaps regarding the choice of technology being a balance between cost, time, the degree of technical, scientific and bioinformatic expertise required, and the performance/diagnostic yield in particular diagnostic settings.

Best practices have been developed on how to report genetic findings^27^. The results of genetic tests may, however, be challenging to interpret^28^. Identifying a pathogenic variant may confirm a diagnosis of monogenic diabetes, indicate that a person is a carrier of a particular genetic variant, or identify an increased risk of developing diabetes. Although a “no pathogenic variant identified” test result does not confirm this, it is quite possible for a person who lacks a known pathogenic variant to have or be at risk for monogenic or other types of diabetes–sometimes because of limitations in technology but often due to inability to anticipate all possible genes that might be involved and limitations in our ability to interpret them depending on the technology used. In some cases, a test result might not give any useful information being uninformative, indeterminate, or inconclusive. If a genetic test finds a (VUS), it means there is not enough scientific research to confirm or refute causality of monogenic diabetes, or data are conflicting^29^. Two expert panels have formed to develop guidelines for reviewing evidence to determine which genes (ClinGen Monogenic Diabetes Gene Curation Expert Panel [MDEP GCEP, https://clingen.info/affiliation/40016/]) and gene variants (MDEP VCEP, https://clinicalgenome.org/affiliation/50016/) are considered causative of monogenic diabetes. But what is the evidence for these guidelines being used by the many diagnostics laboratories around the world?

The Precision Medicine in Diabetes Initiative (PMDI) was established in 2018 by the American Diabetes Association (ADA) in partnership with the European Association for the Study of Diabetes (EASD)^30^. The ADA/EASD PMDI includes global thought leaders in precision diabetes medicine who are working to address the burgeoning need for better diabetes prevention and care through precision medicine^31^. This systematic review is written on behalf of the ADA/EASD PMDI as part of a comprehensive evidence evaluation in support of the 2^nd^ International Consensus Report on Precision Diabetes Medicine^32^.

To investigate the evidence for who to test for monogenic diabetes, how to test them and how to interpret a gene variant, we set out to systematically review the yield of monogenic diabetes using different criteria to select people with diabetes for genetic testing and the technologies used. In addition, we sought to evaluate current guidelines for genetic testing for monogenic diabetes using a systematic review and grading of the studies available. The aim for this review was to fill the knowledge gaps indicated to improve diagnostics of monogenic diabetes and hence enhance the opportunity to identify those who would benefit from precision diagnostics. The evidence underpinning the link between the genetic test result and clinical management and prognostics are covered as separate systematic reviews in this series, by other members of the Precision Medicine in Diabetes Initiative (PMDI) addressing precision treatment and prognostics for monogenic diabetes.

## Methods

### Transparency and Openness Promotion Statement

The authors declare that all supporting data are available within this article and its supplemental material.

### Registration

We have registered a PROSPERO (International Prospective register of Systematic Reviews) protocol (ID:CRD42021243448) at link https://www.crd.york.ac.uk/prospero/. We followed the preferred reporting items for systematic reviews and meta-analysis guidelines^33^.

### Search strategy

We focused on seven questions for our review. For the questions of whom to test for monogenic diabetes, and which technologies should be used to test them, we searched PubMed (National Library of Medicine) and Embase.com using relevant keywords and thesaurus terms for relevant monogenic diabetes subtypes such as MODY, neonatal diabetes, lipodystrophy, combined with key genes of interest (**Supplementary Table 1**). Publication date limitation was set to 1990-2022, human studies only and English as a language limitation. A first search was performed in October 2021 with an update in June 2022. For the remaining questions our search strategies were adapted to recognize guidelines already in place for these areas. Details of our PICOTS framework is provided in **Supplementary Table 2**.

### Screening

For all questions except those relating to current guidelines, we carried out screening of papers using COVIDENCE (www.covidence.org). At least two reviewers independently screened titles and abstracts of all publications identified in the searches, blinded to each other’s decisions. Conflicts were resolved by two further reviewers. All remaining articles were retrieved and screened by at least two reviewers for eligibility, recording any reasons for exclusion. Disagreements were resolved by a third reviewer.

### Inclusion/Exclusion Criteria

For the question of whom to test for monogenic diabetes we included original research of any study design (cohort, case-control) but not case reports, which studied the pediatric or adult population with diabetes or mild hyperglycemia in whom the yield of monogenic diabetes was provided. A minimum of 100 unrelated probands with genetic testing results using sequencing of at least one or more genes implicated in monogenic diabetes had to be provided. Studies that only tested selected variant(s) within a gene or provided association of common variants in monogenic diabetes genes with type 2 diabetes risk were excluded. Reviews, commentaries, editorials, and conference abstracts were excluded. Other reasons for exclusion were if studies only involved animal models or *in vitro* data. Studies which did not provide any diabetes screening measurements or those where the outcome was not a subtype of monogenic diabetes or those focusing on treatment response or prognosis were excluded.

For the question of which technologies should be used to test for monogenic diabetes we included original research of any study design where a genetic testing methodology was employed to diagnose monogenic diabetes in a neonatal, pediatric, or adult population with diabetes, or where an evaluation of a genetic testing method had been undertaken. This included mitochondrial diabetes due to the m.3243A>G variant since this has recently been shown to be a common cause of diabetes in patients referred for MODY genetic testing^34^. We excluded studies using outdated or obsolete methods very rarely used by diagnostic laboratories. Functional studies on variants, studies detecting risk variants for polygenic forms of diabetes and linkage studies to identify candidate diabetes genes were excluded. The study had to provide a clear description of the methodology used, and studies were excluded where insufficient detail was provided.

#### Data extraction

From each included publication, we extracted data on the first author, publication year, and the following data:

1. Type of study, country, number of individuals genetically tested. For the question of who to test we also recorded their ancestry or country of the study, proportion female to male, BMI, other characteristics of those who were tested such as age of diabetes diagnosis, or other clinical or biomarker criteria. Where available, the extracted data also included measures of diagnostic test accuracy including sensitivity, specificity, receiver operating characteristic curve, and the area under the curve for discriminating between those with monogenic diabetes and those with other etiologies of diabetes.
2. Genetic testing methodology and number of genes tested gene variant curation method.
3. Number of individuals diagnosed with different monogenic diabetes subtypes, yield by different selection approaches if applicable.

#### Data synthesis

For the question of whom to test for monogenic diabetes, we summarized the total number of monogenic diabetes studies concerning neonatal diabetes, gestational diabetes, and other atypical presentations of diabetes. For each of these presentations of diabetes we group them according to whether they were tested for a single gene, small (2-5 genes) or a large gene panel ≤ 6 genes. We also summarized the studies where possible by whether they included international cohorts or those that includes individuals of predominantly European ancestry or non-European ancestry.

#### Critical appraisal and grading the certainty of evidence

A ten-item checklist for diagnostic test accuracy studies^35^ was used to assess the methodological quality of each study by two critical appraisers, and any conflicts were resolved by a third reviewer for Questions 1 and 2. This tool is designed to evaluate the risk of bias relating to diagnostic accuracy studies using three items regarding patient selection and seven items regarding the index test. Patient selection items included whether there was a consecutive or random sample of patients enrolled (Item 1). This was interpreted as yes if the cohort described consecutive enrolment from any given collection of individuals. For items 4-8, the index test was defined as the clinical features or biomarkers used to select individuals for genetic testing. The genetic test was considered the reference test, of which the current reference standard was decided to be at least a six-gene panel, including the genes most commonly associated with the phenotype. This for neonatal diabetes phenotype was considered to include *ABCC8, KCNJ11, INS, GCK, EIF2AK3, PTF1A*, and for non-neonatal beta-cell monogenic diabetes was considered to include *GCK, HNF1A, HNF4A, HNF1B, ABCC8*, *KCNJ11, INS* and m.3243A>G. The reference standard genetic test for diabetes associated with a lipodystrophy phenotype was considered to include at least *PPARG* and *LMNA*. Item eight, regarding an appropriate interval between the index test and the reference test to ensure that the status of the individual could not have meaningfully changed, was deemed not applicable to monogenic diabetes as the genetic test result remains stable throughout the person’s lifetime, hence a total of 9 items of this checklist were scored for each paper. We then synthesized the data from tabulated summaries and assessed the certainty of evidence by using the GRADE approach^36^.

The GRADE approach for diagnostic tests and test strategies was applied to answer the clinical question of who with diabetes should be offered the reference genetic test if we could not afford to provide this to everyone. The aim of the test (i.e. the clinical features and/or biomarkers) was to perform a triage function for selecting those with diabetes who had a greater likelihood of having a monogenic diabetes etiology, which when correctly diagnosed would enhance their clinical management. In assigning levels of evidence to the included studies considering various triage tests, 5 criteria were used as per the Canadian guidelines for grading evidence for diabetes studies^36^. Firstly, independent interpretation of the triage test results, without knowledge of the diagnostic standard (reference genetic test result) which was item 4 of the bias tool. This was considered to always be the case, given that clinical features and laboratory biomarkers (triage tests) were assessed independently of the genetic testing and variant curation. Secondly, independent interpretation of the diagnostic standard (the reference genetic test result) without knowledge of the triage test result, which was item seven of the bias tool. Whilst gene variant curation often relies on knowledge of the clinical features and laboratory biomarkers, this criterion was not deemed sufficiently informative for decisions about grading the evidence for the question of whom to offer genetic testing for monogenic diabetes. Thirdly, selection of people suspected (but not known) to have the disorder was considered for the summary of the evidence and related to item two of the bias tool of avoiding a case-control design. Fourthly, reproducible description of the test and diagnostic standard was considered. Finally, at least 50 patients with and 50 patients without clinical suspicion of monogenic diabetes was a key criterion that was considered. This criterion was incorporated into the inclusion criteria for studies considered relevant for the question of whom to test, by having a minimum of 100 unrelated probands with genetic testing results. To derive the overall level of evidence to the published studies, all five criteria had to be present for level 1, four criteria for level 2, three criteria for level 3 and one or two criteria for level 4 evidence. We developed guideline recommendations for whom to test for monogenic diabetes by assigning grade A for those criteria that were supported by best evidence at level 1, grade B for those that were supported by best evidence at level 2, grade C for those that were supported by best evidence at level 3 and Grade D for those that were supported by level 4 or consensus.

Answering Questions 3 (On what basis is a gene considered a cause of monogenic diabetes), 4 (On what basis is a variant considered a cause of monogenic diabetes), and 5 (How should a gene variant causing monogenic diabetes be reported) are central to putting knowledge about monogenic diabetes etiology into practice. Currently, individual laboratories select the genes to include on NGS panels, interpret variants according to internal guidelines, and create reports based on internal procedures. Recognizing the need for clarity and consistency in these areas, several national and international guidelines have been developed and refined. It was recognized that several general resources exist for assessing whether a gene is implicated in a disease, including the crowd-sourced UKPanelApp^37^ and the ClinGen evidence-based Gene-Disease Validity framework^38^. It was also noted that the ClinGen MDEP GCEP (https://clinicalgenome.org/affiliation/40016/) has convened to apply the ClinGen evidence-based framework to monogenic diabetes. Therefore, a de novo systematic evidence review for this question was not considered necessary or useful for this document, but rather a description of these existing resources and how they can be accessed. Similar to question 3, for question 4, it was recognized that consensus guidelines for assessing the role of specific genetic variants in disease were issued jointly by the American College of Medical Genetics and Genomics (ACMG) and the Association for Molecular Pathology (AMP) in 2015^39^ and the Association for Clinical Genomic Science in 2020. The ACMG/AMP guidelines have been expanded and refined by ClinGen^39–43^, and the ClinGen MDEP VCEP (https://clinicalgenome.org/affiliation/50016/) has convened to develop gene-specific rules for applying the guidelines to monogenic diabetes. For reporting genetic testing results (Question 5), there are some general published consensus guidelines^39,44,45^, and a limited emerging literature reporting studies evaluating report utility^46^ that was deemed not sufficient for a systematic evidence review. In this document, these are summarized and recommendations specific to monogenic diabetes are proposed based on existing practice.

For our evaluation of the next steps after a diagnosis of monogenic diabetes (Question 6), we excluded articles that either did not answer the question or only included a cursory general mention of the value of genetic testing for management. We reviewed the remaining 36 publications, consisting of specific case studies, cohorts, and review articles. Twelve papers discussed MODY testing and/or treatment in adults and children. Seven articles described strategies for testing and/or management of MODY during pregnancy. Three articles focused on maternally inherited diabetes and deafness (MIDD), five centered on neonatal diabetes, and nine covered syndromic forms of monogenic diabetes, including Wolcott-Rallison, Alström, and Wolfram syndromes. We then reviewed the literature for additional published studies relating to the steps after monogenic diabetes diagnosis. Information from publications was combined with expert advice from genetic counselors and physicians who specialize in monogenic diabetes clinical care. This section includes recommendations for results disclosure, cascade testing and addressing non-medical issues that may arise, with a focus on MODY being the most common form of monogenic diabetes. We direct the reader to other systematic reviews in this series for prognostics and treatment recommendations. To evaluate the challenges for diagnostic testing for monogenic diabetes (Question 7) we screened 455 abstracts for challenges for the field of monogenic diabetes diagnosis of which 41 were screened as full text articles and 14 taken forward for full text extraction.

## Results

### Question 1: - Who to test for monogenic diabetes?

For the question of who to test for monogenic diabetes, a total of 12,896 records were retrieved. In Covidence, 2,430 duplicates were identified. We included 100 publications from 10,469 publications screened (**Supplementary Figure 1A**). The key data from each of the 100 studies were included in **Supplementary Table 3** and the 10-item checklist (**Figure 1A**) assessments for these papers were summarized in **Figure 1B**. The summary of evidence from the included studies is detailed in **Table 1**. We also provide a list of recommendations based on this evidence in **Display Box 1.**

**Figure 1:**
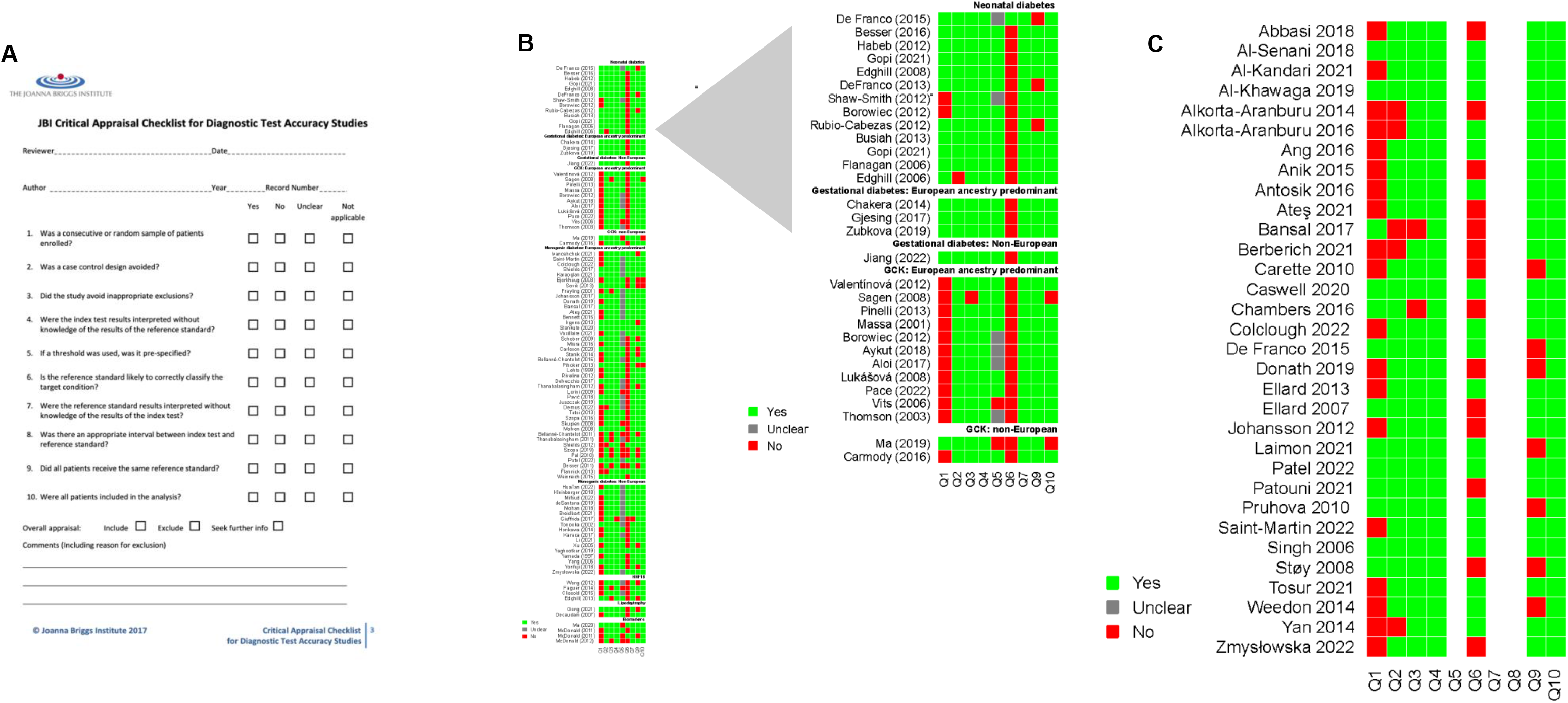
Critical appraisal of evidence using the Joanna Briggs Institute (JBI) Critical Appraisal tool for Systematic Reviews. **(A)** A ten-item checklist for diagnostic test accuracy studies was used to assess the methodological quality of each study. **(B)** Results for papers from question 1. **(C)** Results for papers from question 2. Green is Yes, Red is No, Grey is unclear. Non-applicable answers were left blank. All papers in (B-C) can be found in Supplementary Tables 3 and 4.

**Figure 2.**
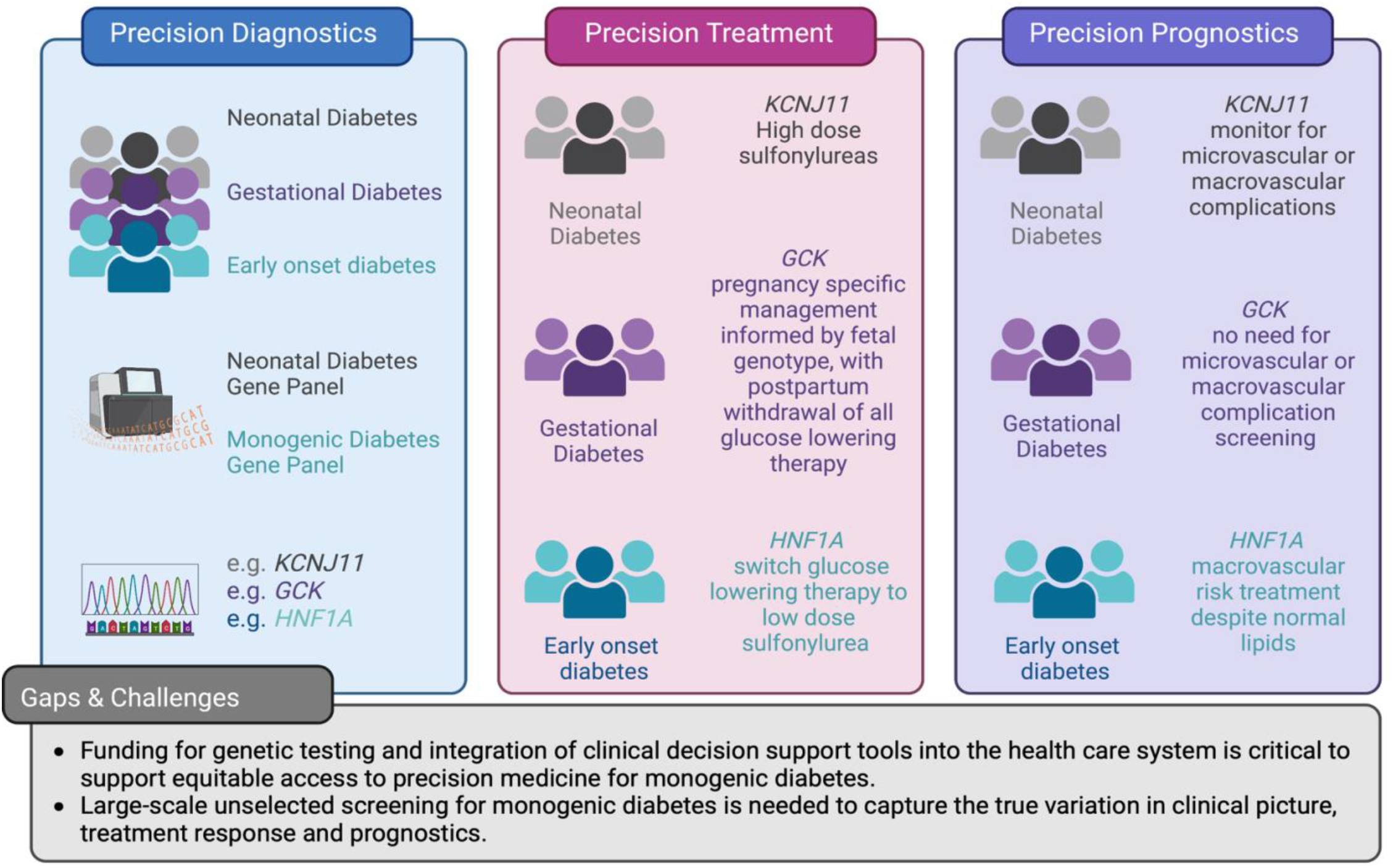
Schematic overview of how precision diagnostics leads to precision treatment and precision prognostics. Examples of genetic forms of diabetes identified through precision diagnostics and how these lead to precision treatment and prognostics. Current gaps and challenges identified through the systematic review are highlighted.

**Table 1:** Important summary of findings from sequencing studies for monogenic diabetes.

| Diabetes population studied<br>(country/ ancestry of population) | Genetic testing methodology | Number of studies<br>(Range of sample size tested) | Yield by key characteristics of diabetes population tested | Grade of evidence |
| --- | --- | --- | --- | --- |
| Neonatal diabetes diagnosed < 6m<br>(International) | >5 genes<br>(23 gene panel) | 1 study<br>(n=1020) | **Neonatal diabetes diagnosed <6 months should be offered genetic testing using large-gene panel**<br>Yield in a large, unselected other than by age at diagnosis, international study: 840/1020 (82%) | A |
| Neonatal diabetes diagnosed < 6m<br>(UK, Saudi Arabia, India) | ≤5 genes including **KCNJ11, ABCC8** | 3 studies<br>(n=165-750) | **Neonatal diabetes diagnosed <6 months tested with small gene panel including KCNJ11/ABCC8 has high yield**<br>UK:598/750 (78%),<br>Saudi/UK: 56/88 (64%) Saudi, 32/77 (42%) British<br>India: 39/181 (22%) | A |
| Neonatal diabetes diagnosed < 6m<br>(International cohorts) | Single genes: **INS**, or **SLC19A2** or after excluding **INS, ABCC8, KCNJ11** then **PDX1** or **GCK** | 4 studies<br>(n=103-212) | **Neonatal diabetes diagnosed <6 months tested for single genes with or without excluding more common gene etiologies have lower yields**<br>INS: 33/141 (23%)<br>SLC19A2: 3/212 (1%)<br>PDX1: 3/103 (3%)<br>GCK homozygous: 1/17 (6%) | A |
| Neonatal diabetes diagnosed <12m<br>(Spain, France, India) | ≤5 genes<br>Including **INS** or **KCNJ11** and **ABCC8** | 3 studies<br>(n=189-405) | **Neonatal diabetes diagnosed <12 months tested for common genes has a lower yield than for those diagnosed <6 months and there are less cases**<br>Spain: 263/405 (65%) diagnosed <6m, and 9/145 (6%) diagnosed 6-12m<br>France: 64/155 (41%), diagnosed <6m and 5/18 diagnosed 6-12m (28%)<br>India: INS only in PNDM, Ab-ve, CP+ve, diagnosed<9m: 8/189 (4%) | A |
| Neonatal diabetes diagnosed <24 months (UK, International samples) | **KCNJ11** only or **INS** after **KCNJ11** negative | 3 studies<br>(n=58-70) | **There were no cases of monogenic diabetes found in children diagnosed age 12-24 months although only limited genes were tested**<br>0/70 **KCNJ11**<br>0/63 **KCNJ11**<br>0/58 **INS** | B |
| Gestational Diabetes (GDM)<br>European cases | GCK only | 3 Studies<br>(n=188-400) | **In European women with GDM, yield for GCK-MODY was 1%-6% when otherwise unselected, rising to 31% when only women without obesity were selected**<br>~UK and Ireland: overall 4/356 (1%)<br>~Diet-treated Danish GDM: 21/354 (6%) mean BMI 28<br>~Non-obese Russian GDM: 59/188 (31%) | **A** |
| Gestational Diabetes (GDM)<br>China | GCK only | 1 Study<br>(n=411) | **There is a lack of studies in non-European individuals to define the best testing criteria for GCK-MODY in pregnancy**<br>In Chinese women with GDM the yield for GCK was 4% (15/411) when otherwise unselected | **C** |
| Children and adults with diabetes, not GDM.<br>Predominantly European ancestry | GCK only | 12 Studies<br>(n =100-722) | In European cases, the yield for GCK-MODY in studies with a high clinical suspicion e.g. those with persistent, stable, mild hyperglycemia or fasting hyperglycaemia is high ranging from 30-74%.<br>Adding other MODY criteria such as absence of obesity or family history of diabetes does not consistently increase the yield (27-88%). | **A** |
| Children and adults with diabetes, not GDM.<br>Predominantly non-European ancestry | GCK only | 3 studies<br>(n=24 and 679) | In a Chinese study with mild fasting hyperglycaemia (5.4-8.3) and low triglycerides, yield was much lower than in the European groups:<br>Discovery group 11/545 (2%) and replication groups 1/207 (0.5%)<br>In the US MODY registry, there were small numbers of non-European participants but the proportion of GCK-MODY was similar at 11/24 (46%) in a Turkish study | **B** |
| Children and adults with diabetes, not GDM<br>Predominantly European ancestry | Large monogenic diabetes panels (5-28 genes) | 16 studies<br>(n=178-6888) | **Yield in Europeans using large MD gene panel varies by selection criteria from 0.7% to 34%. A large study of older, unselected adults showed low yields.**<br>In suspected MODY cohorts yield was 16%-23% (4 studies)<br>In the Norwegian Childhood Registry, yield in Ab-ve children diagnosed <15y was 19/462(4.1%)<br>In Turkish children diagnosed <18 years atypical for T1D or T2D, yield was 24/330 (10%)<br>In those Ab-ve diagnosed<30y and either C-peptide positive or non-insulin treated, yield was 4% (51/1407) in a UK study, and higher at 21% (38/178) in additionally non-obese French individuals diagnosed <35 years | **A** |
|  |  |  | Comparative yield for 2-3 clinical criteria of diagnosed 15-40y, BMI<30, FH was 16-20% vs diagnosed <40y and BMI<25 was 34% in 1 French study (n= 1564)<br>In a large, relatively unselected and older onset study of “non-T1D” there was a low yield diagnosed>40y at 0.7% (18/2670) and diagnosed<40y at 2% (29/1346) (1 German study) |  |
| Children and adults with diabetes, not GDM<br>Predominantly Non-European ancestry | Large MD gene panel<br>(>5 genes) or sequential targeted exome/ whole exome sequencing | 6 studies<br>n=184-488 | **Yield in mixed ethnicity cohorts using large MD gene panel was similar to that seen in the European cohorts at 13%-26% using various selection criteria:**<br>Turkish children diagnosed 5-12y, either low T1D GRS or mod T1D GRS with Ab-ve had yield 34/236 (14%), 14 of the 34 were autosomal recessive diseases (~20% consanguinity)<br>USA/India cohort diagnosed<30y, Ab-ve, FH: 39/152 (26%)<br>France cohort Ab-ve and 2 or more of diagnosed<40y, BMI<30 at diagnosis, FH: 315/1975 (16%)<br>Suspected MODY cohorts 13% and 15% in Brazil and Singapore | **A** |
| Children and adults with diabetes, not GDM<br>Mixed ethnicity | Small MODY panels or 3-5 individual genes | 7 studies<br>n=100-4010 | **Yield from testing 3-5 common MD genes varies widely by age and selection criteria (from 8-97%):**<br>Suspected MODY (4 studies) ranged from 13% in South Asians, 29% in other UK cohort, 40% in French, 57% in Slovakia/Czech, to 97% in German study<br>In those diagnosed 1-18 years, 4 comparative selection criteria yields ranged from 0% (0/182) in Ab+ve, vs 15% (46/303) in Ab-ve, vs 30% (29/96) with additional FH vs 34% (44/131) with additional FH or HbA1c<7.5% at diagnosis<br>The USA SEARCH cohort with a high level of obesity and non-white ethnicity, diagnosed <20 years, Ab-ve, C-pep +ve, had yield 8% | **B** |

**Table 2:**
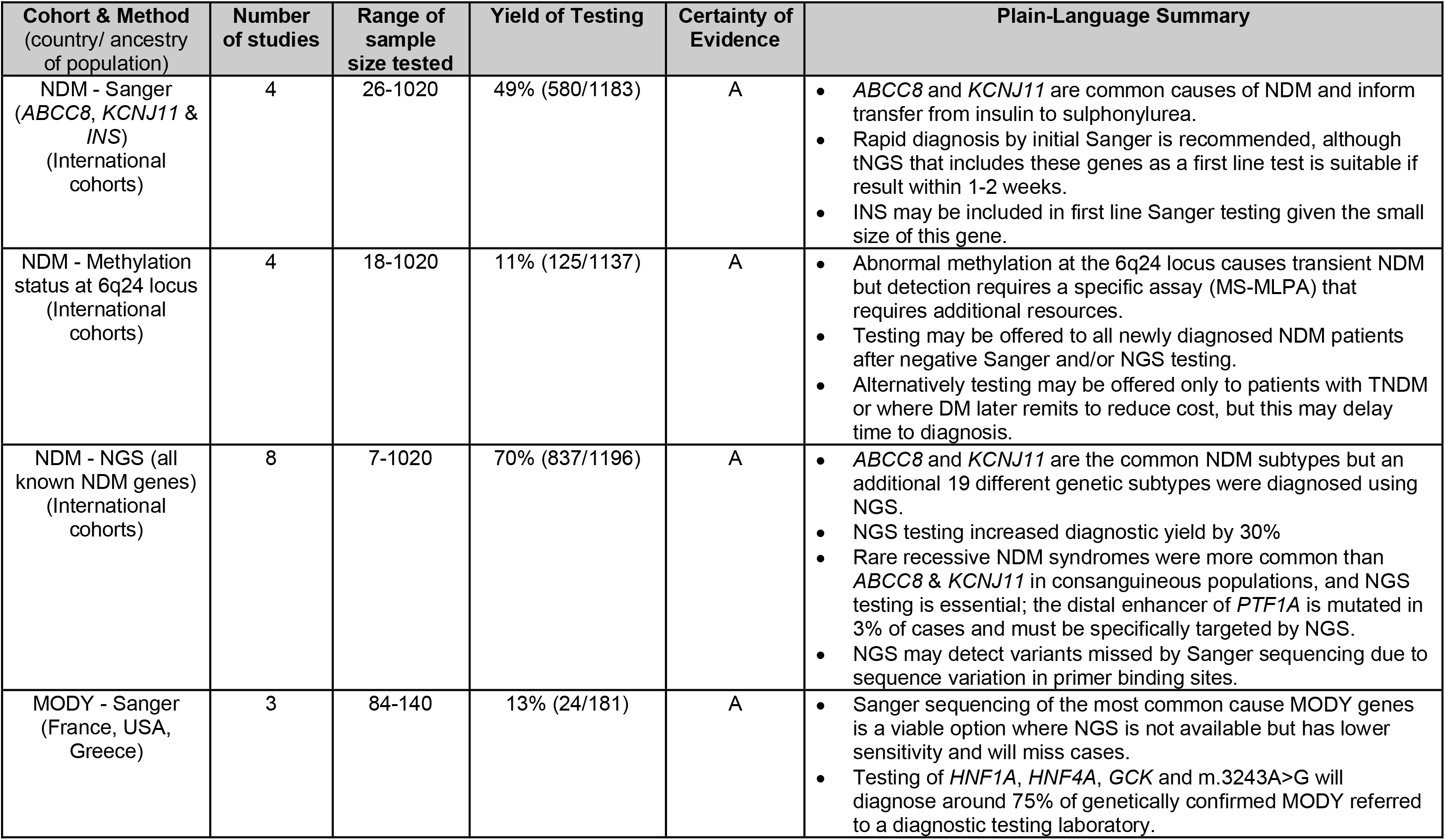
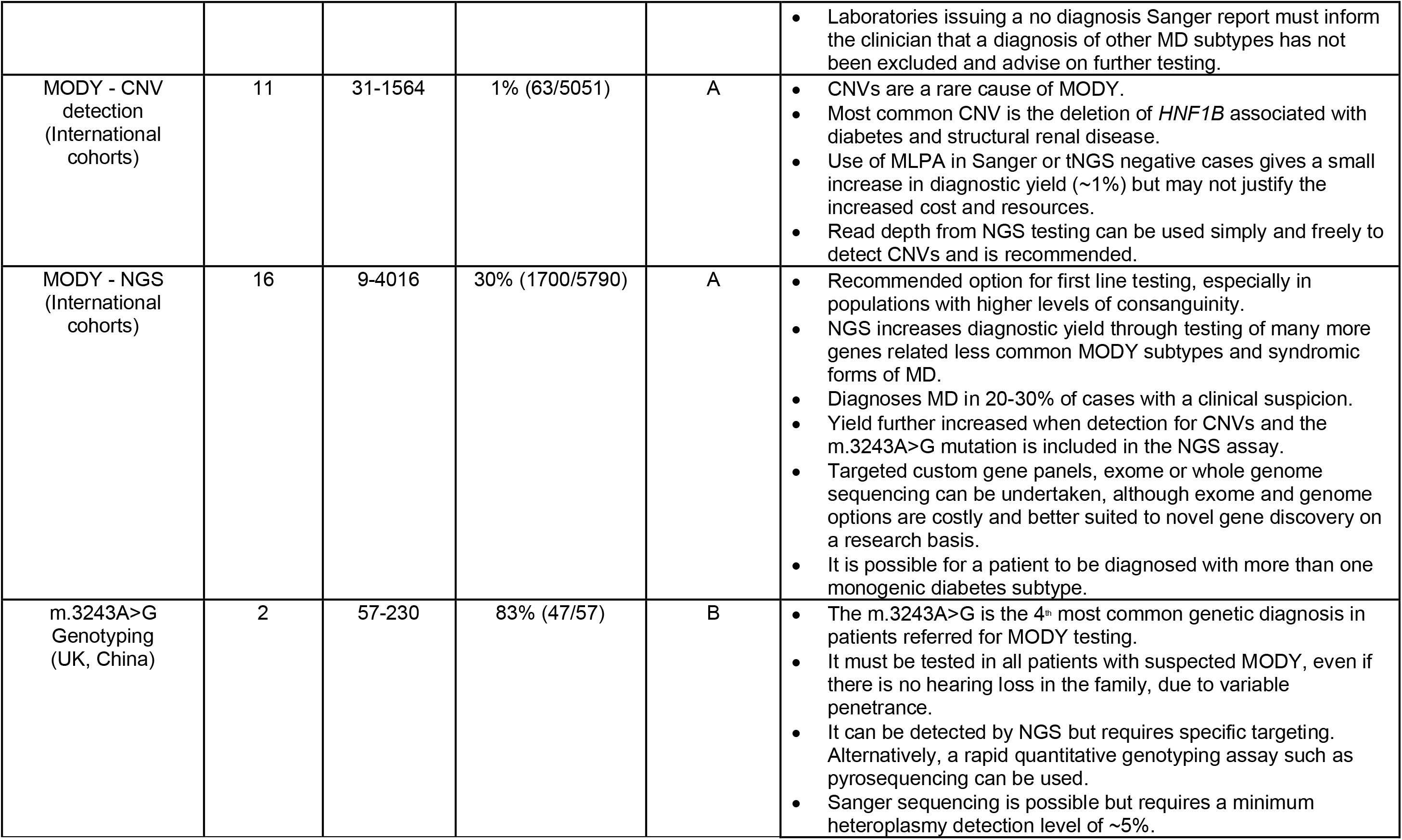
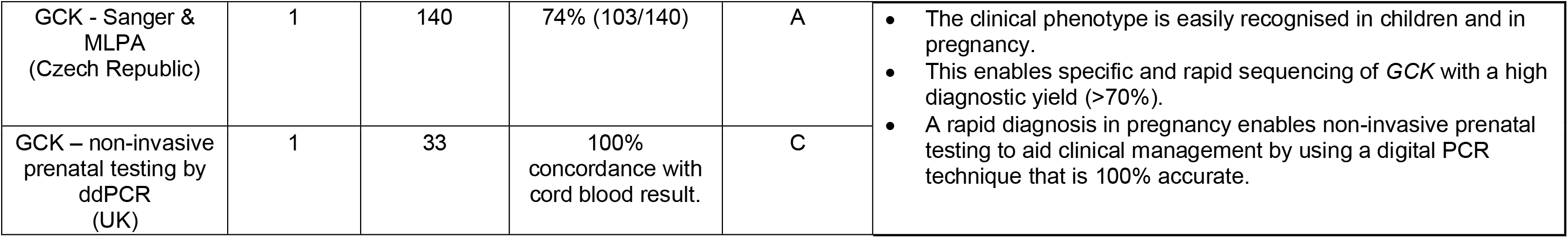
Summary of findings for testing platforms for monogenic diabetes.

**Box 1:**
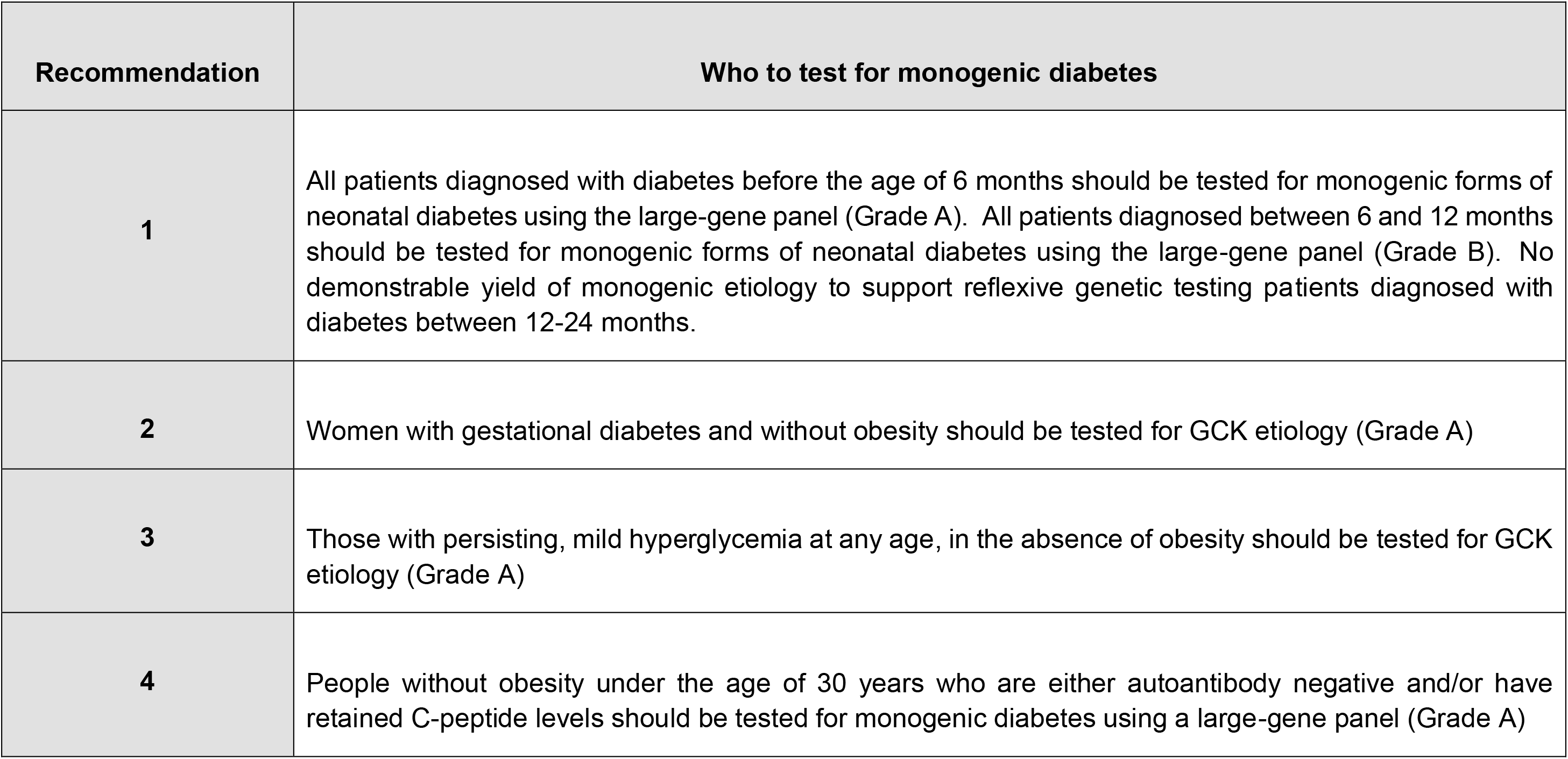
Recommendations based on the synthesis of evidence for who to test for monogenic diabetes.

In neonatal diabetes there were a total of 14 studies, of which three included those diagnosed with diabetes within 24 months of age, three within 12 months of age and the rest within six months of age **(Table 1**). There was only one study which used the reference standard large gene panel for neonatal monogenic diabetes diagnosis, while the rest did not. The highest yield of 82% was obtained in a single international cohort study of 1,020 patients diagnosed with diabetes within six months of age using a large 23-gene panel^47^. Of these, 46% had *KCNJ11* or *ABCC8* followed by *INS* as the next common etiology.

For neonatal diabetes diagnosed between 6-12 months the yield was 0-28% derived from six studies containing sample sizes of 18 to 145 individuals tested using only *KCNJ11, ABCC8, INS genes.* No cases of monogenic diabetes were found in the small subpopulations tested with diabetes diagnosed 12-24 months in three studies sequencing *KCNJ11 and INS* genes only (n=58-70). The risk of bias criterion for patient selection was high for two studies because a case-control study had not been avoided for one^48^ and a consecutive or random sample of patients had not been enrolled in another^49^. Two studies were deemed to be at risk of bias due to not all receiving the reference test^50,51^.

Applying the GRADE approach to the question of who to test for neonatal diabetes due to the patient-important outcomes of precision treatment and prognostics was considered greatest for *KCNJ11/ABCC8*, and genes associated with syndromic conditions. The selection by age of diabetes diagnosis below six months for neonatal diabetes genetic testing was supported by level 1 evidence from 1 study and thereby supports this being a Grade A recommendation. Selection by age of diabetes diagnosis below 12 months was supported by a yield of up to 28% by level 2 evidence from six studies, although these were limited by only testing for *INS* or *KCNJ11* and *ABCC8*. Selection by age of diabetes diagnosis beyond 12 months for monogenic diabetes testing was not supported by three studies examining those diagnosed with diabetes up to 24 months. These failed to find any cases of monogenic diabetes although these were limited by only testing for *KCNJ11* or *INS* etiologies in small cohorts with diabetes diagnosed between 12-24 months (level 2 evidence).

#### Recommendation 1

All patients diagnosed with diabetes before the age of 6 months should be tested for monogenic forms of neonatal diabetes using the large-gene panel (Grade A). All patients diagnosed between 6 and 12 months should be tested for monogenic forms of neonatal diabetes using the large-gene panel (Grade B). No demonstrable yield of monogenic etiology to support reflexive genetic testing patients diagnosed with diabetes between 12-24 months.

In gestational diabetes mellitus (GDM), there were a total of four studies which examined *GCK* diagnosis only, of which three were in predominantly European women^52–54^, and one study was in Chinese women^55^. The yield for *GCK* etiology ranged from 1%-6% in otherwise unselected women with GDM, however, increased to 31% when only non-obese women were selected for *GCK* testing^54^. The Russian study of non-obese women with GDM had a yield of 59/188 (31%) including over 50 women with *GCK* who had been suspected but not known to have the disorder in the study, thereby fulfilling level 1 evidence. Other than only testing for the single gene *GCK*, there were no other concerns about bias in these studies.

#### Recommendation 2

Women with GDM and without obesity should be tested for *GCK* (Grade A).

For *GCK* testing in those without GDM, there were a total of 14 studies of which 12 were in predominantly European populations. Overall, there was frequent assessment of bias in patient selection criteria used in all but one study. There were 5 studies with either unclear or no thresholds provided to define the triage tests which were most commonly persistent, stable, mild hyperglycemia. The yield for *GCK* etiology ranged from 0% in unselected cases of hyperglycaemia and increased to 30-74% in those with persistent, stable, mild hyperglycemia (**Table 1**). There was only one Italian study of 100 individuals that compared two testing strategies^56^. This study demonstrated that the yield for *GCK* increased in those with impaired fasting glucose and without diabetes autoantibodies from 32% when one MODY criteria was added compared to 88% when non-obese and lack of diabetes medications was added. However, this study characteristics provided level 3 evidence. The yield for *GCK* in a Chinese study which used mild, fasting hyperglycemia and low triglycerides was relatively low (2% vs 0.5% in discovery and replication datasets of n=545 and n=207 respectively)^57^. However, in a mixed ethnicity population in the USA, selection of those with persistent, mild fasting hyperglycemia plus either family history or BMI below 30kg/m^2^ or diabetes diagnosis age below 30 years produced a yield of 55%^58^. Overall, four studies supported level 1 evidence for selecting those with persisting, mild, fasting hyperglycemia for *GCK* testing.

#### Recommendation 3

Those with persisting, mild, fasting hyperglycemia at any age, in the absence of obesity should be tested for GCK etiology (Grade A).

There were 60 studies which examined the yield of monogenic diabetes beyond the neonatal period, of which 43 were in predominantly European populations. Of these, 25 studies utilized the reference standard of the large-gene panel (8 in non-European populations). The yield varied by the triage test strategy utilized to select individuals for genetic testing and those receiving the large-gene panel had a greater yield than smaller or single gene approaches. Younger age of diagnosis of diabetes (thresholds included below 15, below 18, below 25, below 35 and below 40 years) and negative diabetes autoantibodies was the most common triage test strategy. Excluding those with type 1 diabetes using either negative diabetes autoantibodies or presence of C-peptide or both was frequently employed. With the large-gene panel approach, the yield for a monogenic etiology ranged from 0.7% to 34%. There was low yield of 18/2670 (0.7%) in those with negative antibodies who had diabetes diagnosed above the age of 40 years^59^. In suspected MODY cohorts, the yield was 16% to 23% (**Table 1**). Most of such studies were assessed as having bias in patient selection and many did not have a clear description of “suspected MODY” (**Figure 1B**). One French study of 1564 individuals provided the comparative yield for (a) 3 clinical criteria of diabetes diagnosis age of 15-40 years, BMI below 30kg/m^2^, and family history of diabetes which was 20% vs (b) for any 2 of these clinical criteria the yield was 16% vs (c) diabetes diagnosis of 15-40 years and BMI below 25kg/m^2^ the yield was 34%^60^. In a Turkish cohort of children with diabetes (diabetes diagnosis age IQR 5-12 years), with either low Type 1 diabetes genetic risk score (T1GRS) or moderate T1GRS and negative diabetes autoantibodies had a yield of 34/236 (14%). This included 14/34 autosomal recessive cases, with approximately 20% prevalence of consanguinity in the tested population^61^. While there was considerable heterogeneity in selection criteria used, the best evidence was at level 1 for selecting those diagnosed with diabetes below the age of 30 years who are either autoantibody negative/and or have retained C-peptide (for lowering probability of type 1 diabetes) and those without obesity (for lowering probability of type 2 diabetes) for testing for monogenic diabetes using the reference large-gene panel.

#### Recommendation 4

People without obesity under the age of 30 years who are either autoantibody negative and/or have retained C-peptide levels should be tested for monogenic diabetes using a large-gene panel. (Grade A evidence)

### Question 2: - How to test for monogenic diabetes?

For the question of which technologies should be used to test for monogenic diabetes, we included 32 studies from 2,102 publications screened **(Supplementary Figure 1B)**. A total of 32 studies which accessed 76 different genes were analyzed (**Supplementary Table 4, Table 3**) and assessed for methodological quality **(Figure 1C)**. NGS was the most used technique, with 16/22 NGS studies using a targeted panel. Where NGS was employed, the MODY diagnostic yield increased by around 30% compared to Sanger sequencing of *GCK*, *HNF1A* and *HNF4A* alone, and resulted in the (often unexpected) diagnosis of rare syndromic forms of diabetes, most commonly m.3243A>G. NGS technologies also enabled the diagnosis of multiple monogenic subtypes in the same patient, and diagnosed patients who were missed by previous Sanger sequencing due to allelic drop-out. Gene agnostic exome and genome strategies were rarely used and did not increase diagnostic yield. Copy-number variant (CNV) analysis (by Multiplex-Ligation-Dependent Probe Amplification [MLPA] or NGS) increased diagnostic yield mostly by detecting *HNF1B* deletions. Non-coding variants were rare but important findings and required genome sequencing or specific targeting of non-coding mutation loci. A high diagnostic yield (74%) was reported when performing Sanger sequencing of *GCK* in patients with a clinical suspicion of GCK-MODY. Similarly, variants in *KCNJ11*, *ABCC8* and *INS* accounted for 50% of neonatal diabetes mellitus (NDM) cases and were sequenced by Sanger first in some studies. 6q24 abnormalities were also a common cause of NDM and required a specific methylation-sensitive assay to detect them. Recessively inherited and syndromic forms of monogenic diabetes were predominant in countries with high rates of consanguinity. Real-time PCR and pyrosequencing were highly sensitive and specific techniques for detecting m.3243A>G and quantifying heteroplasmy, and ddPCR successfully determined all fetal genotypes in a cell-free fetal DNA prenatal testing study of 33 pregnancies.

**Table 3:**
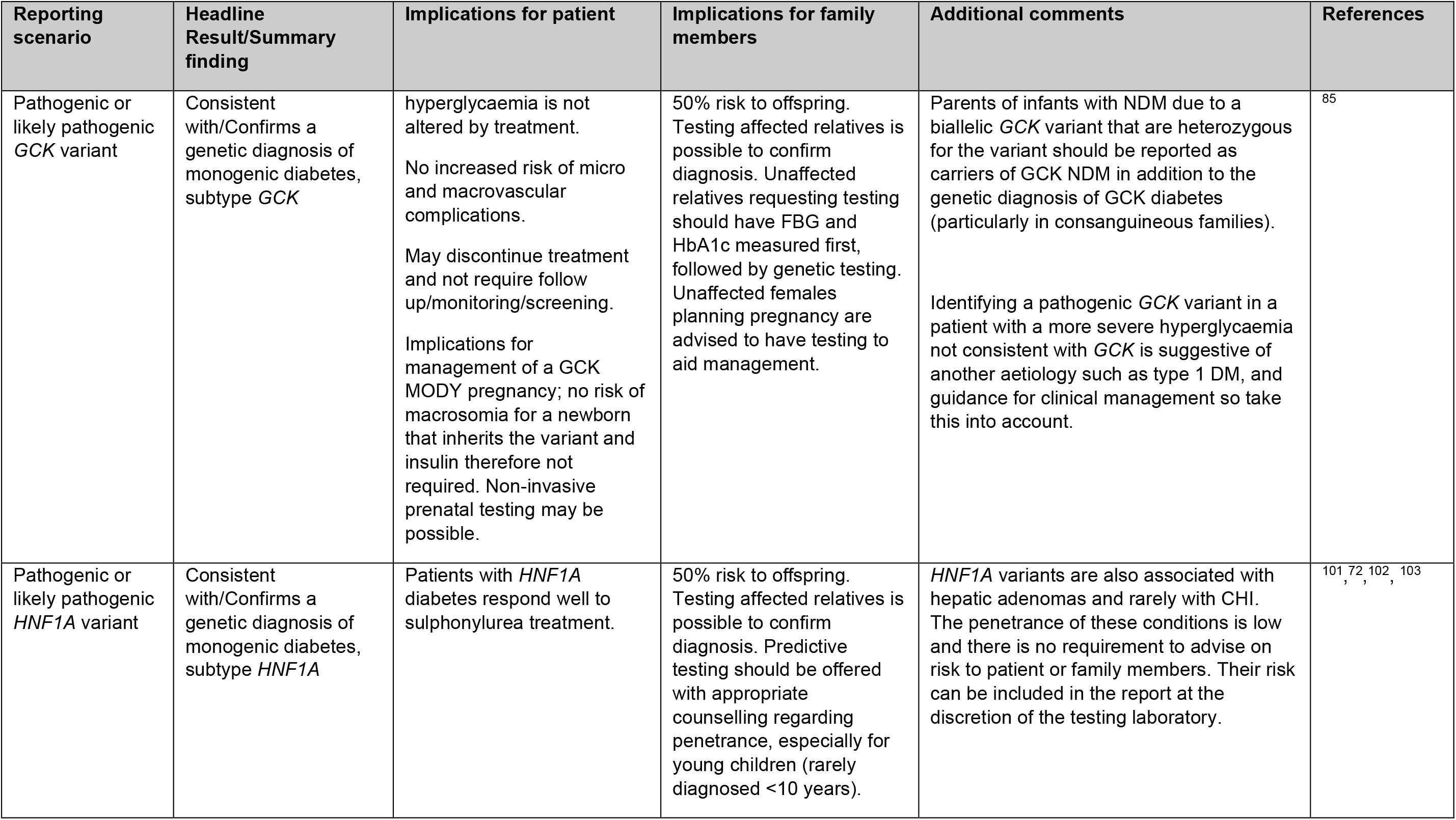
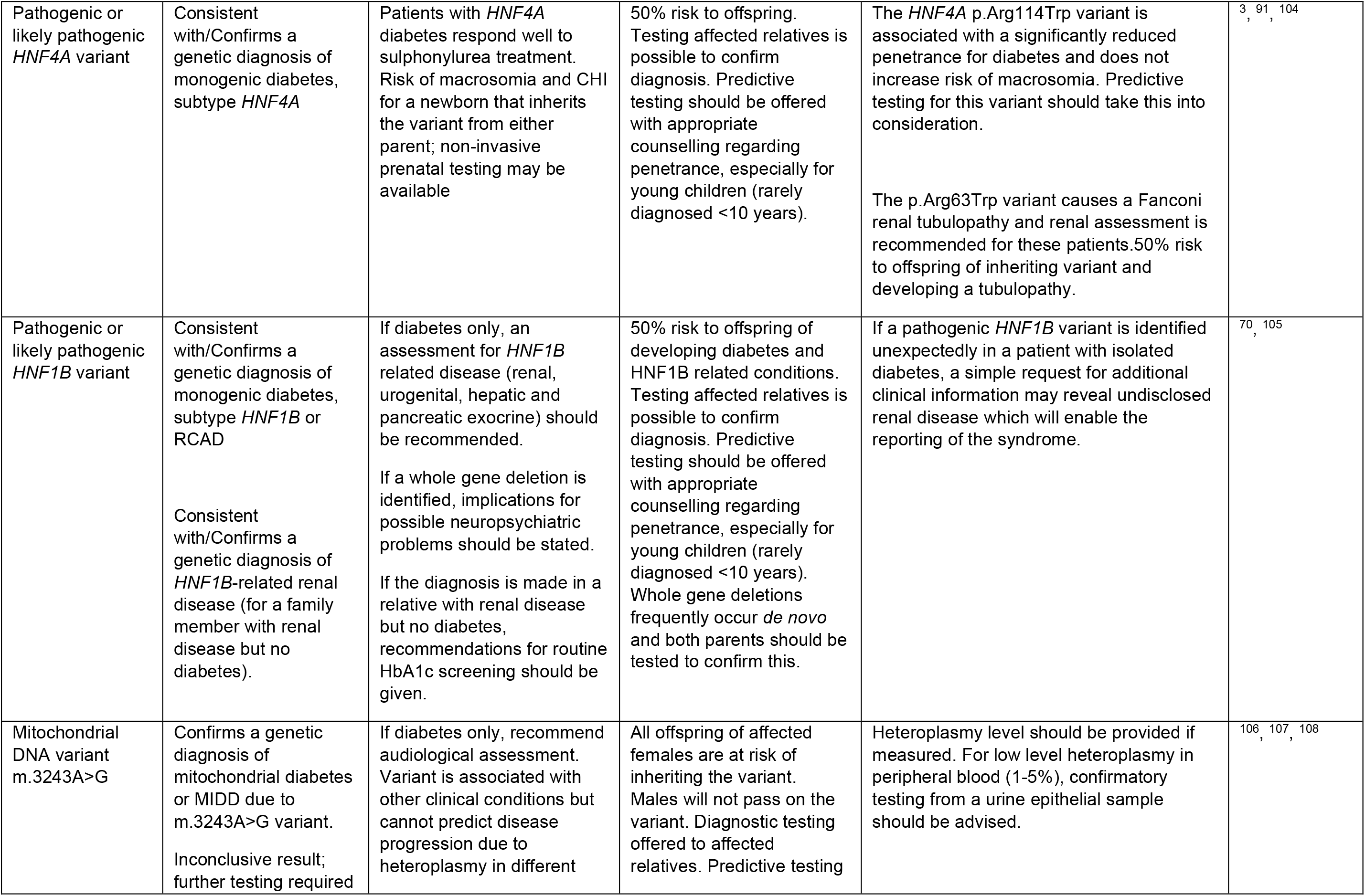
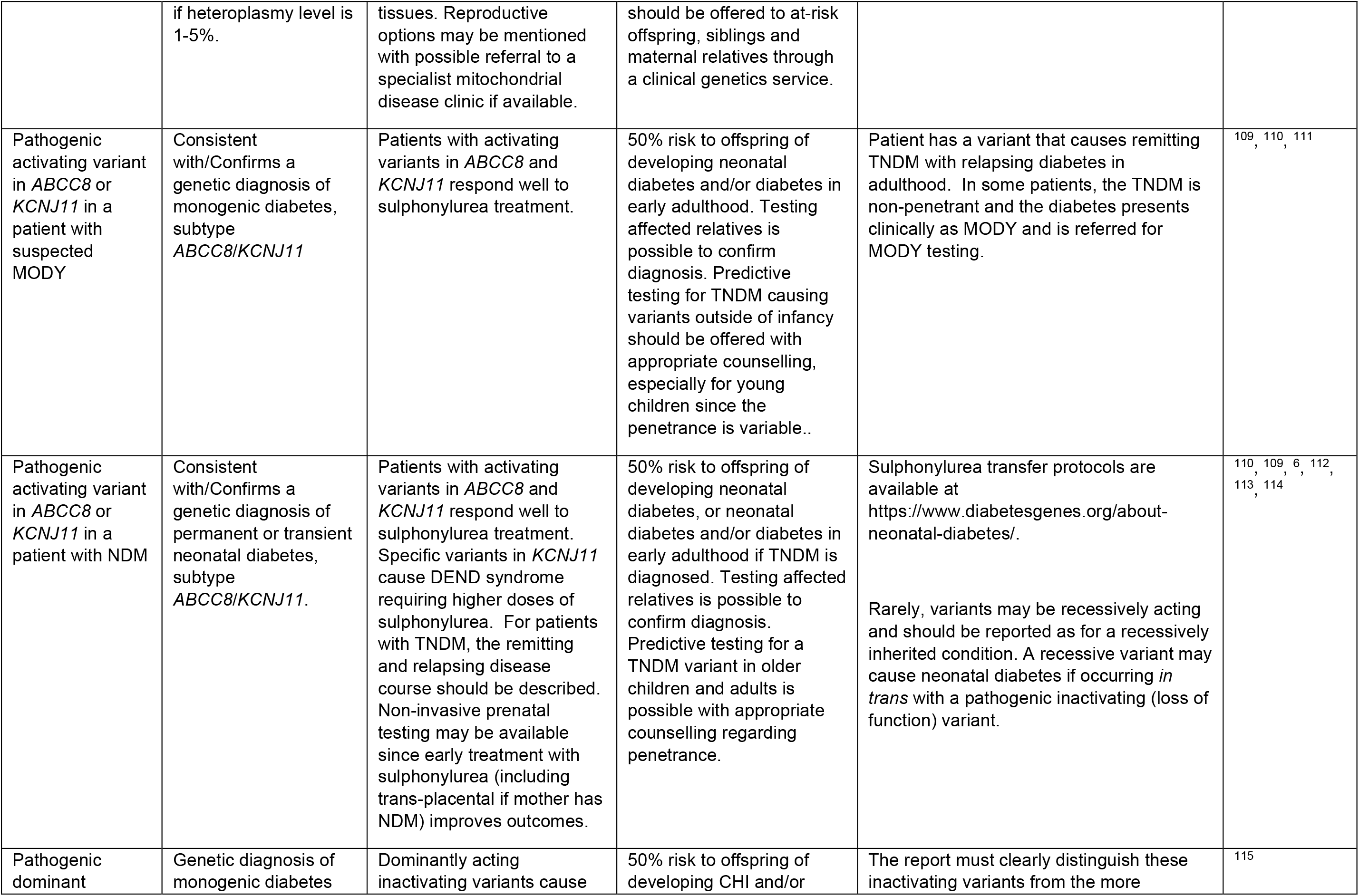
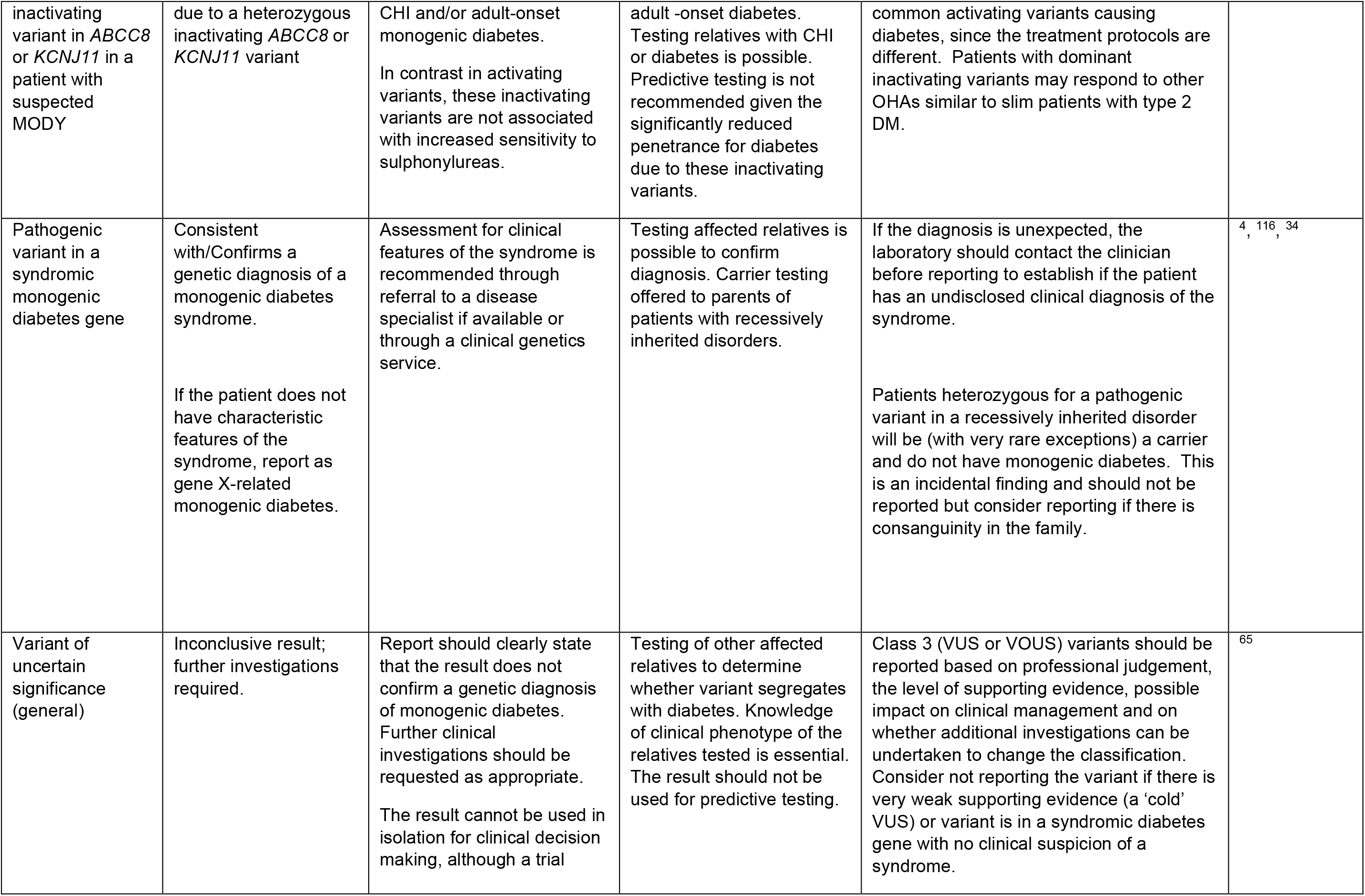
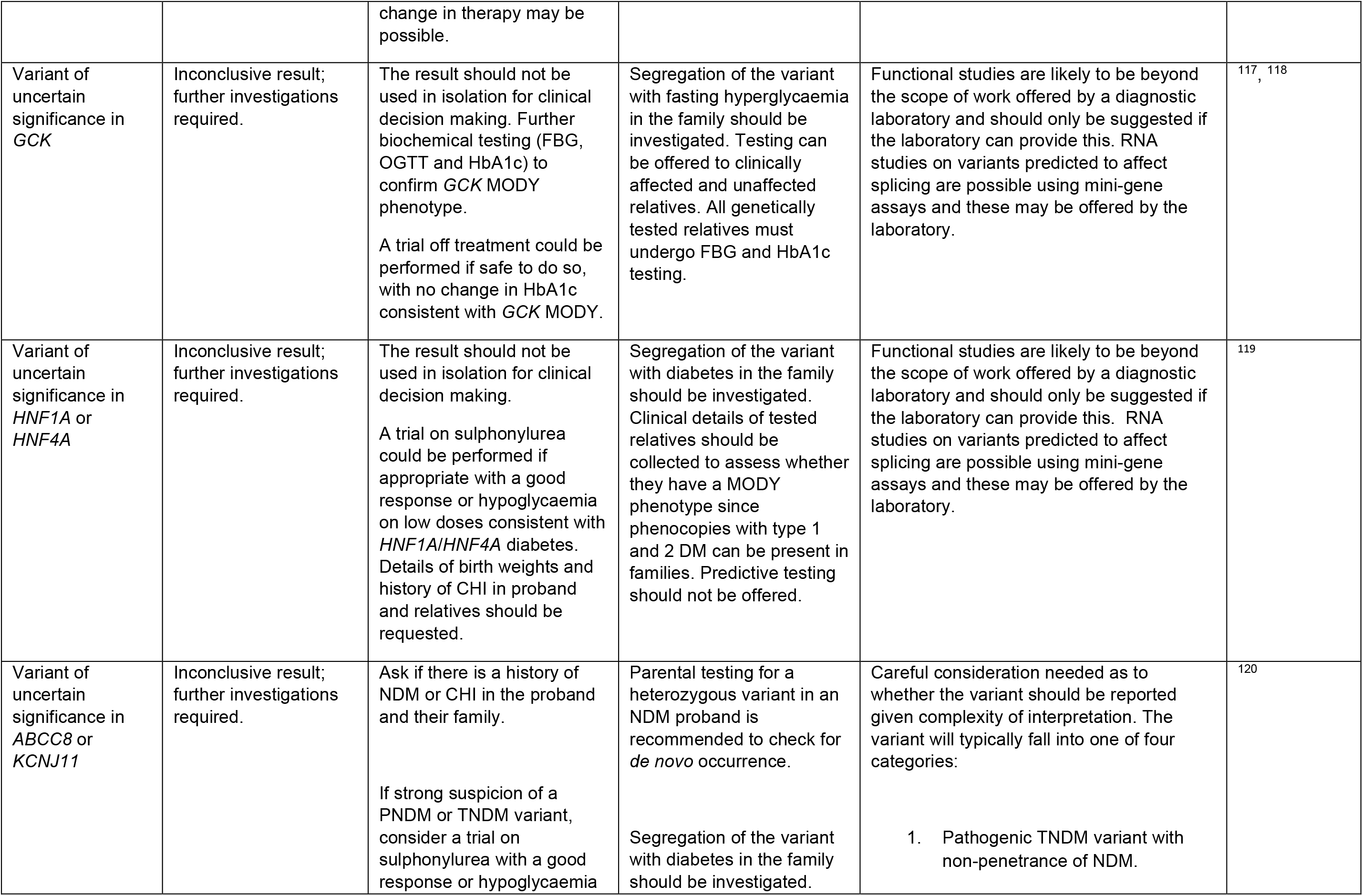
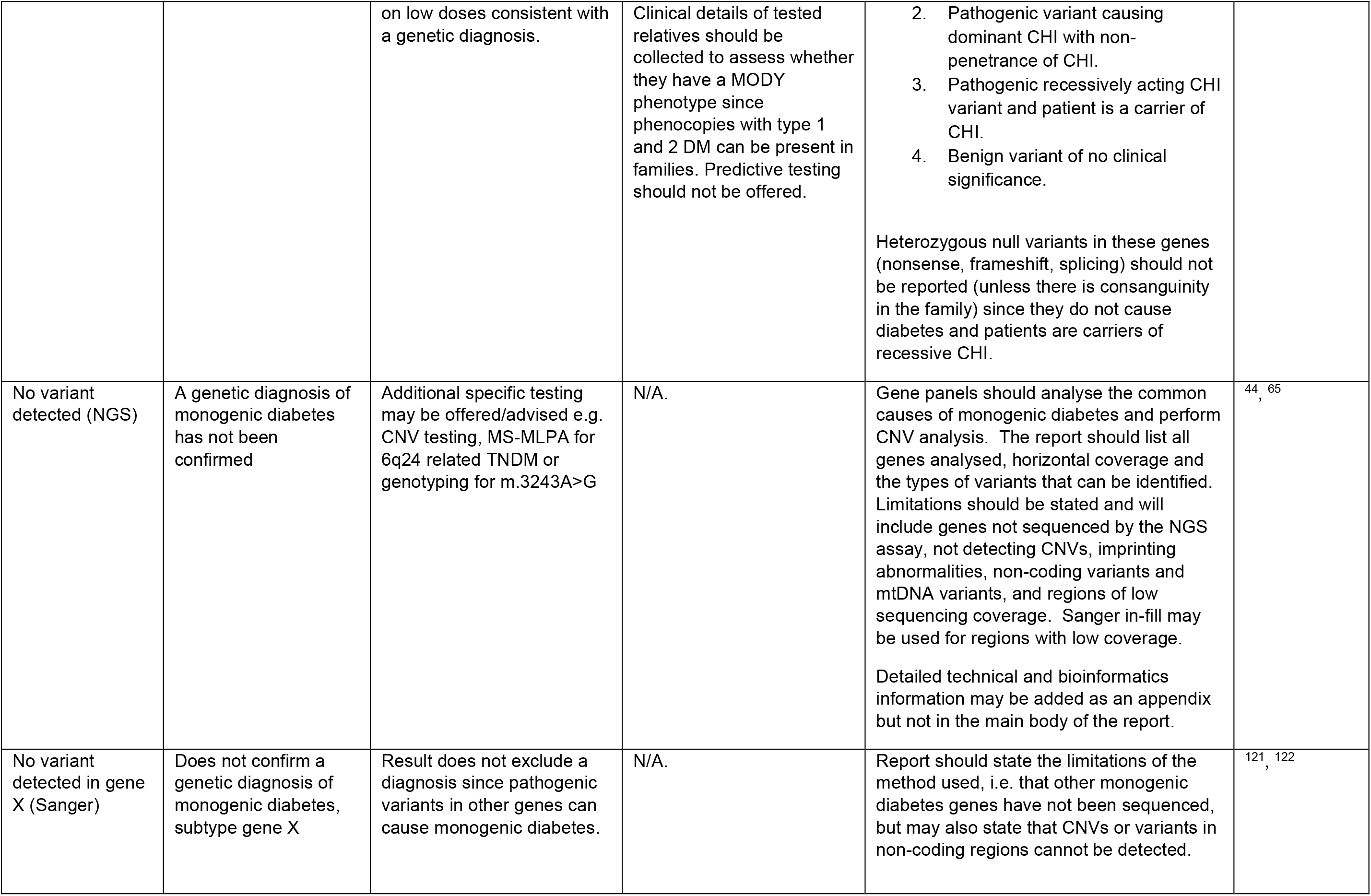
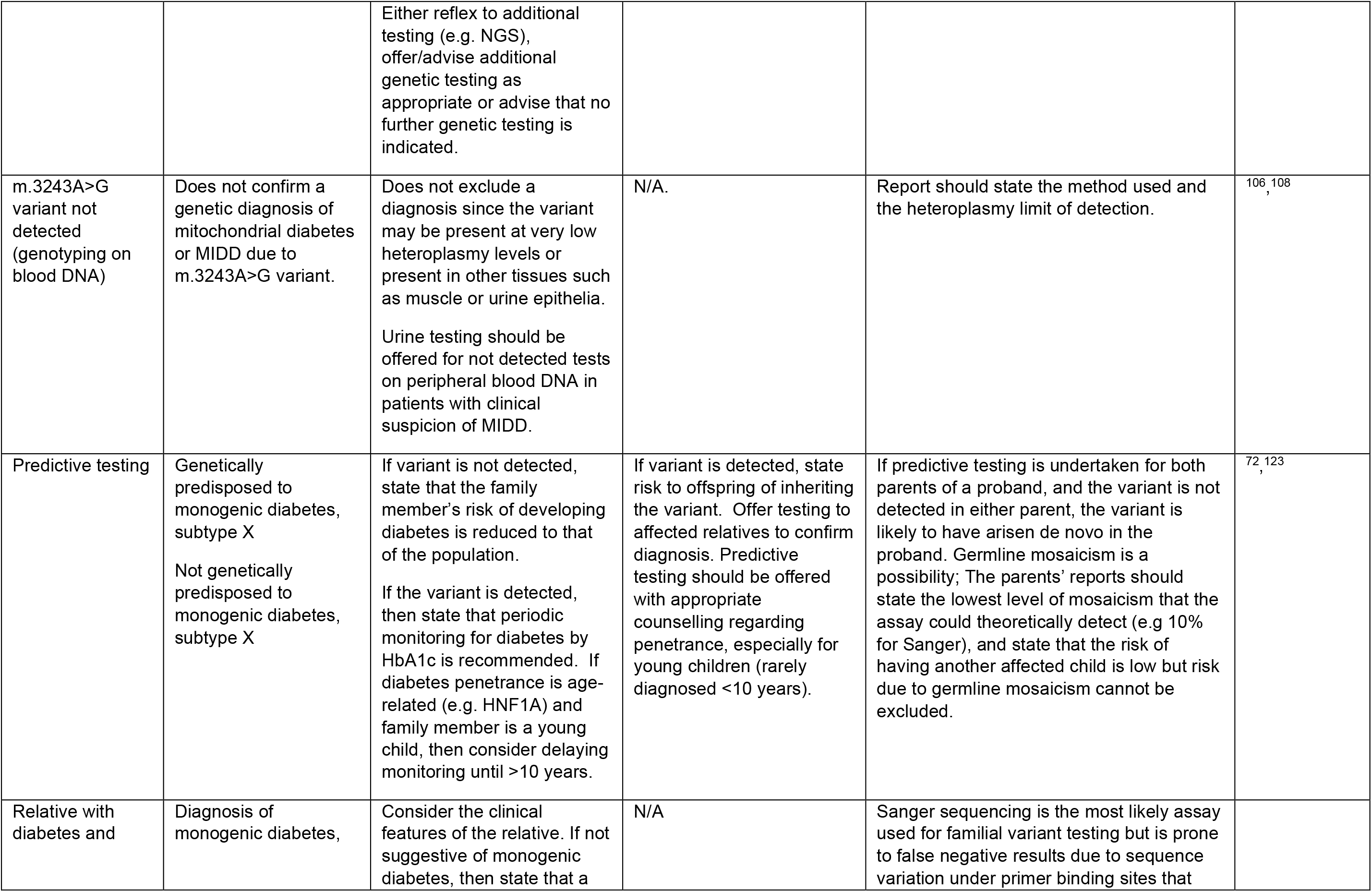
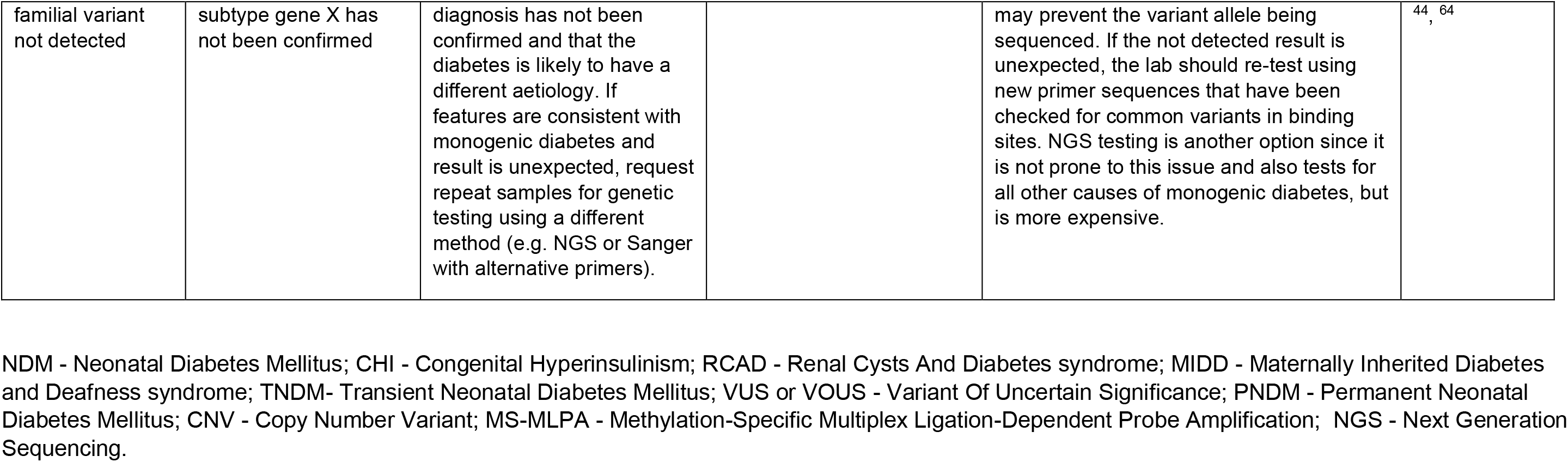
Recommendations for reporting results of monogenic diabetes testing across a range of different testing and reporting scenarios.

| Reporting scenario | Headline Result/Summary finding | Implications for patient | Implications for family members | Additional comments | References |
| --- | --- | --- | --- | --- | --- |
| Pathogenic or likely pathogenic *GCK* variant | Consistent with/Confirms a genetic diagnosis of monogenic diabetes, subtype *GCK* | hyperglycaemia is not altered by treatment.<br>No increased risk of micro and macrovascular complications.<br>May discontinue treatment and not require follow up/monitoring/screening.<br>Implications for management of a *GCK* MODY pregnancy; no risk of macrosomia for a newborn that inherits the variant and insulin therefore not required. Non-invasive prenatal testing may be possible. | 50% risk to offspring. Testing affected relatives is possible to confirm diagnosis. Unaffected relatives requesting testing should have FBG and HbA1c measured first, followed by genetic testing. Unaffected females planning pregnancy are advised to have testing to aid management. | Parents of infants with NDM due to a biallelic *GCK* variant that are heterozygous for the variant should be reported as carriers of *GCK* NDM in addition to the genetic diagnosis of *GCK* diabetes (particularly in consanguineous families).<br>Identifying a pathogenic *GCK* variant in a patient with a more severe hyperglycaemia not consistent with *GCK* is suggestive of another aetiology such as type 1 DM, and guidance for clinical management so take this into account. | 85 |
| Pathogenic or likely pathogenic *HNF1A* variant | Consistent with/Confirms a genetic diagnosis of monogenic diabetes, subtype *HNF1A* | Patients with *HNF1A* diabetes respond well to sulphonylurea treatment. | 50% risk to offspring. Testing affected relatives is possible to confirm diagnosis. Predictive testing should be offered with appropriate counselling regarding penetrance, especially for young children (rarely diagnosed <10 years). | *HNF1A* variants are also associated with hepatic adenomas and rarely with CHI. The penetrance of these conditions is low and there is no requirement to advise on risk to patient or family members. Their risk can be included in the report at the discretion of the testing laboratory. | 101, 72, 102, 103 |
| Pathogenic or likely pathogenic *HNF4A* variant | Consistent with/Confirms a genetic diagnosis of monogenic diabetes, subtype *HNF4A* | Patients with *HNF4A* diabetes respond well to sulphonylurea treatment. Risk of macrosomia and CHI for a newborn that inherits the variant from either parent; non-invasive prenatal testing may be available | 50% risk to offspring. Testing affected relatives is possible to confirm diagnosis. Predictive testing should be offered with appropriate counselling regarding penetrance, especially for young children (rarely diagnosed <10 years). | The *HNF4A* p.Arg114Trp variant is associated with a significantly reduced penetrance for diabetes and does not increase risk of macrosomia. Predictive testing for this variant should take this into consideration.<br>The p.Arg63Trp variant causes a Fanconi renal tubulopathy and renal assessment is recommended for these patients. 50% risk to offspring of inheriting variant and developing a tubulopathy. | <sup>3, 91, 104</sup> |
| Pathogenic or likely pathogenic *HNF1B* variant | Consistent with/Confirms a genetic diagnosis of monogenic diabetes, subtype *HNF1B* or RCAD<br>Consistent with/Confirms a genetic diagnosis of *HNF1B*-related renal disease (for a family member with renal disease but no diabetes). | If diabetes only, an assessment for *HNF1B* related disease (renal, urogenital, hepatic and pancreatic exocrine) should be recommended.<br>If a whole gene deletion is identified, implications for possible neuropsychiatric problems should be stated.<br>If the diagnosis is made in a relative with renal disease but no diabetes, recommendations for routine HbA1c screening should be given. | 50% risk to offspring of developing diabetes and *HNF1B* related conditions. Testing affected relatives is possible to confirm diagnosis. Predictive testing should be offered with appropriate counselling regarding penetrance, especially for young children (rarely diagnosed <10 years). Whole gene deletions frequently occur *de novo* and both parents should be tested to confirm this. | If a pathogenic *HNF1B* variant is identified unexpectedly in a patient with isolated diabetes, a simple request for additional clinical information may reveal undisclosed renal disease which will enable the reporting of the syndrome. | <sup>70, 105</sup> |
| Mitochondrial DNA variant m.3243A>G | Confirms a genetic diagnosis of mitochondrial diabetes or MIDD due to m.3243A>G variant.<br>Inconclusive result; further testing required | If diabetes only, recommend audiological assessment. Variant is associated with other clinical conditions but cannot predict disease progression due to heteroplasmy in different | All offspring of affected females are at risk of inheriting the variant. Males will not pass on the variant. Diagnostic testing offered to affected relatives. Predictive testing | Heteroplasmy level should be provided if measured. For low level heteroplasmy in peripheral blood (1-5%), confirmatory testing from a urine epithelial sample should be advised. | <sup>106, 107, 108</sup> |
|  | if heteroplasmy level is 1-5%. | tissues. Reproductive options may be mentioned with possible referral to a specialist mitochondrial disease clinic if available. | should be offered to at-risk offspring, siblings and maternal relatives through a clinical genetics service. |  |  |
| Pathogenic activating variant in *ABCC8* or *KCNJ11* in a patient with suspected MODY | Consistent with/Confirms a genetic diagnosis of monogenic diabetes, subtype *ABCC8/KCNJ11* | Patients with activating variants in *ABCC8* and *KCNJ11* respond well to sulphonylurea treatment. | 50% risk to offspring of developing neonatal diabetes and/or diabetes in early adulthood. Testing affected relatives is possible to confirm diagnosis. Predictive testing for TNDM causing variants outside of infancy should be offered with appropriate counselling, especially for young children since the penetrance is variable.. | Patient has a variant that causes remitting TNDM with relapsing diabetes in adulthood. In some patients, the TNDM is non-penetrant and the diabetes presents clinically as MODY and is referred for MODY testing. | <sup>109</sup>, <sup>110</sup>, <sup>111</sup> |
| Pathogenic activating variant in *ABCC8* or *KCNJ11* in a patient with NDM | Consistent with/Confirms a genetic diagnosis of permanent or transient neonatal diabetes, subtype *ABCC8/KCNJ11*. | Patients with activating variants in *ABCC8* and *KCNJ11* respond well to sulphonylurea treatment. Specific variants in *KCNJ11* cause DEND syndrome requiring higher doses of sulphonylurea. For patients with TNDM, the remitting and relapsing disease course should be described. Non-invasive prenatal testing may be available since early treatment with sulphonylurea (including trans-placental if mother has NDM) improves outcomes. | 50% risk to offspring of developing neonatal diabetes, or neonatal diabetes and/or diabetes in early adulthood if TNDM is diagnosed. Testing affected relatives is possible to confirm diagnosis. Predictive testing for a TNDM variant in older children and adults is possible with appropriate counselling regarding penetrance. | Sulphonylurea transfer protocols are available at https://www.diabetesgenes.org/about-neonatal-diabetes/.<br>Rarely, variants may be recessively acting and should be reported as for a recessively inherited condition. A recessive variant may cause neonatal diabetes if occurring *in trans* with a pathogenic inactivating (loss of function) variant. | <sup>110</sup>, <sup>109</sup>, <sup>6</sup>, <sup>112</sup>, <sup>113</sup>, <sup>114</sup> |
| Pathogenic dominant | Genetic diagnosis of monogenic diabetes | Dominantly acting inactivating variants cause | 50% risk to offspring of developing CHI and/or | The report must clearly distinguish these inactivating variants from the more | <sup>115</sup> |
| inactivating variant in *ABCC8* or *KCNJ11* in a patient with suspected MODY | due to a heterozygous inactivating *ABCC8* or *KCNJ11* variant | CHI and/or adult-onset monogenic diabetes.<br>In contrast in activating variants, these inactivating variants are not associated with increased sensitivity to sulphonylureas. | adult -onset diabetes. Testing relatives with CHI or diabetes is possible. Predictive testing is not recommended given the significantly reduced penetrance for diabetes due to these inactivating variants. | common activating variants causing diabetes, since the treatment protocols are different. Patients with dominant inactivating variants may respond to other OHAs similar to slim patients with type 2 DM. |  |
| Pathogenic variant in a syndromic monogenic diabetes gene | Consistent with/Confirms a genetic diagnosis of a monogenic diabetes syndrome.<br>If the patient does not have characteristic features of the syndrome, report as gene X-related monogenic diabetes. | Assessment for clinical features of the syndrome is recommended through referral to a disease specialist if available or through a clinical genetics service. | Testing affected relatives is possible to confirm diagnosis. Carrier testing offered to parents of patients with recessively inherited disorders. | If the diagnosis is unexpected, the laboratory should contact the clinician before reporting to establish if the patient has an undisclosed clinical diagnosis of the syndrome.<br>Patients heterozygous for a pathogenic variant in a recessively inherited disorder will be (with very rare exceptions) a carrier and do not have monogenic diabetes. This is an incidental finding and should not be reported but consider reporting if there is consanguinity in the family. | 4, 116, 34 |
| Variant of uncertain significance (general) | Inconclusive result; further investigations required. | Report should clearly state that the result does not confirm a genetic diagnosis of monogenic diabetes. Further clinical investigations should be requested as appropriate.<br>The result cannot be used in isolation for clinical decision making, although a trial change in therapy may be possible. | Testing of other affected relatives to determine whether variant segregates with diabetes. Knowledge of clinical phenotype of the relatives tested is essential. The result should not be used for predictive testing. | Class 3 (VUS or VOUS) variants should be reported based on professional judgement, the level of supporting evidence, possible impact on clinical management and on whether additional investigations can be undertaken to change the classification. Consider not reporting the variant if there is very weak supporting evidence (a 'cold' VUS) or variant is in a syndromic diabetes gene with no clinical suspicion of a syndrome. | 65 |
| Variant of uncertain significance in *GCK* | Inconclusive result; further investigations required. | The result should not be used in isolation for clinical decision making. Further biochemical testing (FBG, OGTT and HbA1c) to confirm *GCK* *MODY* phenotype.<br>A trial off treatment could be performed if safe to do so, with no change in HbA1c consistent with *GCK* *MODY*. | Segregation of the variant with fasting hyperglycaemia in the family should be investigated. Testing can be offered to clinically affected and unaffected relatives. All genetically tested relatives must undergo FBG and HbA1c testing. | Functional studies are likely to be beyond the scope of work offered by a diagnostic laboratory and should only be suggested if the laboratory can provide this. RNA studies on variants predicted to affect splicing are possible using mini-gene assays and these may be offered by the laboratory. | <sup>117, 118</sup> |
| Variant of uncertain significance in *HNF1A* or *HNF4A* | Inconclusive result; further investigations required. | The result should not be used in isolation for clinical decision making.<br>A trial on sulphonylurea could be performed if appropriate with a good response or hypoglycaemia on low doses consistent with *HNF1A*/*HNF4A* diabetes. Details of birth weights and history of CHI in proband and relatives should be requested. | Segregation of the variant with diabetes in the family should be investigated. Clinical details of tested relatives should be collected to assess whether they have a *MODY* phenotype since phenocopies with type 1 and 2 DM can be present in families. Predictive testing should not be offered. | Functional studies are likely to be beyond the scope of work offered by a diagnostic laboratory and should only be suggested if the laboratory can provide this. RNA studies on variants predicted to affect splicing are possible using mini-gene assays and these may be offered by the laboratory. | <sup>119</sup> |
| Variant of uncertain significance in *ABCC8* or *KCNJ11* | Inconclusive result; further investigations required. | Ask if there is a history of NDM or CHI in the proband and their family.<br>If strong suspicion of a *PNDM* or *TNDM* variant, consider a trial on sulphonylurea with a good response or hypoglycaemia | Parental testing for a heterozygous variant in an NDM proband is recommended to check for *de novo* occurrence.<br>Segregation of the variant with diabetes in the family should be investigated. | Careful consideration needed as to whether the variant should be reported given complexity of interpretation. The variant will typically fall into one of four categories:<br>1. Pathogenic *TNDM* variant with non-penetrance of NDM. | <sup>120</sup> |
|  |  | on low doses consistent with a genetic diagnosis. | Clinical details of tested relatives should be collected to assess whether they have a MODY phenotype since phenocopies with type 1 and 2 DM can be present in families. Predictive testing should not be offered. | 2. Pathogenic variant causing dominant CHI with non-penetrance of CHI.<br>3. Pathogenic recessively acting CHI variant and patient is a carrier of CHI.<br>4. Benign variant of no clinical significance.<br>Heterozygous null variants in these genes (nonsense, frameshift, splicing) should not be reported (unless there is consanguinity in the family) since they do not cause diabetes and patients are carriers of recessive CHI. |  |
| No variant detected (NGS) | A genetic diagnosis of monogenic diabetes has not been confirmed | Additional specific testing may be offered/advised e.g. CNV testing, MS-MLPA for 6q24 related TNDM or genotyping for m.3243A>G | N/A. | Gene panels should analyse the common causes of monogenic diabetes and perform CNV analysis. The report should list all genes analysed, horizontal coverage and the types of variants that can be identified. Limitations should be stated and will include genes not sequenced by the NGS assay, not detecting CNVs, imprinting abnormalities, non-coding variants and mtDNA variants, and regions of low sequencing coverage. Sanger in-fill may be used for regions with low coverage.<br>Detailed technical and bioinformatics information may be added as an appendix but not in the main body of the report. | <sup>44</sup>, <sup>65</sup> |
| No variant detected in gene X (Sanger) | Does not confirm a genetic diagnosis of monogenic diabetes, subtype gene X | Result does not exclude a diagnosis since pathogenic variants in other genes can cause monogenic diabetes. | N/A. | Report should state the limitations of the method used, i.e. that other monogenic diabetes genes have not been sequenced, but may also state that CNVs or variants in non-coding regions cannot be detected. | <sup>121</sup>, <sup>122</sup> |
|  |  | Either reflex to additional testing (e.g. NGS), offer/advise additional genetic testing as appropriate or advise that no further genetic testing is indicated. |  |  |  |
| m.3243A>G variant not detected (genotyping on blood DNA) | Does not confirm a genetic diagnosis of mitochondrial diabetes or MIDD due to m.3243A>G variant. | Does not exclude a diagnosis since the variant may be present at very low heteroplasmy levels or present in other tissues such as muscle or urine epithelia.<br>Urine testing should be offered for not detected tests on peripheral blood DNA in patients with clinical suspicion of MIDD. | N/A. | Report should state the method used and the heteroplasmy limit of detection. | <sup>106, 108</sup> |
| Predictive testing | Genetically predisposed to monogenic diabetes, subtype X<br>Not genetically predisposed to monogenic diabetes, subtype X | If variant is not detected, state that the family member's risk of developing diabetes is reduced to that of the population.<br>If the variant is detected, then state that periodic monitoring for diabetes by HbA1c is recommended. If diabetes penetrance is age-related (e.g. HNF1A) and family member is a young child, then consider delaying monitoring until >10 years. | If variant is detected, state risk to offspring of inheriting the variant. Offer testing to affected relatives to confirm diagnosis. Predictive testing should be offered with appropriate counselling regarding penetrance, especially for young children (rarely diagnosed <10 years). | If predictive testing is undertaken for both parents of a proband, and the variant is not detected in either parent, the variant is likely to have arisen de novo in the proband. Germline mosaicism is a possibility; The parents' reports should state the lowest level of mosaicism that the assay could theoretically detect (e.g 10% for Sanger), and state that the risk of having another affected child is low but risk due to germline mosaicism cannot be excluded. | <sup>72, 123</sup> |
| Relative with diabetes and | Diagnosis of monogenic diabetes, | Consider the clinical features of the relative. If not suggestive of monogenic diabetes, then state that a | N/A | Sanger sequencing is the most likely assay used for familial variant testing but is prone to false negative results due to sequence variation under primer binding sites that |  |
| familial variant not detected | subtype gene X has not been confirmed | diagnosis has not been confirmed and that the diabetes is likely to have a different aetiology. If features are consistent with monogenic diabetes and result is unexpected, request repeat samples for genetic testing using a different method (e.g. NGS or Sanger with alternative primers). |  | may prevent the variant allele being sequenced. If the not detected result is unexpected, the lab should re-test using new primer sequences that have been checked for common variants in binding sites. NGS testing is another option since it is not prone to this issue and also tests for all other causes of monogenic diabetes, but is more expensive. | <sup>44</sup>, <sup>64</sup> |

NDM - Neonatal Diabetes Mellitus; CHI - Congenital Hyperinsulinism; RCAD - Renal Cysts And Diabetes syndrome; MIDD - Maternally Inherited Diabetes and Deafness syndrome; TNDM- Transient Neonatal Diabetes Mellitus; VUS or VOUS - Variant Of Uncertain Significance; PNDM - Permanent Neonatal Diabetes Mellitus; CNV - Copy Number Variant; MS-MLPA - Methylation-Specific Multiplex Ligation-Dependent Probe Amplification; NGS - Next Generation Sequencing.

Based on our systematic review of the literature we can make several recommendations **(Display Box 2)**. A targeted NGS approach is the preferred option for testing MODY and NDM to maximize diagnostic yield without significant cost and variant interpretation burden compared to gene agnostic genome sequencing. Genome sequencing can provide data for novel gene and non-coding variant discovery and allows re-analysis for newly associated genes and variants but is prohibitively expensive for many laboratories and requires significant bioinformatics expertise to manage the huge numbers of variants and give correct classifications. Targeted panels should be designed to include all known causes of monogenic diabetes including mitochondrial diabetes, detect known non-coding mutations (located in promoters, deep introns and distal enhancers) and detect CNVs. A comprehensive gene panel that includes all recessively inherited genes is essential in countries and populations with high rates of consanguinity. A separate MLPA assay for CNV detection or genotyping assay such as pyrosequencing for m.3243A>G detection is acceptable but comes at increased cost. NDM testing services should offer a methylation-based assay such as MS-MLPA since 6q24 imprinting defects are a common cause of TNDM. The high diagnostic yield for *GCK* in suspected GCK-MODY and *KCNJ11*, *ABCC8* and *INS* in NDM, and the clinical utility of these diagnoses, justifies rapid Sanger sequencing of these genes initially in these scenarios.

**Box 2:**
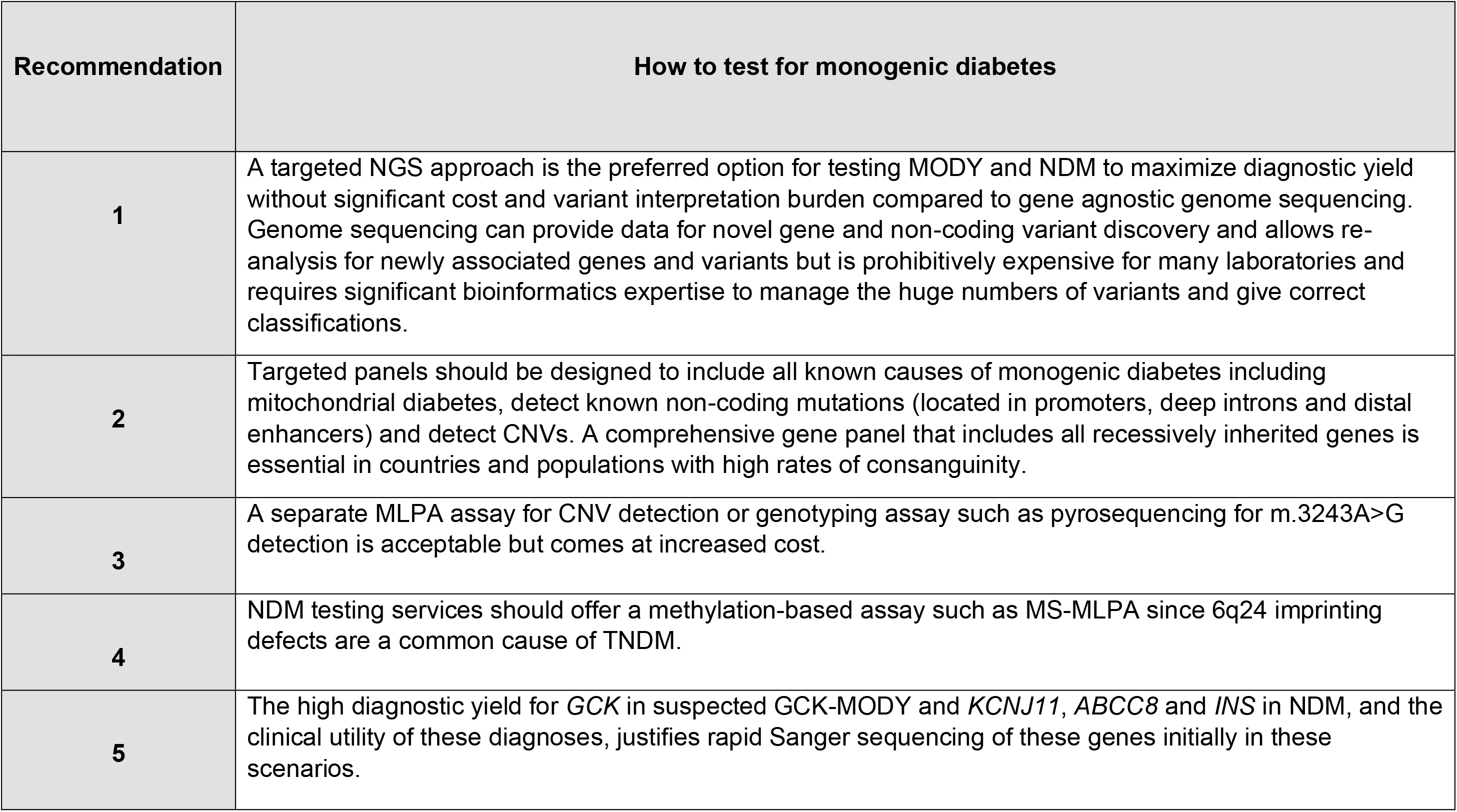
Recommendations based on synthesis of evidence for how to test for monogenic diabetes.

### Question 3: - What is the basis for considering a gene as a cause of monogenic diabetes

A general evidence-based framework for evaluating gene disease validity has been developed by the ClinGen and published by an inter-institutional group of clinical and molecular genetics and genomics experts^38^. This framework involves evaluating case level, segregation, and functional data for previously reported variants and functional data for the gene itself to classify gene-disease validity relationships into Definitive, Strong, Moderate, Limited, Disputed, or Refuted categories based on a point system combined with expert consensus for the final assignment. Tools for implementing this are available at the ClinGen website. The international MDEP GCEP has convened with the goal of curating gene-disease validity for MODY and other monogenic diabetes genes and has completed the MODY genes (https://www.clinicalgenome.org/affiliation/40016/) and is working on expanding beyond these genes. Other general repositories for gene-disease validity curation include the crowd-sourced Genomics England PanelApp^37^. For monogenic diabetes, a curated list of monogenic diabetes genes is available at the website for the University of Exeter, where most of the research and clinical monogenic diabetes testing for the entire UK is conducted (https://www.diabetesgenes.org/).

Over recent years the increased availability of high throughput sequencing has led to a substantial increase in the number of genes reported to cause monogenic diabetes. The evidence that supports these gene-disease relationships does, however, vary widely. Whilst there is overwhelming genetic evidence that established the etiological role of genes such as *HNF1A*, *HNF4A* and *GCK*, recent studies that have investigated variation in genes such as *BLK*, *KLF11* and *PAX4* in large population datasets have not supported their role in causing monogenic diabetes^62^, and these genes were recently refuted as monogenic diabetes genes by the MDEP GCEP.

The consensus opinion of the writing group was that a gene should only be considered causative of monogenic diabetes if it meets the criteria set out in expertly curated guidelines that have been developed to validate gene-disease relationships. These guidelines have already been applied to many of the monogenic diabetes genes by the ClinGen MDEP GCEP. We recommend continued efforts to curate new and updated existing monogenic diabetes genes for gene-disease validity be centralized with the MDEP GCEP. Those interested in contributing to this effort should engage with the MDEP GCEP to ensure that genes used in monogenic diabetes have been curated for gene disease validity in a process that is evidence based and updated on a standard schedule as directed by ClinGen.

### Question 4: - On what basis should a variant be considered a cause of monogenic diabetes?

**I**n 2015, the ACMG and AMP developed general guidelines for the interpretation of sequence variants^39^. The ClinGen Sequence Variant Interpretation (SVI) Working Group has published multiple updates to these original guidelines^40–43^. The Association for Clinical Genomic Science (ACGS) voted to adopt these guidelines^63^. These guidelines have undergone several updates. ClinGen’s MDEP VCEP has modified these general guidelines for three MODY-causing genes (*HNF1A*, *HNF4A* and *GCK*); these guidelines account for many issues inherent in the difficulty in interpreting monogenic diabetes variants and can be used as a framework for interpreting variants in genes for which rules have not yet been established.

The ACMG/AMP guidelines were developed through an evidence-based process involving the sharing, developing, and validating of variant classification protocols among over 45 laboratories in North America. They incorporate various types of evidence to determine if a variant is pathogenic, likely pathogenic, of uncertain significance (VUS), likely benign, or benign. Examples of the types of evidence include: frequency in public databases such as gnomAD; the segregation of a variant with a disease phenotype; results of computational (*in silico*) prediction programs; *de novo* status; functional studies; frequency of variant in cases vs. controls; the presence of other pathogenic variants at the same nucleotide or within the same codon; the location of a variant (i.e., if it is within a well-established functional domain or mutational hotspot); and whether a variant has been found in a patient with a phenotype consistent with the disease. MDEP gene-specific rules incorporate experts’ unpublished case data and knowledge of monogenic diabetes phenotype and prevalence in recommending the evidence and thresholds to apply.

Continued work by MDEP VCEP is needed to develop applications of the guidelines tailored to additional monogenic diabetes types and genes. Improvement in de-identified case-sharing platforms is needed to promote maximizing the ability to gather the evidence needed to evaluate pathogenicity.

### Question 5: - How should a variant in a monogenic diabetes gene be reported?

Well written general guidelines for the reporting of genetic test results are available ^44,45, 64–68^ and this review will therefore summarize the basic requirements and focus on reporting monogenic diabetes tests.

We summarize the recommendations for reporting results for a range of different testing scenarios and methodologies (**Table 3**). A single page report with appendices is preferred. The report should restate the reason for testing, including the clinical characteristics/phenotype of the patient. The report must include a headline result or summary that clearly states the outcome of the test for the patient – this may be stating whether a diagnosis of monogenic diabetes has or has not been made, or whether a patient is or is not genetically predisposed to monogenic diabetes. Patients with specific subtypes may respond well to certain therapies and this should be noted in the report. Testing should be offered to at-risk family members, which may be diagnostic, predictive or carrier testing. Special care should be taken when reporting variants in syndromic diabetes genes in patients with isolated diabetes. The risk to future offspring should be stated according to mode of inheritance. The report should not use terms positive or negative for describing test results. Variants should be reported in a table that includes the HUGO gene name, zygosity of the variant, both nucleotide and protein level descriptions using HGVS nomenclature, genomic coordinates and the classification of the variant based on the ACMG/AMP 5 level classification system^39^. Benign and likely benign variants should not be reported. Class 3 (VUS or VOUS) variants should be reported based on professional judgment, the level of supporting evidence and on whether additional investigations can be undertaken to change the classification such as testing of other affected relatives, further biochemical testing, or additional functional laboratory investigations. Evidence used to classify the variant should be clearly outlined. Technical information should be provided in a section separate from results and interpretation and will include details of the methodology and gene or genes tested. If the testing performed does not cover all known genes and possible mutations, then this should be stated as a limitation with recommendations for further genetic testing (e.g., NGS or MLPA analysis).

The structure, format, and content of MODY reports will vary widely between laboratories across the world. Standardization is difficult due to variability in mandatory report content, such as legal disclaimers, and the ability to include clinical recommendation. But there are essential reporting best practices that should be adopted by all laboratories irrespective of local reporting policies. We recommend that laboratories performing monogenic diabetes testing participate in the EMQN’s monogenic diabetes EQA scheme (www.emqn.org) which aims to educate and improve quality of diagnostic testing and reporting for this condition. Future research is advised to engage patients, providers, and other stakeholders in the design and evaluation of readability, comprehension, and application of information contained in genetic testing reports for monogenic diabetes.

### Question 6: - Research Question: What are the next steps after a diagnosis of monogenic diabetes?

A systematic, comprehensive, and collaborative approach is required after making a monogenic diabetes diagnosis after conducting genetic testing. Our guidance for the next steps after diagnosis of monogenic diabetes focuses on the following: 1) practical recommendations for providing the diagnosis results and clinical follow-up, 2) reviewing genetic testing reports, 3) family testing for adults and children, 4) legal considerations for this diagnosis, 5) considering psychological impact of diagnosis, and 6) recommendations for addressing VUS results and negative monogenic diabetes testing despite atypical features to a patient’s diabetes presentation. In the following paragraphs, the term “clinician” can refer to a physician or genetic counselor. Genetic counselors are specially trained to communicate complex genetic information, facilitate family testing, and address psychosocial issues that may arise with a new diagnosis; thus, we recommend having a genetic counselor as part of the care team if possible. Upon receipt of a genetic test result diagnosing monogenic diabetes (i.e., pathogenic, or likely pathogenic variant identified), the clinician should schedule a 30-60 minute in-person or telehealth appointment with the patient/family^69^. We do not recommend that results be disclosed via an electronic health record (EHR) portal or by clinic staff. Under the 21^st^Century Cures Act in the US, genetic test results may be released to the patient via the EHR before the provider can see them. In this case, we recommend that clinicians discuss whether to consider these results as sufficiently sensitive to delay the immediate release until a provider has had a chance to review with the patient. If delaying the release of genetic test results is not an option, the provider should discuss the possible timing of results in the EHR when planning for results disclosure during pre-test counseling.

After a very brief reminder of what the genetic test analyzed, we recommend the clinician describe the identified variant in patient-friendly language (e.g., a single spelling error in the genetic code) and review how disease-causing variants in the gene impair glucose metabolism. The clinician can explain the evidence used to classify the variant as disease-causing which is often included in the genetic testing report, e.g., if the variant was previously identified in patients with monogenic diabetes or experimental evidence demonstrated loss of function. The clinician should describe the general features of the type of monogenic diabetes indicated by the genetic change, including the inheritance pattern of the disorder, specifying those features that are consistent with the patient’s clinical picture. If the type of monogenic diabetes is characterized by variable expressivity and/or reduced penetrance, these concepts should be introduced to the patient/family, providing specific examples from the disorder at hand. HNF1B syndrome is a prime example of variable expressivity, as the renal and extra-renal phenotypes (diabetes, genital malformations, pancreatic hypoplasia, abnormal liver function) vary among affected individuals, even within the same family^70,71^. The patient/family should be provided a copy of the report for their records. Additionally, a document describing the variant identified and avenues for variant-specific testing can be provided to the patient to distribute to family members if family testing is being pursued. Upon reflecting on the diagnosis, patients may feel relief at a genetic etiology for their symptoms, while others may feel angry or annoyed if they were initially misdiagnosed and prescribed suboptimal treatment^72–77^. Feelings of frustration should be validated. Some patients may find solace in hearing that knowledge and testing of monogenic diabetes have both evolved greatly over time and we hope more diagnoses will be made moving forward. Patients may also be helped by speaking to other patients with monogenic diabetes. At this time, formal support groups are limited for monogenic diabetes, but the provider can consider connecting patients with monogenic diabetes given mutual consent. Patients, providers, and researchers are in the process of creating a consortium for communication and support regarding monogenic diabetes called the Monogenic Diabetes Research and Advocacy Consortium (MDRAC, mdrac.org). Yearly follow-up can be suggested to continue to provide updates on the monogenic diabetes diagnosis, prognosis, and treatment in addition to any new information on the gene and genetic variant identified.

Results of genetic testing should be discussed in context of the family history. The most common forms of monogenic diabetes, HNF1A, HNF4A, and GCK etiologies, are dominantly inherited, and the vertical transmission of diabetes or hyperglycemia is often evident in the pedigree^4,10,78,79^. If a disease-causing variant in one of these conditions is identified in a parent of an affected individual, there is a 50% chance that siblings and children of the proband will inherit the variant. The absence of a family history of diabetes may suggest that a variant associated with a dominant condition is *de novo* in the proband. If parents test negative and maternity and paternity are confirmed, the recurrence risk in siblings is approximately 1%, which accounts for the possibility of gonadal mosaicism^80^. *De novo* disease-causing variants have been reported and are especially common in *HNF1B* ^70,81,82^. With HNF1B ethology, the family history may also include genital tract malformations, renal cysts, or pancreatic hypoplasia^70^. Recurrence risk of recessive forms of monogenic diabetes, such as Wolcott-Rallison syndrome or Rogers syndrome, is 25% in offspring when both the proband and their partner are carriers^4^. Recurrence risk of monogenic diabetes caused by the mitochondrial DNA MIDD (maternally inherited diabetes and deafness) variant (m.3243A>G) is essentially zero when the sperm-producing parent has the variant, as mitochondria are passed down through the oocyte. All offspring and maternal relatives of the egg-producing parent will inherit the variant, albeit at varying heteroplasmy^83^.

Affected family members of individuals with molecular confirmation of monogenic diabetes should be offered variant-specific testing of the familial variant, a process known as cascade testing^10,69^. For probands with GCK-ethology for mild, persistent fasting hyperglycemia, it is important to also discuss cascade testing of family members with gestational diabetes and pre-diabetes, since this is characterized by stable, mild fasting hyperglycemia that is clinically asymptomatic and can also impact pregnancy management in a gestational parent with apparent GDM. Unaffected or undiagnosed first-degree relatives of probands with GCK ethology should undergo a fasting glucose test; if normal, a diagnosis of GCK related mild, persistent fasting hyperglycemia is highly unlikely and genetic testing is unnecessary^10,84,85^.

The risks and benefits of testing, and possible results of testing, should be reviewed in all cases to allow the family to make autonomous testing decisions consistent with their goals and values. Possible benefits of genetic testing include the ability to obtain or advocate for more appropriate treatment, reduced anxiety, and uncertainty, decreased stigma, knowledge of recurrence risk, and the ability to plan for the future^69,72,76,86^. Risks may include increased anxiety, trouble adjusting to a new diagnosis, or learning unexpected information^74^. The risk of insurance discrimination may also need to be reviewed, as different countries have instituted varying rules regulating the use of genetic information in insurance underwriting. In the U.S., the 2008 Genetic Information Nondiscrimination Act (GINA) is a federal law that prevents against discrimination from health insurers and employers based on genetic information. In other words, a health insurer cannot make coverage decisions based on a genetic risk for a medical condition, and an employer cannot consider risk for a genetic disease when making hiring, etc. decisions. GINA’s protections do not extend to employers with less than 15 employees and companies that sell life, long term care and disability insurances. GINA also has limitations with employment and insurance within the U.S. Military. Importantly, GINA does not provide protection from discrimination based on already-diagnosed conditions. As most patients who undergo genetic testing for monogenic diabetes do so because they are symptomatic, it is unclear the extent to which a molecular diagnosis of monogenic diabetes would affect the ability of such a patient to obtain a new life, long term care, or disability insurance policy. The exception to this may come if a diagnosis of GCK related mild, persistent fasting hyperglycemia is confirmed, and a company decides to approve/improve coverage based on the good prognosis of this condition. GINA should be discussed in the context of testing asymptomatic family members, especially children, for an identified variant, given the uncertain impact on insurance of having a positive genetic test result in the medical record of someone without diabetes (https://www.genome.gov/about-genomics/policy-issues/Genetic-Discrimination).

Additional ethical and psychosocial issues surrounding a genetic diagnosis should also be discussed when considering predictive testing in a minor. The clinical relevance of an *HNF1A* or *HNF4A* positive genetic test would likely have minimal clinical relevance prior to adolescence, and thus we generally discourage genetic testing in young children. Indeed, adolescents in families with HNF1A monogenic diabetes preferred testing in adolescence when parents and their children can engage in joint decision-making regarding genetic testing^73^. We do not recommend testing asymptomatic children for GCK persistent elevated fasting hyperglycemia, given this is a benign condition and there are potential adverse psychosocial effects of being labeled as “sick” ^86^. Also, the “GCK-MODY” diagnosis can lead to problems achieving life, long term care, or disability insurances since “GCK-MODY” is classified as a monogenic form of diabetes but in practice is a benign condition. If a child in a family with a GCK ethology is incidentally found to have hyperglycemia, their pediatrician should be informed of the familial variant and familial variant testing can proceed to avoid unnecessary treatment. This strategy also respects the autonomy of the child to make an informed decision about testing when they are able. However, the significant fear of uncertainty that some parents of at-risk children feel should not be dismissed. Genetic testing may decrease anxiety in parents, allow them to gradually introduce the disorder to their child in a developmentally appropriate manner, and empower them to prepare for the future^72,86^.

A positive result of genetic testing would replace the prior diagnosis (of type 1 or type 2 diabetes) with a diagnosis of monogenic diabetes. The clinician should review the prognosis of the condition and potential changes in medical management (e.g., no treatment in GCK persistent elevated fasting hyperglycemia and sulfonylurea treatment with HNF1A and HNF4A monogenic diabetes^10,85^. We refer the reader to the recommendations generated by the Monogenic Diabetes Precision Prognostics and Therapeutics groups for additional information which we also provide a high-level summary of in **Figure 3**.

There may be instances when an asymptomatic family member has a positive result on genetic testing. In this case, the high risk of developing diabetes or hyperglycemia should be emphasized and a plan for monitoring blood sugars should be developed if appropriate. Of course, an asymptomatic family member with negative variant-specific testing may still be at risk of developing more common forms of diabetes^69^. A notable exception to this is that asymptomatic family members, with negative genetic testing of probands with MIDD, are still at risk for diabetes, hearing loss, and potentially other symptoms of mitochondrial disease given the variability in heteroplasmy of the m.3243A>G variant among different body tissues^83^. A negative result of variant-specific testing in a family member with diabetes would indicate another etiology for the diabetes diagnosis, such as type 1 or type 2 diabetes.

Opportunities exist for individuals with a genetic diagnosis of MODY and other forms of monogenic diabetes to participate in research. The Monogenic Diabetes Registry at the University of Chicago in the US is open to individuals with various forms of monogenic diabetes (https://monogenicdiabetes.uchicago.edu/). This study is also open to individuals with a clinical picture consistent with monogenic diabetes, but whose genetic testing was negative or returned a variant of uncertain significance. Such individuals can also consider the Rare and Atypical Diabetes Network (RADIANT) study in the US, which aims to identify new forms of atypical diabetes (https://www.atypicaldiabetesnetwork.org/). Variants of uncertain significance can also be directed to the ClinGen Monogenic Diabetes Expert Panel (MDEP, described above) for further review (https://clinicalgenome.org/affiliation/50016/). In some cases, functional studies of unresolved gene variants can be helpful to decide pathogenicity^87,88^. Several groups in Europe (www.uib.no/en/diabetes) and North America https://med.stanford.edu/genomics-of-diabetes.html) are able to study VUS in monogenic genes that they have domain expertise in and work in close collaboration with the ClinGen Monogenic Diabetes Expert panel to interpret results from functional assays. In the UK, the Genetic Beta Cell Research Bank (GBCRB) is a tissue bank at the Exeter Laboratory that uses residual clinical samples for research on the genetics of diabetes (https://www.diabetesgenes.org/current-research/genetic-beta-cell-research-bank/).

### Question 7: - What are the current challenges for the field in precision diagnostics for monogenic diabetes?

We reviewed the abstracts of 455 abstracts and the full text of 42 papers before extracting data from 14 articles meeting our criteria. Several key themes emerged which present on-going challenges for the field of precision diagnostics for monogenic diabetes. With the generation of exome and now genome sequencing data from both larger clinical cohorts and biobanks we have new insights into variant frequency and penetrance. For some previously reported pathogenic variants in monogenic diabetes genes there is now evidence for reduced penetrance in unselected populations and that individuals carrying the variants do not necessarily display the hallmark characteristics (e.g BMI <30kg/m^2^, age of diagnosis <35 years) of monogenic diabetes ^89–92^. The interpretation of novel rare variants in monogenic diabetes genes is challenging, functional studies can assist but multiple assays are required in concert with frequency and clinical data^87,93^. Functional studies are slow, lack standardization and are usually retrospectively performed after variant discovery. Efforts to generate variant maps in genes of interest are a potential route forward but will require coordination and implementation of standards^94^. The perpetuation of errors in the literature remains a concern with ongoing reporting of novel variants in genes which are not considered by experts in the field to be causal for monogenic diabetes^95^. Whilst the reporting of potential novel genes can be misleading as they do not necessarily meet the criteria for classification as a novel genetic cause of diabetes^96^. There remain inequalities in sequencing data across diverse ancestries and populations even when there are examples of the importance of rare variation in monogenic diabetes genes ^97–99^.

Finally, barriers to genetic testing remain including limited provider awareness of monogenic diabetes. It is important that all clinicians treating diabetes patients are considering monogenic etiologies as a potential diagnosis, especially when diagnosis can occur in adults that have had diabetes since youth^100^. Future research should focus on increasing representation of sequence data in monogenic diabetes genes in diverse populations, generating variant maps of clinically actionable diabetes genes and continued efforts to share knowledge and expertise of monogenic diabetes in underserved communities and populations.

## Discussion

To diagnose monogenic diabetes offers an opportunity to find those who can benefit from precision medicine^3,4^. Phenotypic overlap with other diabetes subtypes^14^ and lack of awareness^28^ often results in misclassification of monogenic diabetes and hence missed prospects for optimal management. Although several diagnostic technologies for genetic testing are available, which technology to choose is a balance between cost, time, scientific and bioinformatic expertise required and the diagnostic yield. To fill these knowledge gaps, we aimed to systematically review the yield of monogenic diabetes using different criteria for how to select people with diabetes for genetic testing and which technologies to use. Moreover, we sought to evaluate current guidelines for genetic testing.

Why a systematic review? Most of the time a single study does not tell us enough. The best answers are found by combining the results of many studies. We therefore applied a systematic literature search which is widely recognized as a critical component of the systematic review process. It involved a systematic search for studies aiming for a transparent report of study identification, leaving clear what was done to identify studies, and how the findings of the review are relative to the evidence base. The relevant parts within each article were then evaluated and re-evaluated, with the aim of determining key methodological stages. These were identified and defined. This data was reviewed to identify agreements and areas of unique recommendations between the articles.

In this article, we provide recommendations on practical steps for communicating a diagnosis of monogenic diabetes to patients, methods for family testing, and considering the psychological impact of diagnosis **(Display Boxes 1-2; Figure 3)**. The practice of communicating genetic testing results for monogenic diabetes to patients with a genetic diagnosis is evolving as monogenic diabetes testing becomes more prevalent. Although communicating genetic testing results for disease-causing variants is more straightforward, it remains challenging in communicating results of a VUS or a no genetic diagnosis resulting in a patient with a clearly atypical presentation of diabetes. Fortunately, collaborative efforts in variant curation and precision medicine research will continue to reduce the ambiguity in VUS or no diagnosis results and improve our ability to effectively provide recommendations for diagnosis, treatment and family testing for people who undergo monogenic diabetes testing.

The major strength with our study is being first of its kind and a comprehensive overview of all available evidence on diagnostics of monogenic diabetes based on screening more than 12,500 peer reviewed articles published during the last 32 years extracting data from >100 studies including 42,775 participants that met the predefined criteria. This makes it easier for healthcare professionals to make evidence-based decisions. We used rigorous and transparent methods, leading to a higher quality of the evidence for how to use precision diagnostics in monogenic diabetes than other types of studies. Moreover, we aimed to reduce bias in the selection of studies, data extraction, and analysis making our findings more reliable and credible. Finally, our systematic review is an efficient way to identify knowledge gaps and prioritize future research, as it avoids duplication of efforts and resources.

Our study also has several limitations. For the question of who to test, the index/triage test of clinical or laboratory biomarkers used to select people for monogenic diabetes testing would ideally be compared with the reference standard of genetically testing all individuals with diabetes (without any such selection) however, such studies were rare. Most were cohort or cross-sectional studies in patients with diabetes diagnostic uncertainty, that only genetically tested a smaller sample by certain criteria, so it was not possible to discern the number of cases missed (false negatives) with this approach. Only a few studies directly compared two or more approaches in the same study population, so most recommendations were based on comparative yields in different populations. We were not able to provide recommendations on selecting whom to provide genetic testing for syndromic forms including mitochondrial diabetes and deafness, severe insulin resistance, lipodystrophy, or obesity related diabetes phenotypes with monogenic etiologies.

It was limited by the availability of relevant studies, and sometimes there were not enough high-quality studies to draw meaningful conclusions. Hence, we were not able to address Questions 3-7 initially or ultimately (for Questions 6-7) by a systematic review using the method offered by Covidence. However, the co-authors have been working on diagnostics of monogenic diabetes for 10-30 years and are experts in the field. We therefore used expert opinion for the Questions 3-7. Another weakness is that only papers in the English language were included in the analysis. Thus, non-English papers potentially offering useful information on Questions 1-2 were missed. It is, however, not likely since we defined a cut-off of 100 study individuals genetically that were tested to ensure a high scientific quality. Conducting the systematic review was a time-consuming process searching for and evaluating many studies. It was also resource-extensive necessitating a team of trained researchers and specialized software. And we cannot completely exclude publication bias, where only studies with significant results are published, and non-significant results are not reported. Despite some limitations, we believe our systematic review will prove a valuable tool in precision diagnostics of monogenic diabetes providing high-quality evidence to inform clinical decision-making.

What is needed next? Our systematic review reveals that improved access to genetic testing for monogenic diabetes to prevent health disparities is important. There are issues regarding equity and utility in non-European countries where background prevalence of type 2 diabetes is higher. Moreover, type 1 diabetes genetic risk score (a tool for using common susceptibility variants for type 1 diabetes to pre-assess the likelihood of having type 1 vs. other types of diabetes) data has not been well characterized in these countries. Another step is generation of and access to systematic measurements of autoantibodies and C-peptide for people diagnosed with diabetes under the age of 45 years with the addition of validated ancestry-appropriate type 1 diabetes genetic risk score data. This information would be advantageous to better discriminate monogenic diabetes from type 1 diabetes. What is more, improvement in de-identified case-sharing platforms is needed to promote maximizing the ability to gather the evidence needed to evaluate pathogenicity. As such, continued work by expert panels such as the MDEP VCEP is warranted to develop guidelines for which gene variants should be considered causative of monogenic diabetes as well as applications of the guidelines tailored to additional monogenic diabetes types and genes. One relevant instrument is generation of deep mutational scanning maps of monogenic diabetes genes to aid variant classification. It is also important to remember that the genetic and genomic testing landscape is ever evolving, with a strong possibility of universal genome sequencing in the future, which would reduce concerns on whom to test but place an even higher burden on having adequate tools, expertise, and workforce for interpretation. Finally, further clinical guidance is needed for steps following monogenic diabetes testing which includes genetic counseling, subsequent referrals, and family testing in addition to research on the outcomes of implementation.

## Data Availability

No original data is present in this manuscript it is a systematic review.

## Acknowledgements

The ADA/EASD Precision Diabetes Medicine Initiative (PMDI), within which this work was conducted, has received the following support: The Covidance license was funded by Lund University (Sweden) for which technical support was provided by Maria Björklund and Krister Aronsson (Faculty of Medicine Library, Lund University, Sweden). Administrative support was provided by Lund University (Malmö, Sweden), University of Chicago (IL, USA), and the American Diabetes Association (Washington D.C., USA). The Novo Nordisk Foundation (Hellerup, Denmark) provided grant support for in-person writing group meetings (PI: L Phillipson, University of Chicago, IL).

SM has a personal award from Wellcome Trust Career Development scheme (223024/Z/21/Z) and holds Institutional funds from the NIHR Biomedical Research Centre Funding Scheme. P.R.N. was supported by grants from the European Research Council (AdG #293574), Stiftelsen Trond Mohn Foundation (Mohn Center of Diabetes Precision Medicine), the University of Bergen, Haukeland University Hospital, the Research Council of Norway (FRIPRO grant #240413), the Western Norway Regional Health Authority (Strategic Fund “Personalised Medicine for Children and Adults”), and the Novo Nordisk Foundation (grant #54741). J.M.I. was supported by a grant from the National Institute of Health (K08DK133676). S.E.F. is supported by a Wellcome Trust Senior Research Fellowship (Grant Number 223187/Z/21/Z). ALG is a Wellcome Trust Senior Fellow (200837/Z/16/Z) and is also supported by NIDDK (UM-1DK126185). EDF is a Diabetes UK RD Lawrence Fellow (19/005971). KRO is supported by the National Institute for Health Research (NIHR) Oxford Biomedical Research Centre (BRC). The views expressed are those of the author(s) and not necessarily those of the NHS, the NIHR or the Department of Health.

## Author Contributions

*Review Design*: RM, KC, TIP, PRN, LKB, KRO, ALG

*Systematic Review Implementation*: RM, KC, TIP, JIM, PS, KAM, CS, JM, SM, IA, EdF, SEF, PRN, LKB, KRO, ALG

*Data extraction manuscripts:* RM, KC, TIP, JMI, PS, LKB, KRO, ALG *Manuscript writing:* RM, KC, TIP, LKB, KAM, PRN, KRO, ALG *Project Management:* RM, LKB, KRO, ALG

