## Supplementary Tables and Figures for "A Systematic Review of the use of Precision Diagnostics in Monogenic Diabetes"

#### Supplementary Table 1: Search strategy using PubMed

| Question 1: Who should you test for monogenic diabetes | Number of References |
|---|---|
| **Query** | |
| **#1**<br>Diabetes Mellitus, Permanent Neonatal" [Supplementary Concept]) OR ( "Maturity-Onset Diabetes Of The Young, Type 9" [Supplementary Concept] OR "Maturity-Onset Diabetes of the Young, Type 7" [Supplementary Concept] OR "MODY, Type 6" [Supplementary Concept] OR "Maturity-Onset Diabetes of the Young, Type 8, with Exocrine Dysfunction" [Supplementary Concept] OR "Maturity-Onset Diabetes of the Young, Type 1" [Supplementary Concept] OR "Maturity-Onset Diabetes of the Young, Type 2" [Supplementary Concept] OR "Maturity-Onset Diabetes of the Young, Type 3" [Supplementary Concept] OR "Maturity-Onset Diabetes of the Young, Type 4" [Supplementary Concept] )) OR ( "Lipodystrophy, Congenital Generalized"[Mesh] OR "Diabetes Mellitus, Congenital Autoimmune" [Supplementary Concept] )) OR "Lipodystrophy"[Mesh]) OR "Pancreatic Agenesis, Congenital" [Supplementary Concept]) OR "Pancreatic Hypoplasia, Congenital, with Diabetes Mellitus and Congenital Heart Disease" [Supplementary Concept]) OR "Donohue Syndrome"[Mesh]) OR "Mason-Type Diabetes" [Supplementary Concept]) OR "Lipodystrophy, Familial Partial"[Mesh]) OR "Renal cysts and diabetes syndrome" [Supplementary Concept]) OR ( "Diabetes Mellitus, Transient Neonatal, 2" [Supplementary Concept] OR "Diabetes Mellitus, Transient Neonatal, 3" [Supplementary Concept] OR "Diabetes Mellitus, Transient Neonatal, 1" [Supplementary Concept] OR "6q24-Related Transient Neonatal Diabetes Mellitus" [Supplementary Concept] )) OR "Noninsulin-dependent diabetes mellitus with deafness" [Supplementary Concept] Filters: English | **4731** |
| **#2**<br>"monogenic diabetes"[Title/Abstract] OR "neonatal diabetes"[Title/Abstract] OR "maturity onset diabetes of the young"[Title/Abstract] OR "maturity-onset diabetes of the young"[Title/Abstract] OR "infancy onset diabetes"[Title/Abstract] OR "infancy-onset diabetes"[Title/Abstract] OR "congenital diabetes"[Title/Abstract] OR TNDM OR PNDM* OR "early onset diabetes"[Title/Abstract] OR "early-onset diabetes"[Title/Abstract] OR "syndromic diabetes"[Title/Abstract] OR lipodystroph* OR "mitochondrial diabetes"[Title/Abstract] OR "pancreatic agenesis"[Title/Abstract] OR "pancreatic hypoplasia"[Title/Abstract] OR leprechaun* OR "donohue syndrome"[Title/Abstract] OR "rabson mendenhall"[Title/Abstract] OR rabson-mendenhall OR dunnigan OR "wolfram syndrome"[Title/Abstract] OR RCAD OR "renal cysts and diabetes"[Title/Abstract] OR "chromosome 17q12 deletion syndrome"[Title/Abstract] OR "chromosome-17q12 deletion syndrome"[Title/Abstract] OR "mason type diabetes"[Title/Abstract] OR "mason-type diabetes"[Title/Abstract] OR "transient neonatal diabetes"[Title/Abstract] OR "permanent neonatal diabetes"[Title/Abstract] OR maternally inherited diabetes and deafness OR "M.3243A>G mutation"[Title/Abstract] OR "mitochondrial tRNA"[Title/Abstract] OR "MODY" Filters: English | **14256** |
| **#3**<br>**#1 OR #2** | **14295** |

| | |
|---|---|
| **#4**<br>“Hepatocyte Nuclear Factor 1-beta"[Mesh] OR "Hepatocyte Nuclear Factor 1-alpha"[Mesh] OR "Hepatocyte Nuclear Factor 4"[Mesh] Filters: English | **3585** |
| **#5**<br>HNF1A OR HNF1-A OR "hepatocyte nuclear factor 1"[Title/Abstract] OR "hepatocyte nuclear factor 1a"[Title/Abstract] OR "hepatocyte nuclear factor 1 alpha"[Title/Abstract] OR "HNF1 alpha"[Title/Abstract] OR HNF1alpha OR HNF4A OR HNF4-A OR HNF-4A OR "hepatocyte nuclear factor 4"[Title/Abstract] OR "hepatocyte nuclear factor 4a"[Title/Abstract] OR "hepatocyte nuclear factor 4 alpha"[Title/Abstract] OR HNF4alpha OR GCK OR HNF1B OR HNF1-b OR HNF-1B OR "hepatocyte nuclear factor 1b"[Title/Abstract] OR "hepatocyte nuclear factor 1 beta"[Title/Abstract] OR HNF1beta OR "HNF-1 beta"[Title/Abstract] OR "INS gene"[Title/Abstract] OR "INS mutation"[Title/Abstract] OR KCNJ11[Title/Abstract] OR ABCC8[Title/Abstract] OR PDX1[Title/Abstract] OR CEL-MODY [Title/Abstract] OR "CEL gene"[Title/Abstract] OR "CEL mutation"[Title/Abstract] OR "caroboxyl ester lipase"[Title/Abstract] OR NEUROD1[Title/Abstract] OR WFS1[Title/Abstract] | **11142** |
| **#6**<br>(INSR[Title/Abstract]) AND (diabetes[Title/Abstract]) Filters: English | **273** |
| **#7**<br>(LMNA[Title/Abstract] OR PPARG[Title/Abstract] OR PLIN1[Title/Abstract]) AND (lipodystrophy[Title/Abstract]) Filters: English | **347** |
| **#8**<br>#4 OR #5 OR #6 OR #7 | **11988** |
| **#9**<br>#3 OR #8 | **23982** |
| **#10**<br>"Diagnosis"[Mesh] Filters: English | **7662064** |
| **#11**<br>"Phenotype"[Mesh] Filters:English | **307182** |
| **#12**<br>Neonatal Screening[Mesh] Filters:English | **9677** |
| **#13**<br>"etiology" [Subheading] Filters:English | **8144644** |

| | |
|---|---|
| **#14**<br>Biomarkers[MeSH] Filters:English | **760545** |
| **#15**<br>#10 OR #11 OR #12 OR #13 OR #14 | **7868628** |
| **#16**<br>diagnosis[Title/Abstract] Filters: English | **1367001** |
| **#17**<br>"clinical characterist*"[Title/Abstract] Filters: English | **72805** |
| **#18**<br>"identif*"[Title/Abstract] Filters:English | **3418989** |
| **#19**<br>phenotyp*[Title/Abstract] Filters: English | **613305** |
| **#20**<br>"risk score*"[Title/Abstract] Filters: English | **25785** |
| **#21**<br>"screening pathway*"[Title/Abstract] Filters: English | **127** |
| **#22**<br>etiolog*[Title/Abstract] Filters: English | **258874** |
| **#23**<br>"systematic assessment"[Title/Abstract] Filters: English | **2720** |
| **#24**<br>biomarker*[Title/Abstract] Filters: English | **320083** |
| **#25**<br>"clinical suspicion"[Title/Abstract] Filters: English | **12335** |
| **#26**<br>#16 OR #17 OR #18 OR #19 OR #20 OR #21 OR #22 OR #23 OR #24 OR #25 | **5290922** |

| | |
|---|---|
| **#27**<br>#15 OR #26 | **7868578** |
| **#28**<br>#9 AND #27 | **7487** |
| **#29**<br>#28 Filters: English, Humans | **6119** |
| 1990-2000: 626<br>2001-2010: 2138<br>2011-2021: 3025 | |
| **Question 2 How should you test someone for monogenic diabetes** | |
| **#30**<br>(((("High-Throughput Nucleotide Sequencing"[Mesh]) OR "Whole Exome Sequencing"[Mesh]) OR "Multiplex Polymerase Chain Reaction"[Mesh]) OR "Real-Time Polymerase Chain Reaction"[Mesh]) OR "Chromatography, High Pressure Liquid"[Mesh]) OR "Polymorphism, Restriction Fragment Length"[Mesh] Filters: Humans, English | **156758** |
| **#31**<br>"next generation sequencing"[Title/Abstract] OR "next-generation sequencing"[Title/Abstract] OR "sanger sequencing"[Title/Abstract] OR "dna sequencing"[Title/Abstract] OR "multigene panel"[Title/Abstract] OR "genetic testing"[Title/Abstract] OR "genetic screening"[Title/Abstract] OR "dna pooling"[Title/Abstract] OR "high-throughput sequencing"[Title/Abstract] OR "high throughput sequencing"[Title/Abstract] OR "targeted sequencing"[Title/Abstract] OR "target region capture sequencing"[Title/Abstract] OR "exome sequencing"[Title/Abstract] OR "genome sequencing"[Title/Abstract] OR mlpa OR "dosage analysis"[Title/Abstract] OR "cnv detection"[Title/Abstract] OR "cnv analysis"[Title/Abstract] OR "allele specific pcr"[Title/Abstract] OR "real-time pcr"[Title/Abstract] OR "fluorescent pcr"[Title/Abstract] OR taqman OR sscp OR hplc OR rflp OR "chip n3 genotyping"[Title/Abstract] OR "whole exome sequencing"[Title/Abstract] OR "multiplex ligation dependent probe amplification"[Title/Abstract] OR "allele specific polymerase chain reaction"[Title/Abstract] OR "real time polymerase chain reaction"[Title/Abstract] OR "single strand conformation polymorphism"[Title/Abstract] OR "high performance liquid chromatography"[Title/Abstract] OR "restriction fragment length polymorphism"[Title/Abstract] Filters: Humans, English<br>=304221 | **304221** |
| **#32**<br>#30 OR #31 | **335251** |
| **#33**<br>#9 AND #32 | **1460** |

| | |
|---|---|
| Paper per decade | 1990-2000: 114<br>2001-2010: 301<br>2011-2021: 1041 |
| **Question 6 What are the next steps after a diagnosis of monogenic diabetes** | |
| #91<br>("Genetic Counseling"[Mesh]) OR "Cell-Free Nucleic Acids"[Mesh] Filters: Humans, English | 16553 |
| #92<br>"cascade testing"[Title/Abstract] OR "cascade screening"[Title/Abstract] OR "genetic counselling"[Title/Abstract] OR "cell-free dna"[Title/Abstract] OR "cell free dna"[Title/Abstract] OR "noninvasive prenatal testing"[Title/Abstract] OR "non-invasive prenatal testing"[Title/Abstract] Filters: Humans, English | **8338** |
| #93<br>#91 OR #92 | 21814 |
| #94<br>#9 AND #93 | **61** |
| #95<br>"Penetrance"[Mesh] OR "Oceanic Ancestry Group"[Mesh] OR "American Native Continental Ancestry Group"[Mesh] OR "Asian Continental Ancestry Group"[Mesh] OR "European Continental Ancestry Group"[Mesh] OR "African Continental Ancestry Group"[Mesh] OR "Economic evaluation"[Mesh] OR "Continental Population Groups"[Mesh] Filters: Humans, English | **223571** |
| #96<br>variant[Title/Abstract]) AND (unknown[Title/Abstract] OR significan*[Title/Abstract] OR ancestry group[Title/Abstract] OR penetrance[Title/Abstract] OR economic evaluation[Title/Abstract]) Filters: Humans, English | **4866** |
| #97<br>#95 OR #96 | **2601031** |
| #98<br>#9 AND #97 | **844** |
| *Papers per decade* | *1990-2000: 87*<br>*2001-2010: 308*<br>*2011-2020: 492* |
| **Question 7 What are the current challenges for the field in precision diagnostics?** | |
| **#26**<br>'variant'/exp OR 'ancestry group'/exp OR 'penetrance'/exp OR 'economic evaluation'/exp | 992252 |
| **#27**<br>('variant of unknown significance':ab,ti OR 'variant n3':ab,ti) AND (unknown:ab,ti OR significan*:ab,ti) OR 'ancestry group':ab,ti OR penetrance:ab,ti OR 'economic evaluation':ab,ti | 33291 |

| | |
|---|---|
| **#28** #26 OR #27 | 992252 |
| Combined Search<br>#35 #7 AND #28<br>2230 records<br>AND [embase]/lim NOT ([embase]/lim AND [medline]/lim)<br>1004 records<br>Filter Human, English<br>859 records<br>NOT ‘conference abstract’:it | **201** |
| *Papers per decade* | *1990-2000: 4*<br>*2001-2010: 32*<br>*2011-2020:165* |

**Supplementary Table 2 PICOTS**

| | **Diagnostic validity** |
|---|---|
| **Population** | Patients with diabetes |
| **Intervention** | Genetic testing for monogenic diabetes (using at least sequencing of a single gene if not multiple genes or whole exome or whole genome) |
| **Comparison** | Unselected vs selected cases using various clinical or biomarker criteria to increase yield of monogenic diabetes |
| **Outcomes** | · Diagnostic yield (fraction and percentage)<br>· Sensitivity/specificity (if thresholded)<br>· AUROC (if not thresholded)<br>· PPV/NPV |
| **Timing** | n/a |
| **Setting** | n/a |

#### Supplementary Table 3: Complete set of papers extracted for question 1. Who to test for monogenic diabetes.

| Study Details | Study type, Country, Ancestry of Participants | n=Number of probands, M:F Males:Females BMI | Method and genes tested | Variant curation method | Characteristics of diabetes population tested (age of diagnosis, or other clinical or biomarker criteria) | Individuals diagnosed, yield by different selection approaches |
|---|---|---|---|---|---|---|
| **Neonatal Diabetes** | | | | | | |
| DeFranco (2015) [1] | Cohort study, International, referred to Exeter Molecular Genetics from 79 countries | n=1021 neonates with NDM, M:F 571:449, BMI NA | Type of genetic testing: Neonatal Diabetes Panel. Genes Sequenced: 6q24, ABCC8, EIF2AK3, FOXP3, GATA4, GATA6, GCK, GLIS3, HNF1B, IER3IP1, INS, KCNJ11, IER3IP1, MNX1, NEUROD1, NEUROG3, NKX2-2, PDX1, PTF1A, RFX6, SLC2A2, SLC19A2, ZFP57 | Bioinformatics method ALAMUT | Diagnosed with diabetes before 6 months of age | Overall Yield 840/1021 (82%) including 30 GCK, 1 HNF1B, 150 ABCC8, 240 KCNJ11, 110 INS, 1 RFX6, 3 NEUROD1, 6 PDX1, 76 EIF2AK3, 22 PTF1A, 14 FOXP3, 113 6q24, 7 SLC19A2, 29 GATA6, 4 GATA4, 9 GLIS3, 1 IER3IP1, 1 MNX1, 1 NEUROG3, 2 NKX2-2, 6 SLC2A2, 12 ZFP57 |
| Besser (2016) [2] | Cohort study, United Kingdom, | n=750, M:F 333:417, BMI NA | Type of genetic testing: Neonatal diabetes panel. Genes sequenced: GCK; HNF1B; KCNJ1, ABCC8, INS (all patients). 6q24, EIF2AK3, NEUROD1, RFX6, GCK, FOXP3, SLC19A2, GLIS3, PDX1, PTF1A, NEUROG3, HNF1B, BSCL2 (if clinically indicated). | Local lab guidelines | Patients with neonatal diabetes (hyperglycemia presenting <26 weeks of age) who were referred for genetic testing and who had a gestational age provided. | Neonatal DM (diabetes diagnosed <26 weeks): 598/750 (79.7%);<br>-Preterm (<37 wk) with neonatal DM: 66% (97/146);<br>-Term (>=37 wk) with neonatal DM: 83% (501/604);<br>-Very preterm (<32 wk) with neonatal DM: 31% (13/42) |
| Habeb (2012) [3] | Cross sectional study, United Kingdom, Arab and British | n=165, M:F Arab 60:28; British 39:38, BMI NA | Type of genetic testing: Single gene(s). Genes sequenced: KCNJ11, ABCC8, INS (all), EIF2AK3, GCK, FOXP3, GLIS3, SLC19A2, PTF1A, NEUROD1, RFX6, SLC2A2 (based on phenotype) | None specified | 88 Arab and 77 British probands with neonatal diabetes | Arab group: 56/88; British group: 32/77 |

| | | | | | | |
|---|---|---|---|---|---|---|
| Gopi (2021) [4] | Cohort study, India, South East Asian - Indian | n=181, M:F NA, BMI NA | Type of genetic testing: Single gene(s). Genes sequenced: ABCC8, KCNJ11 | ACMG | Neonatal diabetes diagnosed up to 6 months | Diabetes diagnosed before 6 months: 39/181 |
| Edghill (2008) [5] | Cohort study, United Kingdom, Multiple ethnicities - international cohort | n=1044, M:F NA, BMI NA | Type of genetic testing: Single gene(s). Genes sequenced: INS | Local lab guidelines | Neonatal diabetes diagnosed up to 2 years and young adults diagnosed up to 25 years from the Barts and Oxford (BOX) study and the British Diabetes Association 1972–1981 cohort. | <6 months, insulin treated, KCNJ11 negative: 35/285<br>-ND diagnosed <6 months yield: 33/141<br>-ND diagnosed 6-12 months yield: 2/86<br>-ND diagnosed >12 months yield: 0/58<br>-MODY yield in probands with clinically diagnosed MODY: 1/296<br>-Mutation rate found in type 2 diabetes diagnosed<45: 1/463 |
| DeFranco (2013) [6] | Cross sectional study, United Kingdom, Multiple ethnicities - international cohort | n=103, M:F NA, BMI NA | Type of genetic testing: Single gene(s). Genes sequenced: PDX1 | None specified | Probands with permanent neonatal diabetes diagnosed before 6 months | Diabetes <6 months, mutations in ABCC8, KCNJ11 and INS (and EIF2AK3 in consanguineous) had been excluded, None of the patients had pancreatic agenesis or exocrine insufficiency: 3/103 |
| Shaw-Smith (2012) [7] | Cohort study, United Kingdom, Multiple ethnicities - internationally cohort | n=212, M:F NA, BMI NA | Type of genetic testing: Single gene(s). Genes sequenced: SLC19A2 | None specified | International probands with permanent neonatal diabetes diagnosed up to 6 months | Total yield index (dx <6 months) and extended (dx>6 months): 5/400<br>-ND diagnosed <6 months of unknown genetic cause: 3/212;<br>-ND diagnosed >6 months: 2/188 |
| Borowiec (2012) [8] | Cross sectional study, Poland, Polish | n=211, M:F NA, BMI NA | Type of genetic testing: Single gene(s). Genes sequenced: GCK | None specified | Either diabetes diagnosed before 6 months or 2 or more clinical criteria of MODY diagnosed up to 35 years | Neonatal Diabetes and negative KCNJ11, ABCC8, INS: 1/17 homozygous GCK;<br>>2 MODY criteria: 68/194;<br>Negative for GCK: 0/126 |

| | | | | | | |
|---|---|---|---|---|---|---|
| Rubio-Cabezas (2012) [9] | Cross sectional study, Spain, | n=520, M:F NA, BMI NA | Type of genetic testing: Neonatal diabetes panel. Genes sequenced: GCK, KCNJ11, INS, ABCC8 | None specified | Infants with diabetes diagnosed < 12 months old | Total age 0-52 weeks: 272/550 (49.5%); KCNJ11: 144/550 (26%); ABCC8: 56/520 (11%); INS: 64/550 (12%); GCK 8/58 (screened after the others if no findings)<br>-ND diagnosed <6 months yield: 263/405<br>-ND diagnosed 6-12 months yield: 9/145 |
| Busiah (2013) [10] | Cohort study, France, Multiple Ethnicities, 20 countries | n=174, M:F NA, BMI NA | Type of genetic testing: Neonatal diabetes panel. Genes sequenced: 6q24, INS, KCNJ11, ABCC8 | None specified | French Neonatal Diabetes Study Group - neonatal diabetes diagnosed up to 12 months | All unrelated cohort participants: 127/174;<br>-6q24: 40/174; KATP (KCNJ11/ABCC8): 74/174; INS: 13/174<br>-ND diagnosed <6 months yield among all diagnosed <12m: 64/155<br>-ND diagnosed 6-12 months yield among all diagnosed <12m: 5/18 |
| Gopi (2021) [11] | Cohort study, India, South East Asian - Indian | n=189, M:F NA, BMI NA | Type of genetic testing: Single gene(s). Genes sequenced: INS | ACMG | Antibody negative children with neonatal diabetes diagnosed up to 9 months from India. | <9 months, persists >12 months of age, antibody negative, C-peptide detectable: 8/189<br>-ND diagnosed <6 months of total group diagnosed <9 months: 8/189<br>-ND diagnosed 6-9 months of total group diagnosed <9 months: 0/189 |
| Flanagan (2006) [12] | Cross sectional study, United Kingdom, Multiple ethnicities - international cohort | n=239, M:F 129:110, BMI NA | Type of genetic testing: Single gene(s). Genes sequenced: KCNJ11 | None specified | Permanent diabetes diagnosed before 2 years of age | Total cohort ND yield: 31/239<br>-ND diagnosed <6 months: 31/120<br>-ND diagnosed 6-12 months: 0/50<br>-ND diagnosed 12-24 months: 0/70 |

| | | | | | | |
|---|---|---|---|---|---|---|
| Edghill (2006) [13] | Case control study, United Kingdom, Multiple ethnicities - international cohort | n=187, M:F NA, BMI NA | Type of genetic testing: Single gene(s). Genes sequenced: KCNJ11 | None specified | Recruited worldwide with diabetes diagnosed before 2 years of age | Permanent diabetes diagnosed under the age of 2 years: 32/187;<br>-ND diagnosed <6 months: 32/79<br>-ND diagnosed 6-12 months: 0/45<br>-ND diagnosed 12-24 months: 0/63<br>-KCNJ11 positive, diagnosed with diabetes under the age of 6 months (n=32): 3% had highest risk HLA genotype, 16% protective genotypes, similar to healthy controls (3% and 17%);<br>-KCNJ11 negative, diagnosed with diabetes <6months (n=47): 9% had highest HLA genotype, 17% protective genotypes |
| **Gestational Diabetes: European Ancestry Predominant** | | | | | | |
| Chakera (2014) [14] | Cohort study, UK and Ireland, 93% White European | n=356, M:F 0:400, BMI NA | Type of genetic testing: Single gene(s). Genes sequenced: GCK | Local lab guidelines | Atlantic Diabetes in Pregnancy (Atlantic DIP) cohort in which all women with GDM, and a random group without GDM, had GCK sequencing. Characteristics of women diagnosed with GCK-MODY during pregnancy were also analyzed. | Yield of GCK cases per total with genetic testing: 4/356<br>-GDM with fasting glucose <5.1 mmol/L: 0/109<br>-GDM with fasting glucose 5.1-5.5 mmol/L: 1/129<br>-GDM with fasting glucose >=5.5 mmol/L: 3/118 |
| Gjesing (2017) [15] | Cohort study, Denmark, Danish | n=354, M:F 0:354, BMI 27.6 (23.9-32.9) | Type of genetic testing: Large MODY panel (>= 5 genes). Genes sequenced: HNF1A; GCK; HNF4A; HNF1B; INS | ACMG | Antibody negative women with diet-treated GDM recruited in Denmark | Diet-treated GDM: 21/354 |
| Zubkova (2019) [16] | Cohort study, Russia, Russian | n=188, M:F 0:188, BMI NA | Type of genetic testing: Large MODY panel (>= 5 genes). Genes sequenced: 28 gene panel | ACMG | Russian pregnant non-obese women with diabetes during pregnancy | GDM, non-obese: 59/188 |
| **Gestational diabetes: Non-European** | | | | | | |

| | | | | | | |
|---|---|---|---|---|---|---|
| Jiang (2022) [17] | Case control study, China, China, Shanghai | n=835 including 400 controls, M:F 229: 208 , BMI GDM: 22.7±3.4, GCK-MODY: 20.2±2.1) | Type of genetic testing: Single gene(s). Genes sequenced: GCK | GnomADFunctional studies | Women with GDM and controls | GDM: 15/411;<br>Affected Fathers: 6/24;<br>Health Controls: 0/400 |
| **GCK: European Ancestry predominant** | | | | | | |
| Valentínová (2012) [18] | Cohort study, Slovakia, | n=100, M:F NA, BMI NA | Type of genetic testing: Single gene(s). Genes sequenced: GCK | None specified | Individuals diagnosed up to age 35 years with a suspected diagnosis of GCK-MODY | Persistent and stable fasting hyperglycemia (5.5–10.0 mmol/L), HbA1c <8% [64 mmol/ml], age of diagnosis <35years, and when on insulin treatment, dose <0.4 IU/Kg/day: 36/100 |
| Sagen (2008) [19] | Cohort study, Norway, Northern European | n=112, M:F NA, BMI GCK 22.3(+/-5.0) No GCK mutation 25.1(+/-4.7) | Type of genetic testing: Single gene(s). Genes sequenced: GCK | HGDM | Cases suspected to have MODY diagnosed up to 25 years referred to a diagnostic centre | Suspected MODY (initial referral): 23/329; Suspected MODY sequenced for GCK: 23/112 |
| Pinelli (2013) [20] | Cohort study, Italy, Italian | n=232, M:F NA, BMI BMI Z-score close to zero | Type of genetic testing: Single gene(s). Genes sequenced: GCK | None specified | Used known GCK-MODY cases to devise diagnostic criteria composed of seven clinical, biochemical and anamnestic criteria for case finding of GCK-MODY prospectively in those aged 6 months-25 years. | Validated criteria in two retrospective cohorts and applied to prospective cohort of 921 patients of which 21 fulfilled 7 criteria and were tested with yield of 13/21. |
| Massa (2001) [21] | Cohort study, Italy, Italian | n=131, M:F NA, BMI NA | Type of genetic testing: Single gene(s). Genes sequenced: GCK | Local lab guidelines | Children with suspected GCK-MODY based on negative islet antibodies, family history and stable fasting hyperglycaemia | MODY pedigree index patients: 54/132;<br>Chronic fasting hyperglycaemia, negative for serological markers of Type I<br>insulin-dependent) diabetes |

| | | | | | | |
|---|---|---|---|---|---|---|
| | | | | | | mellitus and glucose-tolerant parents: 3/9 |
| Borowiec (2012) [22] | Intervention (campaign on MODY)., Poland, Polish | n=178, M:F NA, BMI NA | Type of genetic testing: Single gene(s). Genes sequenced: GCK | Checked against Human Gene Mutation Database | Cases referred with clinical suspicion of GCK-MODY following a public information campaign. | Yield 54/178 (30%) |
| Aykut (2018) [23] | Cohort study, Turkey, Turkish | n=177, M:F 85:92, BMI NA | Type of genetic testing: Single gene(s). Genes sequenced: GCK | Local lab guidelines | Clinically suspected MODY referred from 2009-2015. Diabetes diagnosed up to 50 years with a family history of diabetes and absence of islet autoantibodies. | Yield 79/177 (45%) |
| Aloi (2017) [24] | Cohort study, Italy, Italian | n=100, M:F 56:44, BMI Non-obese | Type of genetic testing: Single gene(s). Genes sequenced: GCK | Local lab guidelines | Impaired fasting glycaemia diagnosed 6 months to 56 years recruited from a single clinic 2009-2015.<br>Group 1 "strict criteria": age of onset <25 yrs, FPG >99mg/dl, OGTT 2 hr 140mg/dl, absence of islet antibodies, family history, absence of obesity and no treatment.<br>Group 2: IFG and absence of islet antibodies + at least one other MODY criteria. | Group 1 32 cases out of 36 (88%)<br>Group 2 21 cases out of 64 (32%)<br>Overall yield 53% |

| | | | | | | |
|---|---|---|---|---|---|---|
| Lukášová (2008) [25] | Case control study, Czech Republic, Czech | n=722, M:F 210:490, BMI T2D: 30.53 +/- 5.53, GDM: 26.41 +/-5.03, Healthy offspring of DM: 25.51 + 4.16, Controls: 23.13 +/- 3.38 | Type of genetic testing: Single gene(s). Genes sequenced: GCK | None specified | Clinically suspected MODY in cases of T2D, GDM and non-diabetic, with and without family history of diabetes | 12 MODY probands: 2/12; GDM, T2D, offspring, controls: 0/710 |
| Pace (2022) [26] | Cohort study, Malta, Maltese | n=145, M:F 59:86, BMI IFG: Median 27.6/IQR2.5, Diabetes:Median28.5/IQR 2.5 | Type of genetic testing: Single gene(s). Genes sequenced: GCK | ACMG | Adults with suspected MODY diagnosed up to age 40 years from a single centre between 2018-2019. | All (IFG + Diabetes): 3/145; -Subgroup (IFG): 3/104 |
| Vits (2006) [27] | Cohort study, Belgium, North European | n=124, M:F NA, BMI NA | Type of genetic testing: Single gene(s). Genes sequenced: GCK | Local lab guidelines | 124 families, referred to a tertiary centre between 2002 and2005. All probands fulfilled at least two of the following criteria: early-onset hyperglycaemia (age of onset up to 40 years), the absence of islet autoantibodies and a positive familial history for diabetes | Hyperglycemia age < 40, autoantibody negative, positive family history with at least 2 generations affected: 33/124 |
| Thomson (2003) [28] | Cohort study, United Kingdom, | n=212, M:F NA, BMI NA | Type of genetic testing: Single gene(s). Genes sequenced: GCK | Local lab guidelines | Suspected MODY, recruited for research or diagnostic referral | Research testing 29/86 (34%); Diagnostic testing 32/126 (25%) |
| **GCK: Non-European** | | | | | | |

| | | | | | | |
|---|---|---|---|---|---|---|
| Ma (2019) [29] | Cohort study, Other: China, Chinese | n=679, M:F NA, BMI NA | Type of genetic testing: Genes sequenced: GCK | ACMG | pre-diabetes, newly diagnosed diabetes and existing diabetes | GCK-MODY in Discovery cohort 11/545 (2%), Replication cohort 3/134 (2%), Test cohort using criteria fasting glucose 5.4-8.3 and TG<1.43: 1/207 (0.5%) |
| Carmody (2016) [30] | Cohort study, United States, Multiple ethnicities | n=135, M:F NA, BMI NA | Type of genetic testing: Single gene(s). Genes sequenced: GCK | Local lab guidelines | Probands from the US Monogenic Diabetes Registry with GCK-MODY phenotype had sequencing. Additional individuals with known GCK-MODY were also included in the study | Initial Criteria as follows: Chronic mild fasting hyperglycemia (100–140 mg/dl, 5.55–7.75 mmol/l) and HbA1c 5.6–7.8 % (38–62 mmol/mol) plus either a linear three-generation family history of hyperglycemia or diabetes mellitus or BMI less than 30 kg/m2 in adults and BMI below the 95th percentile in children and age at diagnosis less than 30 years: 64/117; Initial criteria + negative autoantibodies: 3/7; Initial criteria + Latin ancestry: 4/8; Initial Criteria + Asian ancestry: 4/9 |
| **Monogenic diabetes: European ancestry predominant** | | | | | | |
| Ivanoshchuk (2021) [31] | Cohort study, Russian, Western Siberia | n=178, M:F NA, BMI NA | Type of genetic testing: Large MODY panel (>= 5 genes). Genes sequenced: HNF4A,GCK,HNF1A,PDX1,HNF1B,NEUROD1,KLF11,CEL,PAX4,INS,BLK,KCNJ11,ABCC8,APPL1 | ACMG | Probands with diabetes diagnosed up to 35 years and clinically suspected MODY | <35 years, family history of diabetes, without obesity, islet autoantibody negative, normal or mildly reduced c-peptide, no insulin therapy, absence of diabetic ketoacidosis: 38/178 |
| Saint-Martin (2022) [32] | Cross sectional study, France, Eurocaucasian | n=1676, M:F 771:905, BMI 23.5[20.5-27.2] | Type of genetic testing: Large MODY panel (>= 5 genes). Genes sequenced: 18 gene panel | Monogenic Diabetes Expert Panel (ClinVar) rules, adapted from the ACMG guidelines | Probands referred for MODY genetic testing in France | Probands referred for MODY testing between 2017 and 2020: 307/1676 |

| | | | | | | |
|---|---|---|---|---|---|---|
| Colclough (2022) [33] | Cohort study, United Kingdom, Multiple ethnicity | n=1280, M:F 556:724, BMI 25.7(22.4–30.0) | Type of genetic testing: Large MODY panel (>= 5 genes). Genes sequenced: HNF1A; GCK; HNF4A; HNF1B; LMNA; ABCC8, CEL, CISD2, GATA4, GATA6, INS, INSR, KCNJ11, MTTL1 m.3243A>G, NEUROD1, PAX6, PCBD1, PDX1, PLIN1, POLD1, PPARG, RFX6, SLC29A3, TRMT10A, WFS1, ZBTB20, ZFP57 | ACMG | All referrals from U.K. clinicians for routine MODY genetic testing 2011-2018 | Suspected MODY 297/1280; -MODY confirmed cases who had syndromic gene: 56/297; m.324A>G 24/56, HNF1B 18/56, 6 other genes (WFS1, INSR, GATA6, SLC29A3, TRMT10A, PPARG) 14/56 -Patients with syndromic variant with clinical features of syndrome without diabetes: 11/56 |
| Shields (2017) [34] | Cross sectional study, United Kingdom, Multiple ethnicity (98% white) | n=216, M:F NA, BMI NA | Type of genetic testing: Large MODY panel (>= 5 genes). Genes sequenced: HNF1A; GCK; HNF4A; HNF1B; LMNA; Mitochondrial genome | None specified | People with diabetes diagnosed <30 years of age, currently younger than 50 years, in two U.K. regions with existing high detection of monogenic diabetes (UNITED study). | UCPCR positive or Non-insulin treated with negative autoantibodies for IA2 and GAD: 51/216 |
| Karaoglan (2021) [35] | Cross sectional study, Turkey, Turkish | n=230, M:F 113:117, BMI NA | Type of genetic testing: Large MODY panel (>= 5 genes). Genes sequenced: HNF1A; GCK; HNF4A; HNF1B; LMNA; ABCC8, GATA6, INS, KCNJ11, NEUROD1, PAX6, PDX1, PPARG, INSR | None specified | Diagnosed up to 18 years with atypical presentation for type 1 and type 2 diabetes | Full cohort with MODY genetic variant: 24/230 including GCK (15), HNF1A (1), HNF4A (7), PDX1(1); -Lower total daily insulin dose: 20 MODY genetic variant/122; -Positive islet antibody: 4 MODY genetic variant/24 |

| | | | | | | |
|---|---|---|---|---|---|---|
| Bjørkhaug (2003) [36] | Cohort study, Norway, Norwegian | n=130, M:F NA, BMI NA | Type of genetic testing: Single gene(s). Genes sequenced: HNF1A | Local lab guidelines | Families recruited from the Norwegian MODY Registry with either clinical MODY (fulfilled strict criteria including autosomal dominant inheritance in two generations, at least two relatives of the proband having either diabetes or impaired glucose tolerance, onset of diabetes before 25 yr of age in at least one subject, and reduced insulin secretion), suspected MODY (provider suspicion, limited clinical data), or pedigrees with multiplex type 1 diabetes. | Clinical MODY 22/42 (52%); Suspected MODY 15/75 (20%), Multiplex T1D 1/13 (7.7%) |
| Søvik (2013) [37] | Cross sectional study, Norway, Norwegian | n=not supplied, registry contains 198 families with MODY , M:F NA, BMI NA | Type of genetic testing: Single gene(s). Genes sequenced: HNF1A,GCK,HNF4A,HNF1B, CEL, INS, KCNJ11, ABCC8, NEUROD1 | None specified | The Norwegian MODY registry, diagnosed up to 40 years | 1500 tested referred cases: 458/1500; HNF1A (208/105 families)/ GCK (138/59 families); HNF4A (40/15 families)/HNF4B (19/11 families)/CEL (39/2 families), INS (9/4 families), NEUROD1 (4/2 families) |
| Frayling (2001) [38] | Cross sectional study, United Kingdom, 97% UK Caucasian | n=210, M:F , BMI NA | Type of genetic testing: Large MODY panel (>= 5 genes). Genes sequenced: HNF1A; GCK; HNF4A; HNF1B; IPF-1, NEUROD1 | None specified | Type 2 diabetes diagnosed up to age 35 with clinical suspicion of MODY (specific MODY criteria fulfilled) | Full cohort (screened for frameshift mutation p.291fsinsC/p.291fsdelC): 24/210; MODY panel screening 101 sequenced: HNF1A 34, GCK 22, HNF4A 2, HNF1B 4 |

| | | | | | | |
|---|---|---|---|---|---|---|
| Johansson (2017) [39] | Case control study, Norway, European | n=938, M:F 554:384, BMI NA | Type of genetic testing: Large MODY panel (>= 5 genes). Genes sequenced: HNF1A,GCK,HNF4A,HNF1B, INS,ABCC8,KCNJ11,BLK,CEL,NEUROD1,KLF11,PAX4,PDX1. | Score system as defined by Plon et al Hum Mut 2018 | Norwegian Childhood Diabetes Registry diagnosed up to age 15, absence of islet antibodies. Genetic testing reported with and without class 3 variants (VUS) | Diabetes <15 years, antibody negative, class 3-5 variants: 30/462;<br>Diabetes <15 years, antibody positive, class 3-5 variants: 11/468;<br>Diabetes <15 years, antibody negative, class 4-5 variants: 19/462;<br>Diabetes <15 years, antibody positive, class 4-5 variants: 1/468. |
| Donath (2019) [40] | Cross sectional study, France, 60% Eurocaucasian | n=1564, M:F 737:827, BMI NA | Type of genetic testing: Large MODY panel (>= 5 genes). Genes sequenced: GCK,HNF1A,HNF4A,HNF1B, ABCC8,KCNJ11,INS | ACMG | Clinically suspected MODY from 116 Endocrinology departments, Islet antibody negative | At least 2 of a) age of onset 15-40 years; b) BMI < 30 kg/m2 and c) at least 2 gen FH: 254/1564 (16%);<br>Age of onset 15-40 years AND BMI < 30 kg/m2 and AND at least 2 gen FH: 198/999 (20%);<br>Age of onset 15-40 years AND BMI < 30 kg/m2 and AND at least 2 gen FH AND EuroCaucasian origin: 54/504 (11%);<br>At least 2 gen FH, age at onset ≤40 years, and BMI < 25 kg/m: 101/301 (34%) |

| | | | | | | |
|---|---|---|---|---|---|---|
| Bansal (2017) [41] | Case control study, Germany, Northern European | n=6888, M:F Stage 1 Cases: 1042:838 Stage 2 Cases: 1333:803 Stage 1 Controls: 852:988 Stage 2 Controls: NA, BMI Stage1 Cases:27.3 Stage2 Cases: 27.8 Stage1 Controls:25.6 Stage2Controls: 26.6 | Type of genetic testing: Large MODY panel (>= 5 genes). Genes sequenced: HNF1A,HNF4A,HNF1B,INS, NEUROD1,PDX1,PAX4,ABCC8,KCNJ11,KLF11,CEL,BLK, WFS1,NEUROG3,EIF2AK3, GLIS3,RFX6,SLC19A2,PAX, GATA6, PPARG | ACMG | Islet antibody negative with no clinical features of T1D recruited at tertiary centre. Not selected for typical MODY features and mean age >40years. | Young onset and late onset type 2 diabetes: 47/4016; Young onset (<40) diabetes: 29/1346; Older onset (>=40) diabetes: 18/2670 (all the yields are for GCK, HNF1A, HNF4A, HNF1B, ABCC8 only) |
| Ateş (2021) [42] | Cohort study, Turkey, Turkish | n=182, M:F 94:88, BMI | Type of genetic testing: Large MODY panel (>= 5 genes). Genes sequenced: HNF1A; GCK; HNF4A; HNF1B; KCNJ11, ABCC8, INS | ACMG | Clinical diagnosis of MODY, diabetes diagnosed below 35, and family history of diabetes. | Clinical diagnosis of MODY diagnosed with diabetes before the age of 35: 30/182 |
| Bennett (2015) [43] | Cohort study, United States, North America | n=184, M:F 47:53, BMI NA | Type of genetic testing: Single gene(s) (specify below). Genes sequenced: HNF1A; GCK; HNF4A; Other: INS, ABCC8, KCNJ11 | Local lab guidelines | Patients with neonatal diabetes or diabetes suspected to be MODY referred to reference diagnostic laboratory | Neonatal diabetes or suspected MODY: 51/184 Subset number of ND vs MODY NA (1/3 of population had no phenotype data) |

| | | | | | | |
|---|---|---|---|---|---|---|
| Irgens (2013) [44] | Cross sectional study, Norway, Norwegian | n=118, M:F NA, BMI NA | Type of genetic testing: Small MODY panel (<5 genes): KCNJ11, ABCC7, INS (PNDM); HNF1A; HNF4A, INS, MT3243A>G (MODY A); or GCK (MODY B). | None specified | Norwegian Paediatric diabetes Registry, diagnosed aged 0-14 years, includes 91% of children with diabetes. | PNDM Group: 4/24 (all KCNJ11, diagnosed <6m); MODY A (Autoantibody - and diabetes in parent): 6/46 (5 HNF1A, 1 INS)<br>MODY B (A1c <7.4%, no insulin): 4/98 (3 GCK, 1HNF1A)<br>Total estimated Norwegian prevalence (plus 30 extra from MODY registry): 48/323 autoantibody negative and patients not found in the NCDR registry of Norwegian children (15 patients) |
| Stankute (2020) [45] | Cross sectional study, Lithuania, Lithuanian | n=153, M:F 80:73, BMI NA | Type of genetic testing: Large MODY panel. Genes sequenced: 307 gene panel including 14 MODY and 28 NDM genes | None specified | LithuanianSwiss project, Lithuanian pediatric diabetes registry with absence of islet autoantibodies. | Yield 42/153 (27%) |
| Vaxillaire (2021) [46] | Cross sectional study, France , Multiple ethnicities | n=204, M:F 91:54, BMI NA | Type of genetic testing: Large MODY panel. Genes sequenced: 35 gene panel | ACMG | Unrelated individuals from 11 Mediterranean countries with a clinical suspicion of MODY | Overall yield 36/204 (17.6%) |
| Schober (2009) [47] | Cross sectional study, Germany and Austria, | n=272, M:F NA, BMI NA | Type of genetic testing: Small MODY panel (<5 genes). Genes sequenced: HNF1A; GCK; HNF4A | None specified | Diagnosed up to 18 years, clinically defined as MODY, based on family history and clinical presentation. 272 had diagnostic genetic testing through routine labs. | 263/272 (97%) confirmed as MODY. |
| Misra (2016) [48] | Cohort study, United Kingdom, South Asian and European | n=4010, M:F NA, BMI NA | Type of genetic testing: Small MODY panel (<5 genes). Genes sequenced: HNF1A; GCK; HNF4A; HNF1B | According to Wallis, et al, 2013. ACGS Practice guidelines | People referred for MODY testing at National centre whose ancestry was known | Individuals referred for MODY testing: 1011/3472;<br>South Asian individuals referred for MODY testing: 37/293 |

| | | | | | | |
|---|---|---|---|---|---|---|
| Carlsson (2020) [49] | Cohort study, Sweden, Swedish | n=485, M:F 2178:1755, BMI Z score - 0.35 (1.55) | Type of genetic testing: Small MODY panel (<5 genes). Genes sequenced: HNF1A; GCK; HNF4A | ACMG | Paediatric probands newly diagnosed with diabetes between 2005-2010 for who there was an islet antibody result | Diagnosis 1-18 yrs, antibody negative x4: 46/303; Diagnosis 1-18 yrs, antibody positive: 0/182: Diagnosis 1-18 yrs, antibody negative, HbA1c <7.5% at dx or family history: 44/131; Diagnosis 1-18 yrs, antibody negative, positive family history: 29/96 |
| Stanik (2014) [50] | Cohort study, Slovakia, Slovakian and Czech | n=150, M:F NA, BMI NA | Type of genetic testing: Single gene(s). Genes sequenced: HNF1A; GCK; HNF4A | None specified | Probands diagnosed up to age 25 clinically suspected to be MODY but without family history of diabetes. | Patients fulfilling inclusion criteria: Total 85/150 including GCK 59, HNF1A 13, HNF4A 1 |
| Bellanné-Chantelot (2016) [51] | Case control study, France, French | n=453, M:F NA, BMI NA | Type of genetic testing: Single gene(s). Genes sequenced: HNF1A | None specified | Diagnosed up to 40 years, islet antibody absent with (1) Suspected HNF1A–MODY, (2) Familial young T2D (YT2D) (3) General T2D. Clinical model and use of hsCRP tested | Clinical model: HNF1A vs T2D, ROC C-statistic 0.99; HNF1A vs YT2D ROC C-statistic 0.82 Clinical model + CRP, HNF1A vs T2D ROC C-statistic 0.99, HNF1A vs YT2D ROC C-statistic 0.85 |
| Pihoker (2013) [52] | Cross sectional study, United States, 36% Non-Hispanic white, 20 % African American, 31%hispanic, 11 % Asian/Pacific Islander | n=586, M:F 239:347, BMI NA | Type of genetic testing: Sanger sequencing. Genes sequenced: HNF1A; GCK; HNF4A | Local lab guidelines | Diabetes <20 yrs from the SEARCH study | All MODY 47/586 (8% of auto-antibody negative, C-peptide positive) |

| | | | | | | |
|---|---|---|---|---|---|---|
| Lehto (1999) [53] | Cohort study, Sweden, Swedish, Finnish | n=115, M:F 55:60, BMI 26.5 (5.5) | Type of genetic testing: Single gene(s). Genes sequenced: HNF1A; GCK; HNF4A; Mitochondrial genome | Local lab guidelines | Families with early-onset diabetes (up to age 40) and strong family history of diabetes | Total cohort 12/115 including HNF4A 2, GCK 4, HNF1A 3, MIDD 3 |
| Riveline (2012) [54] | Cross sectional study, France, | n=146, M:F NA, BMI NA | Type of genetic testing: Single gene(s). Genes sequenced: ABCC8 | None specified | Family members or adults with ABCC8 mutations plus 139 adults with diabetes well controlled on SU agents. | Patients with affected family member: 7/7<br>T2D controlled on sulfonylurea: 4/139 |
| Delvecchio (2017) [55] | Cross sectional study, Italy, 93% white European, 3.4 % North African, 1.3 % African | n=240 already diagnosed monogenic diabetes, M:F NA, BMI NA | Type of genetic testing: Sanger sequencing. Genes sequenced: HNF1A; GCK; HNF4A; HNF1B; Screened first gene based on clinical suspicion. First GCK if IFG or stable fasting hyperglycemia Max 8,3mol/L AND neg autoantibodies. If negative > HNF1A if negative >HNF4A. Severe progressive >HNF1A first. Urogenital malformations > HNF1B. PDX1, NEUROD1, INS, ABCC8, KCNJ11. Investigated last > GATA6 and INSR | Checked against Human Gene Mutation Database | Children consecutively diagnosed with diabetes or impaired fasting glucose from 2007-2012 | Monogenic diabetes represented 240/3781 of the cohort including MODY cases: GCK 181, HNF1A 16, HNF4A 6, HNF1B 3; PDX1 2, INS 1, ABCC8 1; Neonatal diabetes 21 cases, genetic syndromes 9 cases |
| Thanabalasingham (2012) [56] | Cross sectional study, United Kingdom, 90% white British | n= 140, M:F NA, BMI NA | Type of genetic testing: Sanger sequencing. Genes sequenced: HNF1A; GCK; HNF4A | None specified | Young Diabetes in Oxford Study: diagnosed up to 45 years and investigated if met certain clinical criteria for MODY. | HNF1A-MODY was found in 2/20 patients with Clinically Defined T1D with evidence of residual beta-cell function, 13/80 patients with clinically defined T2D with either onset<30years or absence of metabolic syndrome |

| | | | | | | |
|---|---|---|---|---|---|---|
| Lorini (2009) [57] | Cohort study, Other: Italy, Italian | n=172, M:F , BMI GCK 17(14-24);HNF1A 21 (13-25);cases without GCK or HNF1A 17(16-20) | Type of genetic testing: Single gene(s). Genes sequenced: HNF1A; GCK | Local lab guidelines | Multicenter study on incidental hyperglycemia, diagnosed up to age 25 | GCK 109/172<br>HNF1A 12/172 |
| Pavić (2018) [58] | Cross sectional study, Croatia, Croatian | n=477, M:F NA, BMI NA | Type of genetic testing: Single gene(s). Genes sequenced: HNF1A | Local lab guidelines | Adults diagnosed up to 45 years, HNF1A sequencing in all antibody negative, C-peptide positive individuals. Strict pathogenicity criteria. | Yield 8/477 (1.7%). Estimated prevalence of HNF1A-MODY in Croatia of 66 per million. |
| Juszczak (2019) [59] | Cohort study, United Kingdom, UK and Croatia | n=989, M:F 587:402, BMI HNF1A-MODY 25.5+/-9.4 : no variant 30.4+/-8.3 | Type of genetic testing: Single gene(s). Genes sequenced: HNF1A | ACMG | Young Diabetes in Oxford Study: diagnosis up to 45 years, fasting C-peptide >= 0.2 nmol/L, negative GAD antibodies. CRP and plasma glycans assessed as biomarkers. | All probands: 16/989 (1.6%); Using CRP<0.81, 14/269 (5%), sensitivity 88%, specificity 69%; Strict MODY criteria 8/99 (8%), sensitivity 50%; MODY calculator probability>20%: 9/136 (6.6%), sensitivity 56% |
| Demus (2022) [60] | Case control study, Croatia (n=501) and UK (n=499), UK and Croatia | n=947, M:F 540:407, BMI NA | Type of genetic testing: Single gene(s). Genes sequenced: HNF1A | ACMG | Diabetes diagnosed up to 45 years, islet antibody negative and C-peptide positive, all HNF1A sequenced and variants classified and tested against a plasma glycan biomarker plate assay constructed for the glycan pattern associated with HNF1A-MODY | Likely damaging variant vs no HNF1A variant: ROC curve C statistic 0.87 |
| Tatsi (2013) [61] | Cohort study, Greece, Greek | n=395, M:F NA, BMI NA | Type of genetic testing: Single gene(s). Genes sequenced: HNF1A | Bioinformatics | Clinically suspected MODY diagnosed up to 25 years, referred for diagnostic testing | Overall yield 88/395 (22%) |

| | | | | | | |
|---|---|---|---|---|---|---|
| Szopa (2016) [62] | Cross sectional study, Poland, Polish | n=156, M:F NA, BMI NA | Type of genetic testing: Single gene(s). Genes sequenced: NEUROD1 | None specified | Clinically suspected MODY diagnosed up to 30 years, recruited at University Hospital, Krakow, Poland; negative for HNF1A and/or GCK based on clinical presentation | Yield 1/156 (0.6%) |
| Skupien (2008) [63] | Case control study, Other: Poland, Polish | n=no new sequencing, M:F NA, BMI HNF1A-MODY 23+/-3.4 T2D 35.1 +/- 7.9 | Type of genetic testing: No new sequencing. Genes sequenced: Biomarker study for 1,5AG levels in HNF1A-MODY | None specified | HNF1A-MODY cases, T2D and non-diabetic control groups. | Biomarker study: 1,5 AG could discriminate HNF1A-MODY from T2D with a ROC C-statistic of 0.73, sensitivity 73%, specificity 65% |
| Molven (2008) [64] | Cohort study, Norway, | n=315, M:F NA, BMI NA | Type of genetic testing: Single gene. Genes sequenced: INS (in those negative for HNF1A, GCK, KCNJ11) | None specified | Two populations based registries NCDR and The Norwegian MODY registry. Children from NCDR are <18 yrs. Subjects from the MODY registry can have any age if at least 2 criteria are fulfilled | Overall yield 2/315(0.6%);<br>NCDR 1/124 (0.8%);<br>MODY registry 1/62 who met all MODY criteria. |
| Bellanné-Chantelot (2011) [65] | Cohort study, France, French | n=487, M:F 228 :251, BMI NA | Type of genetic testing: Sanger sequencing. Genes sequenced: HNF1A, GCK, HNF4A | Local lab guidelines | Referrals with a strong clinical suspicion of MODY because of a clinical suspicion of HNF1A-MODY, based on a diagnosis of diabetes at age < 40 years, a family history of diabetes in at least two generations and the absence of obesity. | HNF1A-MODY: 196/487 (40%). |

| | | | | | | |
|---|---|---|---|---|---|---|
| Thanabalasingham (2011) [66] | Cross sectional study, Other: UK, France, Denmark, Finland, Slovakia, Poland, Norway, European | n=1497, M:F Summary data not available, BMI Summary data not available | Type of genetic testing: No new sequencing. Genes sequenced: investigation of CRP levels in MODY subtypes | None specified | Individuals with HNF1A-MODY, GCK-MODY, HNF4A-MODY or T2D diagnosed <=45 yrs | Biomarker study: hsCRP discriminated HNF1A-MODY from T2D with a ROC C-statistic of 0.79-0.97. |
| Shields 2012 [67] | Case control study, UK, White European | n=594, M:F 473:718, BMI NA | Type of genetic testing: large NGS panel, Genes sequenced: HNF1A, HNF4A, GCK | ACMG | Individuals referred for MODY testing in national UK centre diagnosed up to 35 years. MODY probability model derived by logistic regression comparing MODY groups with gold standard T1D and T2D | Any MODY vs T1D: ROC C-statistic 0.95, 87% sensitivity and 88 specificity for a probability of 40%. Any MODY vs T2D: ROC C-statistic 0.98, Sensitivity 92%, Specificity 95% for a probability of 60% |
| Szopa (2019) [68] | Case control study, Other: Poland, Polish | n=165, M:F 157 :199, BMI | Type of genetic testing: No new sequencing. Genes sequenced: Investigation of biomarkers in HNF1A; GCK | None specified | Cases of 77 HNF1A, 88 GCK, controls: 99 T1D, 91 T2D | Testing a decision algorithm using several biomarkers. 92.9% of T1D, 84.8% of T2D, 64.9% of HNF1A-MODY, and 52.3% of GCK-MODY were correctly identified, C-peptide and clinical features performed best and overall MODY subgroups could not be discriminated with the model. |
| Pal (2010) [69] | Cohort study, United Kingdom, 90% White British | n=no new genetic sequencing, M:F NA, BMI NA | Type of genetic testing: No new sequencing. Genes sequenced: Investigation of 1,5AG levels in HNF1A; GCK; HNF4A | None specified | Participants with known MODY (HNF1A or GCK) and Type 1, LADA and Type 2 diabetes who underwent measurement of 1,5 AG. This is a replication study. | Biomarker study of 1,5AG levels. 1,5AG is lower in HNF1A-MODY than T2D and higher in GCK-MODY than other groups, ROC C-statistic 0.79 for GCK-MODY vs T2D. |
| Patel 2022 [70] | Cross-sectional study from seven paediatric diabetes clinics across | n=236, M:F NA, BMI NA | *Type of genetic testing: Large Monogenic Diabetes Panel. Genes Sequenced: ABCC8, AGPAT2, AKT2, APPL1, BSCL2, CISD2, CEL, COQ2,* | None specified | Combination of T1D-GRS and autoantibody status to select 236 children with high to moderate likelihood of monogenic diabetes (111 | 34/236 including 11 GCK, 3 HNF1A, 1 HNF4A, 2 HNF1B, 1 KCNJ11, 2 INS, 1 m.3243A>G, |

| | | | | | | |
|---|---|---|---|---|---|---|
| | Turkey; European ancestry | | *CTLA4, EIF2AK3, FOXP3, GATA4, GATA6, GCK, GLIS3, HNF1A, HNF4A, HNF1B, IER3IP1, IL2RA, INS, INSR, ITCH, KCNJ11, LMNA, LRBA, MNX1, NEUROD1, NEUROG3, NKX2-2, m.3243A>G, PAX6, PCBD1, PDX1, PIK3R1, PLIN1, POLD1, PPARG, PTF1A, RFX6, SIRT1, SLC2A2, SLC19A2, SLC29A3, STAT1, STAT3, STAT5B, TRMT10A, WFS1, ZFP57* | | children with low T1D-GRS irrespective if islet autoantibody status and 125 islet-autoantibody-negative children with moderate T1D GRS | 1 PTF1A, 7 WFS1, 3 SLC19A2, 1 SLC29A3, 1 TRMT10A |
| Besser (2011) [71] | Case control study, United Kingdom, | n=, M:F 90:110, BMI NA | Type of genetic testing: No new sequencing. Genes sequenced: Investigation of Urinary C-Peptide levels in HNF1A-MODY | None specified | Individuals with known MODY compared to T1 and T2 | Biomarker study of Urinary C-peptide. ROC C-statistic 0.98 for discriminating long duration T1D from HNF1A/4A-MODY |
| Flannick (2013) [72] | Cohort study, European and African American | n=4003, M:F 1745:2259, BMI range 23.6 to 34.2 depending on cohort | Type of genetic testing: Large MODY panel. Genes sequenced: HNF1A, GCK, HNF4A, HNF1B, PDX1, INS, NEUROD1 | In-house rules using computational prediction and prior reports | Individuals drawn from three population-based cohorts including Framingham Heart Study Offspring cohort (randomly ascertained 1541), Jackson Heart Study (1691), and extremes of T2D genetic risk from Finnish and Swedish cohorts (771). | Overall yield 121/4003 (0.03%) 13 Nonsynonymous MAF>1% and 108 Nonsynoymous MAF <1% of which 21 possibly pathogenic, 25 HGMD (Human Gene Mutation Database), 6 Putative pathogenic |
| Weinreich (2015) [73] | Cross sectional study, Netherlands, European | n=1319, M:F NA, BMI NA | Type of genetic testing: Single. Genes sequenced: HNF1A, HNF4A, GCK | None specified | People with clinically suspected MODY referred to molecular testing centre over 10 years | Any MODY: 502/1319 (38%) including HNF1A 222 and GCK 204 |
| **Monogenic Diabetes: Non-European** | | | | | | |
| HuaTan (2022) [74] | Cohort study, Singapore | n=267, M:F NA, BMI <32.5 | Type of genetic testing: Large MODY panel (>= 5 genes). Genes sequenced: HNF1B | None specified | Diabetes diagnosed up to age 35 with a clinical suspicion of MODY (BMI<32.5, islet antibody negative). | 40/267 (15%) diagnosed with MODY including 4/267 (1.5%), *HNF1B* |

| | | | | | | |
|---|---|---|---|---|---|---|
| Kleinberger (2018) [75] | Cohort study, United States, Hispanic, non-Hispanic white, non-Hispanic black | n=488, M:F 177:311, BMI Z-score:2.31±0.41 | Type of genetic testing: Large MODY panel (>= 5 genes). Genes sequenced: 40 autosomal genes known or predicted to cause monogenic diabetes, including 13 known MODY genes | ACMG | TODAY cohort: adolescents age 10–17 diagnosed with T2D within 2 years of study enrollment, C-peptide present, islet antibodies absent | Overweight/obese adolescents diagnosed with type 2 diabetes (T2D): 22/488 including HNF4A (7), GCK (7), HNF1A (5), INS (2), KLF11 (1) |
| Mifsud (2022) [76] | Cohort study, France, Multiple ethnicities | n=1975, M:F 937:1038, BMI NA | Type of genetic testing: Large MODY panel (>= 5 genes). Genes sequenced: GCK, HNF1A, HNF4A, HNF1B, ABCC8, KCNJ11, INS | ACMG | Adult probands (42% non-EuroCaucasians), selected on the absence of diabetes autoantibodies and ≥2 of the following criteria: age ≤40 years, BMI <30 kg/m2 at diagnosis, and a family history of diabetes in ≥2 generations | Absence of antibodies and 2/3 of (1) age at diabetes diagnosis ≥15 years and ≤40 years in the proband, or in at least two relatives with diabetes; (2) the absence of obesity in the proband or in at least two relatives with diabetes; (3) 2 gen family history of diabetes: 315/1975 |
| deSantana (2019) [77] | Cohort study, Brazil, Brazilian | n=102, M:F NA, BMI NA | Type of genetic testing: Large MODY panel (>= 5 genes). Genes sequenced: HNF1A; GCK; HNF4A; HNF1B; PDX1, NEUROD1, CEL, INS, KCNJ11, ABCC8, APPL1 | ACMG | Probands referred for MODY testing, diagnosed up to 35 years with negative testing for HNF4A, GCK, and HNF1A and deletions in HNF1B excluded | Yield 13/102 (12.7%) including 8 GCK, 1 PDX1, 2 HNF1B, 1 NEUROD1, 1 ABCC8 |
| Mohan (2018) [78] | Cross sectional study, India and US, Southern India | n=152, M:F 96:56, BMI 24.29 | Type of genetic testing: Whole exome (n=31), targeted exome (n=150). Genes sequenced: Exome | None specified | Clinically diagnosed with MODY, up to 30 years at diagnosis | <30 years, negative for auto antibodies, three-generation family history: 39/152 including HNF1A: 11/152, ABCC8: 5/152 |
| Breidbart (2021) [79] | Cross sectional study, United States, Asian 10.6%, Black 3.1%, European 53.1%, Latina 23.8%, mixed 7.5% | n=160, M:F 84:76, BMI NA | Type of genetic testing: GCK/HNF1A followed by Exome sequencing. Genes sequenced: GCK and HNF1A first | ACMG | Probands referred to MODY registry, neonatal diabetes diagnosed up to 12 months excluded | Yield of full selected cohort: 60/160 including GCK (45), HNF1A (11), HNF4A (1), PDX1 (1), KCNJ11 (1), HNF1B (1) |

| | | | | | | |
|---|---|---|---|---|---|---|
| Giuffrida (2017) [80] | Cohort study, Brazil, Brazilian | n=311, M:F 64% males (GCK probands). 40% males (HNF1A probands), BMI 19 ±6 (GCK,probands only), 25±6(HNF1A,probands only) | Type of genetic testing: Single gene(s). Genes sequenced: HNF1A; GCK | None specified | Multicentre study of 140 probands with clinical characteristics of MODY | Clinical profile consistent with MODY in Brazilian probands and families: Total 103/311 including 72 GCK, 31 HNF1A |
| Tonooka (2002) [81] | Cohort study, Japan, Japanese | n=203, M:F 79:124, BMI 23.7 +/- 6 | Type of genetic testing: Single gene(s). Genes sequenced: HNF1A | None specified | Non-type 1 diabetes dx before 15 years of age. Glycosuria picked up by annual school health exam. | Yield 18/203 |
| Horikawa (2014) [82] | Cross sectional study, Japan, Japanese | n=230, M:F 103:127, BMI 21.8 | Type of genetic testing: Single gene(s). Genes sequenced: HNF1A; HNF1B | None specified | Suspected MODY, onset up to 35 yrs, islet antibody negative, BMI <30 | Diabetes diagnosis </= 35 years, antibody negative: 30/230 |
| Karaca (2017) [83] | Cross sectional study, Turkey, Turkish | n=136, M:F 65:71, BMI NA | Type of genetic testing: Single gene(s). Genes sequenced: HNF1A | Bioinformatics | Clinically suspected MODY with age of onset up to 49 years, Islet antibody absent and family history of diabetes | Yield for strict MODY criteria 10/136 (7.35%) |
| Li (2021) [84] | Cross sectional study, Other: China, Han Chinese | n=543, M:F 375:168, BMI probands 27.6 ± 4.9 controls 25.8 ± 3.5 | Type of genetic testing: Customized gene panel for ABCC8 and exon capture. Genes sequenced: ABCC8 | ACMG | Diagnosed with diabetes up to 40 years, antibody negative, C-peptide positive and without Mitochondrial 3243 variant | Yield 8/543 (1.8%) |
| Xu (2005) [85] | Cohort study, China, Chinese | n=146, M:F NA, BMI NA | Type of genetic testing: Single. Genes sequenced: HNF1A, HNF4A, GCK | None specified | Clinically suspected MODY with age of diagnosis <25years from diabetes clinics in Hong Kong | Any MODY 15/146 (10%) including HNF1A 13 cases, GCK 2 cases, HNF4A 0 cases |

| | | | | | | |
|---|---|---|---|---|---|---|
| Yaghootkar (2019) [86] | Cross sectional study, Iran/UK, Iranian | n=127, M:F 63:64, BMI NA | Type of genetic testing: Large MODY panel. Genes sequenced: 35 gene panel | ACMG | Children diagnosed 9months-5years from 2 hospital clinics, 32% consanguineous | Any MODY 6/126 (4.7%) including 1 GCK, 3 WFS1, 1 SLC19A2, 1 SLC29A3. T1GRS <15th centile for T1D identified all MODY Cases, 100% sensitive, 82% specific, ROC C-statistic 0.9. MODY cases were islet antibody negative. |
| Yamada (1997) [87] | Cross sectional study, Japan, Japanese | n=103, M:F NA, BMI 22+/-2.2 | Type of genetic testing: Single. Genes sequenced: HNF1A | None specified | Diagnosed >40years, attending a diabetes clinic | 0/103 (1 VUS) |
| Yang (2006) [88] | Case-control study, China, Chinese | n=192, M:F 54:40, BMI 24.2+/-0.78 | Type of genetic testing: Single. Genes sequenced: HNF1A | In-house rules, local control group. Some doubt over VUS. | Attending the Shanghai Diabetes Institute, not clinically T1D. Group A diag<40years, 1 relative with DM, Group B diag>40years, 3 gen FH | Whole group 3/192 (1.6%); young onset 2/94; older-onset familial 1/98 |
| Yorifuji (2018) [89] | Cohort study, Japan, Japanese | n=263, M:F NA, BMI median 45th centile | Type of genetic testing: Single . Genes sequenced: Sequential approach: Mt3243A>G, GCK-MODY, HNF1A, HNF4A, HNF1B, other genes if specific features | None specified | Clinically suspected MODY referred for genetic testing: <30years, islet antibody negative, CP present, non-obese, + group with incidental hyperglycaemia | Overall: 103/263 (39%) including Mt3243A.G 4, GCK 57, HNF1A 29, HNF4A 7, HNF1B 10, other 13 |

| | | | | | | |
|---|---|---|---|---|---|---|
| Zmyslowska 2022 [90] | Cohort study,Polish, European | n=542 probands, 290:252, BMI NA | *Type of genetic testing: Large Monogenic Diabetes Panel. Genes sequenced: ABCC8, AIRE, APPL1, BLK, CISD2, CEL, EIF2AK3, FOXP3, GATA4, GATA6, GCK, GCKR, GLIS3, GLUD1, HADH, HNF1A, HNF4A, HNF1B, INS, ISL1, KCNJ11, KLF11, MAFA, MAFB, MNX1, NEUROD1, NEUROG3, NKX2-2, NKX6-1, PAX4, PAX6, PDX1, PTF1A, RFX6, WFS1* | ACMG | Referred to the Genetic Outpatient Clinic in Poland from 12 Polish Diabetes Centers, with presence of hyperglycaemia or diabetes mellitus, positive family history of diabetes, absence of autoantibodies, preserved insulin secretion based on fasting C-peptide or insulin values and absence of additional organ-specific symptoms. | Overall Yield 192/542 including GCK 148, HNF1A 19, HNF4A 5, HNF1B 6, ABCC8 2, KCNJ11 8, RFX6 1, PDX1 5, WFS1 1, MAFA 1, APPL1 3 |
| **HNF1B** | | | | | | |
| Wang (2012) [91] | Cohort study, China, Chinese | n=104, M:F 74:30, BMI NA | Type of genetic testing: Single gene(s). Genes sequenced: HNF1B | Local lab guidelines | Probands from Shanghai Diabetes Institute, mean age 64.7+/-13.7 years, negative autoantibodies and either renal structural abnormalities or impaired renal function | Overall yield 3/104 (2.9%) |
| Faguer (2014) [92] | Cohort study, France, | n=433, M:F NA, BMI NA | Type of genetic testing: Single gene(s). Genes sequenced: HNF1B | None specified | Cohort with known HNF1B-MODY, clinical features used to derive an "HNF1B-score" (using previously published data), 65% aged <=16years | For an HNF1B score of >8: ROC C-statistic 0.78 (0.72-0.83), sensitivity 92.2%, specificity 41.1%, NPV >99% |
| Clissold (2015) [93] | Cohort study, United Kingdom, | n=686, M:F 343:343, BMI NA | Type of genetic testing: Single gene(s). Genes sequenced: HNF1B | None specified | Probands referred for HNF1B testing at Exeter between 1998-2012 | HNF1B score of >8, sensitivity 80%, specificity 38%, PPV 31%, NPV 85%. |

| | | | | | | |
|---|---|---|---|---|---|---|
| Edghill (2013) [94] | Cohort study, United Kingdom, | n=124, M:F NA, BMI median 24 | Type of genetic testing: HNF1B deletion via MLPA only. Genes sequenced: HNF1B | None specified | Patients referred to the Molecular Genetics Laboratory at the Royal Devon and Exeter Hospital for genetic testing to investigate a possible diagnosis of MODY, but no mutation had been found by sequence analysis. | Yield 3/124 (2.4%) |
| **Lipodystrophy** | | | | | | |
| Gong (2021) [95] | Cohort study, China, Han Chinese | n=1002, M:F NA, BMI NA | Type of genetic testing: Single gene(s). Genes sequenced: PPARG | ACMG | T2D diagnosed 18 -40 years, islet antibody negative, C-peptide positive recruited at the Peking University People'sHospital 2014-18 | Yield 6/1006 (0.6%) |
| Decaudain (2007) [96] | Cohort study, France, French | n=277, M:F NA, BMI NA | Type of genetic testing: Single gene(s). Genes sequenced: LMNA | None specified | Probands with suspected severe insulin resistance (lipodystrophy and/or android body habitus, insulin resistance or altered glucose tolerance) between 2002-6 | Yield 27/277 (9.7%) |
| **Biomarkers** | | | | | | |

| | | | | | | |
|---|---|---|---|---|---|---|
| Ma (2020)[97] | case/control + cohort, China, Chinese | n=535, M:F 375:176, BMI NA | Type of genetic testing: Single gene(s). Genes sequenced: HNF1A; GCK; HNF4A; HNF1B; MT3243A>G | ACMG | Control groups of normoglycaemia, early-onset diabetes (diagnosed at 14–40 years) and HNF1A-MODY were used to define normal ranges for clinical biomarkers (body mass index (BMI), HDL-c and hsCRP). A second group with early onset diabetes were used to validate the effectiveness of the biomarkers established in this study for HNF1A-MODY | Used HNF1A-Clinical screening strategy in cohort of 410 subjects with young onset type 2 diabetes meeting 3 or 4 biomarker criteria for BMI<28 kg/m2, hs-CRP <0.75 mg/L, Fasting insulin <102 pmol/L and HDL-c >1.12 mmol/L levels received genetic testing and yield 17/182. |
| McDonald (2011) [98] | Cohort study, United Kingdom, European | n=540, M:F NA, BMI NA | Type of genetic testing: No new sequencing. Genes sequenced: Investigation of hsCRP in MODY subtypes | *None specified* | 540 patients with a confirmed genetic dx of MODY (220 HNF1A, 245 GCK, 54 HNF4A, and 21 HNF1B). 53 patients with T1D and 157 with T2D | Biomarker study: replication of hsCRP levels. hsCRP was lower in HNF1A-MODY than T2D, T1D, HNF4A-MODY, GCK-MODY and HNF1B-MODY. ROC C-statistic 0.84 HNF1A-MODY vs T2D |
| McDonald (2011) [99] | Case control study, United Kingdom, | n=508, M:F NA, BMI NA | Type of genetic testing: No new sequencing. Genes sequenced: Investigation of prevalence of islet antibodies in MODY | None specified | Antibody status was investigated in participants with a confirmed genetic diagnosis of MODY (263 probands and 245 family members): 227GCK, 229HNF1Aand 52HNF4A. In addition, serum from 98 patients (median age of 15 years ;) diagnosed with Type 1 diabetes within the past 6 months was analyzed. Fifty samples were from the Bart's-Oxford (BOX) family study and 48 from the Diabetes Autoantibody Standardization Programme. To establish an antibody titre cut-off for GAD and IA-2antibodies, we tested 500 routine clinical samples of patients aged between 18 and 75 years | Antibodies found in 5/508 (<1%) of those with MODY |

| | | | | | | |
|---|---|---|---|---|---|---|
| | | | | | without a clinical history of diabetes and an HbA1clevel of less than 6.0% (42 mmol⁄mol). | |
| McDonald (2012) [100] | Cohort study, United Kingdom, | n=564, M:F 327:237, BMI NA | Type of genetic testing: No new sequencing. Genes sequenced: Study of lipid biomarkers in HNF1A-MODY | None specified | Individuals with HNF1A, non-diabetic and T2D | Biomarker study: HDL-Chol was the best lipid measurement to discriminate HNF1A-MODY from T2D, ROC C-statistic 0.76, sensitivity 75%, specificity 64% |

#### Supplementary Table 4: Complete set of papers extracted for question 2. How to test for monogenic diabetes

| Study Details | Individuals tested | Test methodology and number of different genes tested | Genes analysed | Individuals diagnosed, diagnostic yield and number of different genetic subtypes diagnosed | Number of patients diagnosed by subtype: |
|---|---|---|---|---|---|
| Abbasi, 2018 Iran [1] | 60 neonates with NDM | Sanger sequencing; 4 | *ABCC8; EIF2AK3; INS; KCNJ11* | 11 (18%); all *EIF2AK3* | 11 *EIF2AK3* |
| Al-Senani, 2018 Oman [2] | 18 neonates with NDM | tNGS (RNA baits) and MS-PCR; 24 | 6q24*; ABCC8; EIF2AK3; FOXP3; GATA4; GATA6; GCK; GLIS3; HNF1B; INS; KCNJ11; IER3IP1; IL2RA; LRBA; NEUROD1; NEUROG3; NKX2-2; PDX1; PTF1A; RFX6; SLC2A2; SLC19A2; STAT3; WFS1; ZFP57* | 9 (50%); 2 | 5 *GCK*, 1 *KCNJ11* |
| Al-Kandari, 2021 Kuwait [3] | 31 children with suspected MODY | MLPA and targeted exome sequencing (gene panel); 22 | *ABCC8; CISD2; CEL; GATA6; GCK; HNF1A; HNF4A; HNF1B; INS; INSR; KCNJ11; LMNA; NEUROD1;* m.3243A>G*; PAX6; PDX1; PLIN1; PPARG; RFX6; WFS1; ZFP57* | 7 (23%); 5 | 1 *GCK*, 2 *HNF1A*, 1 *HNF4A*, 2 *HNF1B*, 1 *PDX* |
| Al-Khawaga, 2019 Qatar [4] | 7 neonates with NDM | WES (gene agnostic), WGS (gene agnostic) and CNV analysis as part of tNGS/exome/genome sequencing; Not Stated | *Whole exome; Whole genome* | 7 (100%); 6 | 1 *GCK*, 1 *HNF1B*, 2 *INS*, 1 *EIF2AK3*, 1 *PTF1A* |
| Alkorta-Aranburu, 2014 USA [5] | 44 children and adults with suspected MODY and 32 with neonates with NDM | WES (gene agnostic), WGS (gene agnostic) and CNV analysis as part of tNGS/exome/genome sequencing; Not Stated | *ABCC8; AKT2; ALMS1; BLK; CISD2; CEL; CP; DCAF17; EIF2AK3; FOXP3; GATA6; GCK; GLIS3; GLUD1; HADH; HNF1A; HNF4A; HNF1B; IER3IP1; INS; INSR; KCNJ11; NEUROD1; NEUROG3; PAX4; PAX6; PDX1; PTF1A; RFX6; SLC2A2; SLC19A2; SLC29A3; TBC1D4; WFS1; ZFP57* | 12 with MODY (27%); 2 and 7 with NDM (22%); 5 | 11 *GCK*, 2 *HNF1A*, 1 *ABCC8*, 2 *KCNJ11*, 2 *INS*, 1 *EIF2AK3* |
| Alkorta-Aranburu, 2016 USA [6] | 22 neonates with NDM | MS-MLPA and tNGS (RNA baits); 11 | 6q24*; ABCC8; EIF2AK3; FOXP3; GATA4; GCK; INS; KCNJ11; MNX1; NKX2-2; PDX1; ZFP57* | 14 (64%), 5 | 2 *ABCC8*, 2 *KCNJ11*, 1 *INS*, 1 *FOXP3*, 1 6q24 |
| Ang, 2016 Singapore [7] | 84 adults with suspected MODY | tNGS (PCR using Ion AmpliSeq) and Genotyping (TaqMan); 16 | *ABCC8; BLK; CEL; GCK; HNF1A; HNF4A; HNF1B; INS; INSR; KCNJ11; KLF11; LMNA; NEUROD1;* m.3243A>G*; PAX4; PDX1; PPARG* | 13 (16%), 7 | 1 *GCK*, 4 *HNF1A*, 1 *HNF4A*, 1 *ABCC8*, 1 *KCNJ11*, 2 m.3243A>G |
| Anik, 2015 Turkey [8] | 42 children with suspected MODY | tNGS (PCR using Illumina Nextera XT); 11 | *BLK; CEL; GCK; HNF1A; HNF4A; HNF1B; INS; KLF11; NEUROD1; PAX4; PDX1* | 12 (29%), 5 | 8 *GCK*, 1 *HNF1A*, 1 *HNF1B*, 1 *PDX1*, 1 *BLK* |
| Antosik, 2016 Poland [9] | 1 neonate with NDM | tNGS (RNA baits); 1 | *GCK* | 1 (100%), 1 | 1 *GCK* |

| | | | | | |
|---|---|---|---|---|---|
| Ateş, 2021 Turkey [10] | 182 adults with suspected MODY | tNGS (PCR using Agilent MODY-MASTR assay); 7 | *ABCC8; GCK; HNF1A; HNF4A; HNF1B; INS; KCNJ11* | 30 (17%), 6 | 10 *GCK*, 9 *HNF1A*, 2 *HNF4A*, 2 *HNF1B*, 6 *ABCC8*, 1 *KCNJ11* |
| Bansal, 2017 USA [11] | 4016 adults with type 2 DM | tNGS (RNA baits); 22 | *ABCC8; BLK; CEL; EIF2AK3; GATA6; GCK; GLIS3; HNF1A; HNF4A; HNF1B; INS; KCNJ11; KLF11; NEUROD1; NEUROG3; PAX4; PAX6; PDX1; PPARG; RFX6; SLC19A2; WFS1* | 53 (1%), 8 | 21 *GCK*, 17 *HNF1A*, 5 *HNF4A*, 1 *HNF1B*, 6 *ABCC8*, 1 *INS*, 1 *WFS1*, 1 *PPARG* |
| Berberich, 2021 Canada [12] | 57 children and adults wth suspected MODY | tNGS (DNA baits) and CNV analysis as part of tNGS; 14 | *ABCC8; CEL; GCK; HNF1A; HNF4A; HNF1B; INS; INSR; KCNJ11; LMNA; NEUROD1; PDX1; PPARG; RFX6* | 3 (5%), 1 | 3 *HNF1B* |
| Carette, 2010 France [13] | 84 children and adults wth suspected MODY | Sanger sequencing and MLPA; 2 | *HNF1A; HNF4A* | 8 (10%), 2 | 2 *HNF1A*, 6 *HNF4A* |
| Caswell, 2020 UK [14] | 33 at-risk pregnancies | Droplet digital PCR; 1 | *GCK* | 21 (64%), 1 | 21 *GCK* |
| Chambers, 2016 USA [15] | 97 children and adults wth suspected MODY | Sanger sequencing; 5 | *GCK; HNF1A; HNF4A; HNF1B; PDX1* | 20 (21%), 3 | 8 *GCK*, 9 *HNF1A*, 3 *HNF4A* |
| Colclough, 2022 UK [16] | 1280 children and adults wth suspected MODY | tNGS (RNA baits) and CNV analysis as part of tNGS; 27 | *ABCC8; CISD2; CEL; GATA4; GATA6; GCK; HNF1A; HNF4A; HNF1B; INS; INSR; KCNJ11; LMNA; NEUROD1;* m.3243A>G; *PAX6; PCBD1; PDX1; PLIN1; POLD1; PPARG; RFX6; SLC29A3; TRMT10A; WFS1; ZBTB20; ZFP57* | 297 (23%), 17 | 66 *GCK*, 98 *HNF1A*, 42 *HNF4A*, 18 *HNF1B*, 11 *ABCC8*, 5 *KCNJ11*, 6 *INS*, 8 *RFX6*, 3 *NEUROD1*, 2 *PDX1*, 24 m.3243A>G, 6 *WFS1*, 4 *INSR*, 1 *PPARG*, 1 *TRMT10A*, 1 *SLC29A3*, 1 *GATA6* |
| De Franco, 2015 UK [17] | 1020 neonates with NDM | Sanger sequencing, MS-MLPA) and tNGS (RNA baits) and CNV analysis as part of tNGS; 23 | 6q24*; ABCC8; EIF2AK3; FOXP3; GATA4; GATA6; GCK; GLIS3; HNF1B; IER3IP1; INS; KCNJ11; IER3IP1; MNX1; NEUROD1; NEUROG3; NKX2-2; PDX1; PTF1A; RFX6; SLC2A2; SLC19A2; ZFP57* | 840 (82%), 22 | 30 *GCK*, 2 *HNF1B*, 150 *ABCC8*, 240 *KCNJ11*, 110 *INS*, 1 *RFX6*, 3 *NEUROD1*, 6 *PDX1*, 76 *EIF2AK3*, 22 *PTF1A*, 14 *FOXP3*, 113 6q24, 7 *SLC19A2*, 4 *GATA6*, 9 *GLIS3*, 1 *IER3IP1*, 1 *MNX1*, 1 *NEUROG3*, 2 *NKX2-2*, 6 *SLC2A2*, 12 *ZFP57* |
| Donath, 2019 France [18] | 1564 children and adults wth suspected MODY | MLPA and tNGS (PCR using Agilent MODY-MASTR assay); 7 | *ABCC8; GCK; HNF1A; HNF4A; HNF1B; INS; KCNJ11* | 254 (16%), 7 | 109 *GCK*, 82 *HNF1A*, 25 *HNF4A*, 15 *HNF1B*, 8 *ABCC8*, 3 *KCNJ11*, 7 *INS*, |
| Ellard, 2013 UK [19] | 33 children with suspected MODY and 49 neonates with NDM | tNGS (RNA baits) and CNV analysis as part of tNGS; 29 | *ABCC8; BLK; CEL; EIF2AK3; FOXP3; GATA4; GATA6; GCK; GLIS3; HNF1A; HNF4A; HNF1B; INS; KCNJ11; KLF11; IER3IP1; LMNA; NEUROD1; NEUROG3;* m.3243A>G; *PAX6; PDX1; PPARG; PTF1A; RFX6; SLC2A2; SLC19A2; WFS1; ZFP57* | 5 with MODY (15%), 4 and 9 with NDM (18%), 6 | 3 *GCK*, 1 *HNF4A*, 1 *HNF1B*, 1 *ABCC8*, 1 *PDX1*, 2 m.3243A>G, 1 *EIF2AK3*, 2 *SLC19A2*, 2 *GATA6* |

| | | | | | |
|---|---|---|---|---|---|
| Ellard, 2007 UK [20] | 90 children and adults wth suspected MODY | MLPA; 3 | *GCK; HNF1A; HNF4A* | 6 (7%), 2 | 1 *GCK*, 5 *HNF1A* |
| Johansson, 2012 Norway [21] | 9 children and adults wth suspected MODY | targeted exome sequencing (gene panel); 109 | *ABCC8; BLK; CISD2; CEL; EIF2AK3; GATA4; GATA6; GCK; GLIS3; HADH; HNF1A; HNF4A; HNF1B; INSR; KCNJ11; KLF11; LMNA; MNX1; NEUROD1; NEUROG3; NKX2-2; PAX4; PAX6; PDX1; POLD1; PPARG; PTF1A; RFX6; SLC2A2; WFS1* | 3 (33%), 3 | 1 *HNF4A*, 1 *ABCC8*, 1 *PPARG* |
| Laimon, 2021 Egypt [22] | 26 neonates with NDM | Sanger sequencing, tNGS (RNA baits) and CNV analysis as part of tNGS; 22 | *ABCC8; EIF2AK3; FOXP3; GATA4; GATA6; GCK; GLIS3; HNF1B; INS; KCNJ11; IER3IP1; NEUROD1; NEUROG3; NKX2-2; PDX1; PTF1A; RFX6; SLC2A2; SLC19A2; STAT3; WFS1* | 14 (54%), 7 | 1 *GCK*, 2 *ABCC8*, 2 *KCNJ11*, 4 *INS*, 3 *EIF2AK3*, 1 *SLC19A2*, 1 *INSR* |
| Patel, 2022 Greece [23] | 236 children with suspected MODY | tNGS (RNA baits) and CNV analysis as part of tNGS; 51 | *ABCC8; AGPAT2; AKT2; APPL1; BSCL2; CISD2; CEL; COQ2; CTLA4; EIF2AK3; FOXP3; GATA4; GATA6; GCK; GLIS3; HNF1A; HNF4A; HNF1B; IER3IP1; IL2RA; INS; INSR; ITCH; KCNJ11; LMNA; LRBA; MNX1; NEUROD1; NEUROG3; NKX2-2;* m.3243A>G*; PAX6; PCBD1; PDX1; PIK3R1; PLIN1; POLD1; PPARG; PTF1A; RFX6; SIRT1; SLC2A2; SLC19A2; SLC29A3; STAT1; STAT3; STAT5B; TRMT10A; WFS1; ZFP57* | 34 (14%), 12 | 11 *GCK*, 3 *HNF1A*, 1 *HNF4A*, 2 *HNF1B*, 1 *KCNJ11*, 2 *INS*, 1 m.3243A>G, 1 *PTF1A*, 7 *WFS1*, 3 *SLC19A2*, 1 *SLC29A3*, 1 *TRMT10A* |
| Patouni, 2021 Greece [24] | 1 child with type 1 DM | Sanger sequencing and MLPA; 3 | *GCK; HNF1A; HNF1B* | 1 (100%), 2 | 1 *HNF1A*, 1 *HNF1B* |
| Pruhova, 2010 Czech Republic [25] | 140 children and adults wth suspected MODY | Sanger sequencing and MLPA; 1 | *GCK* | 103 (74%), 1 | 103 *GCK* |
| Saint-Martin, 2022 France [26] | 1676 children and adults wth suspected MODY | tNGS (RNA baits) and CNV analysis as part of tNGS; 18 | *ABCC8; CISD2; GATA4; GATA6; GCK; HNF1A; HNF4A; HNF1B; INS; INSR; KCNJ11; NEUROD1; PDX1; PLIN1; RFX6; TRMT10A; WFS1* | 307 (18%), 13 | 124 *GCK*, 63 *HNF1A*, 35 *HNF4A*, 18 *HNF1B*, 8 *ABCC8*, 1 *KCNJ11*, 4 *INS*, 7 *RFX6*, 5 *NEUROD1*, 7 *PDX1*, 24 m.3243A>G, 9 *WFS1*, 2 *INSR* |
| Singh, 2006 UK [27] | 230 adults with type 2 DM and a negative m.3243A>G result using PCR-RFLP | Genotyping (TaqMan); 1 | m.3243A>G | 0 (0%), 0 | |
| Støy, 2008 USA [28] | 77 neonates with NDM | Sanger sequencing and MS-MLPA; 4 | 6q24*; ABCC8; INS; KCNJ11* | 23 (30%), 3 | 14 *KCNJ11*, 7 *INS*, 2 6q24 |
| Tosur, 2021 USA [29] | 10 children with suspected MODY | targeted exome sequencing (gene panel); 70 | *ABCC8; AGPAT2; AIRE; AKT2; APPL1; BLK; BSCL2; CDKN1C; CISD2; CEL; COQ2; COQ9; CP; CTLA4; DCAF17; DNAJC3; EIF2AK3;* | 2 (20%), 2 | 1 *INS*, 1 *RFX6* |

| | | | | | |
|---|---|---|---|---|---|
| | | | *EIF2S3; FOXP3; GATA4; GATA6; GCK; GLIS3; GLUD1; HADH; HNF1A; HNF4A; HNF1B; IER3IP1; IL2RA; INS; INSR; ITCH; KCNJ11; KLF11; LMNA; LPL; LRBA; MNX1; NEUROD1; NEUROG3; NKX2; NKX2-2;* m.3243A>G; *PAX4; PAX6; PCBD1; PDX1; PIK3R1; PLIN1; POLD1; PPARG; PTF1A; RFX6; SIRT1; SLC2A2; SLC19A2; SLC29A3; STAT1; STAT3; STAT5B; TNFAIP3; TRMT10A; WFS1; ZBTB20; ZFP57* | | |
| Weedon, 2014 UK [30] | 22 neonateswith NDM | targeted genome sequencing (homozygosity mapping); 1 | *PTF1A* | 10 (48%), 1 | 10 *PTF1A* |
| Yan, 2014 China [31] | 57 adults with suspected MIDD | Sanger sequencing and genotyping by Pyrosequencing, RFLP and HRM; 1 | m.3243A>G | 47 (83%), 1 | 47 m.3243A>G |
| Zmysłowska, 2022 Poland [32] | 542 children with suspected MODY | tNGS (RNA baits); 35 | *ABCC8; AIRE; APPL1; BLK; CISD2; CEL; EIF2AK3; FOXP3; GATA4; GATA6; GCK; GCKR; GLIS3; GLUD1; HADH; HNF1A; HNF4A; HNF1B; INS; ISL1; KCNJ11; KLF11; MAFA; MAFB; MNX1; NEUROD1; NEUROG3; NKX2-2; NKX6-1; PAX4; PAX6; PDX1; PTF1A; RFX6; WFS1* | 198 (37%), 11 | *GCK* 148, *HNF1A* 19, *HNF4A* 5, *HNF1B* 6, *ABCC8* 2, *KCNJ11* 8, *RFX6* 1, *PDX1* 5, *WFS1* 1, *MAFA* 1, *APPL1* 2 |

### Supplementary Figure 1

A

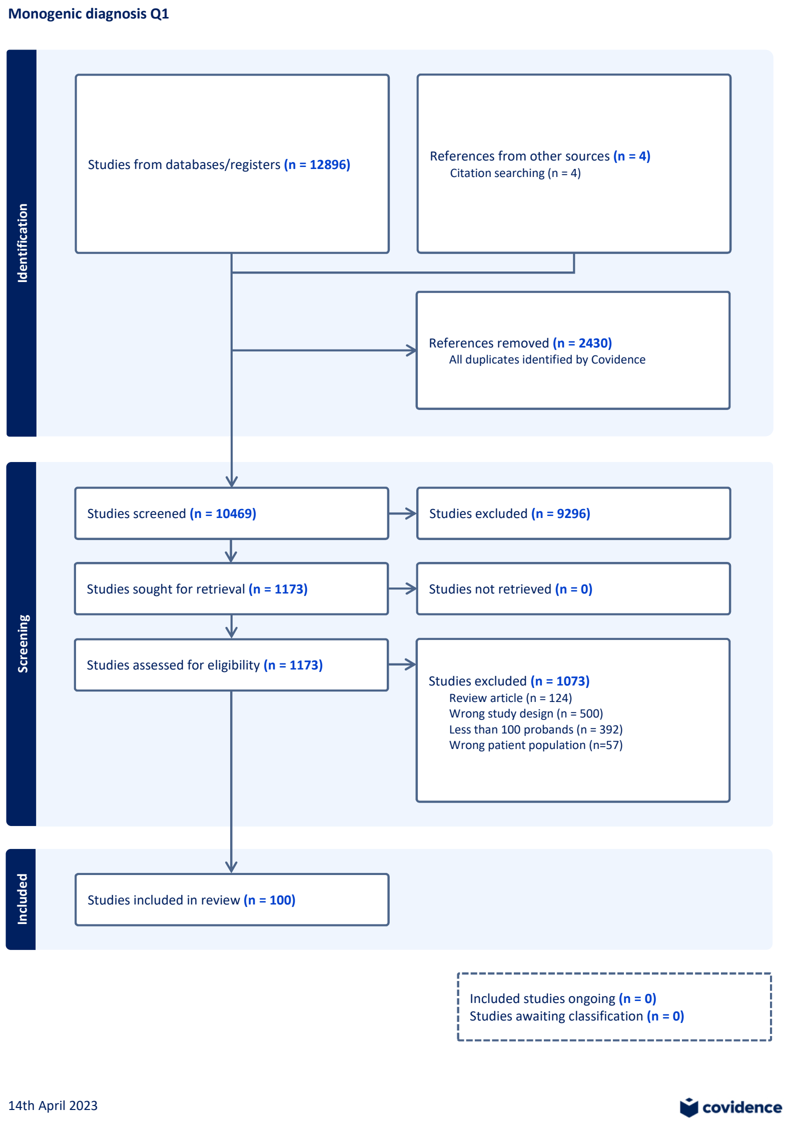


B

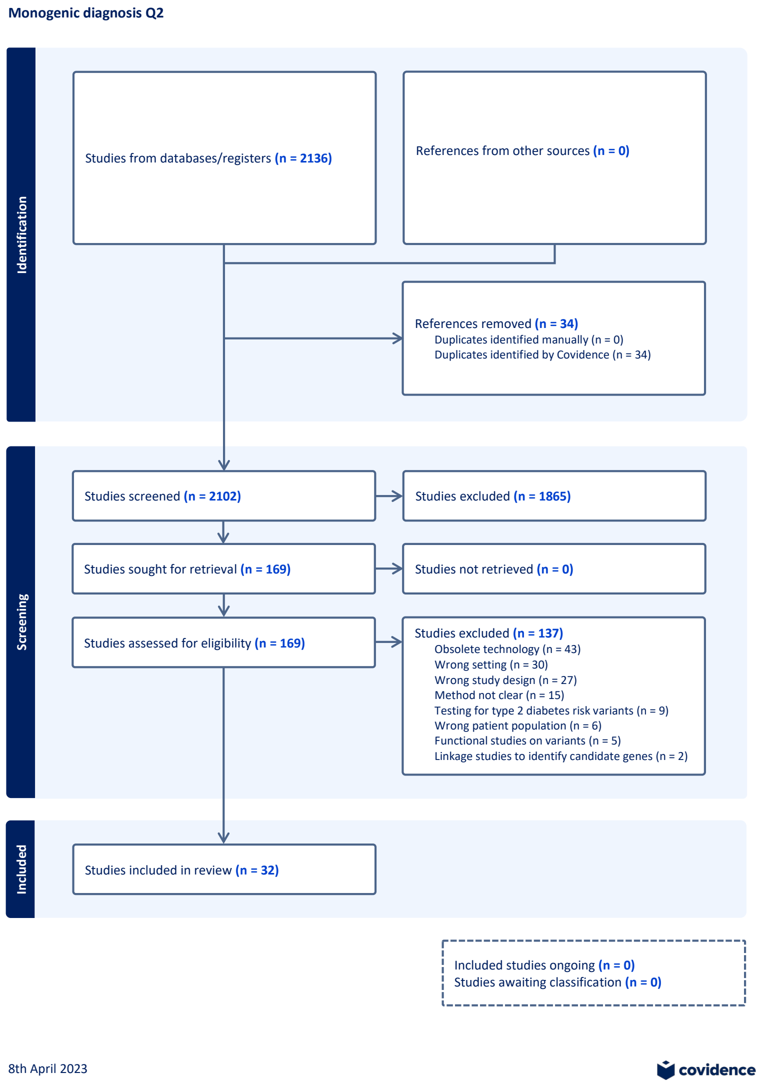
